# Multivariate analysis of metabolomic data to identify biological pathways modified by a clinical intervention

**DOI:** 10.64898/2025.11.28.25340231

**Authors:** Rachel M. Wood, Laura J. Corbin, Jane M. Blazeby, Chris A. Rogers, Nicholas J. Timpson, Daniel J. Lawson

## Abstract

High throughput metabolomic assays offer a huge opportunity to quantify the cellular processes underlying disease and intervention pathways. However, the multi-dimensional inter-relatedness between these processes coupled with the complex noisy measurement environment create a need for generation of new methods that move beyond simple pairwise associations. Here we develop a computationally simple, multivariate, relational comparison method called CLARITY to compare metabolomic data before and after an intervention. This generates a relational anomaly score that combines with traditional methods to increase classification performance of the underlying cause of changes to the levels of and covariances between metabolites. We demonstrate utility in the By-Band-Sleeve (BBS) clinical trial of bariatric surgery using NMR metabolomics data. On supplementing linear regression analysis with CLARITY, previously identified changes form two clusters that imply involvement in different underlying biological pathways. An additional cluster of metabolites are identified as undergoing a relational change which would not have been detected using traditional methods. Gathering insights about metabolites and the biomarkers they capture in the causal pathway between intervention and effect, from observations at scale, will inform the future design of modelling and laboratory experiments to capture the underlying biological process.

## Introduction

Metabolomics is the quantification of small molecules produced by metabolic processes as captured within a biological sample. As the end products of cellular regulatory processes, metabolite levels reflect the response of biological systems to genetic or environmental changes. Capturing cellular responses to endogenous and exogenous perturbations is valuable for the prediction, prognosis, diagnosis and aetiological dissection of disease. With rapid advances in high-throughput technology and bioinformatics now enabling absolute quantification of hundreds (or relative quantification of thousands) of metabolites from a single biological sample, the last decade has seen major growth in the use of metabolomics as a research tool in epidemiology. However, substantial challenges exist in the translation of this research into public health benefit, an issue that has been highlighted by those working directly in the field^1–3^.

Current technologies typically generate (semi-)quantitative data on hundreds to thousands of metabolites per sample analysed and this has prompted a shift from measuring one or a small number of ‘target’ metabolites (whose properties may be relatively well understood), to measuring snapshots of the entire metabolome. In some ways, this change mirrors that seen in the field of genomics in the early 2000s, when technological developments fuelled the step-change from candidate gene studies to genome-wide association studies (GWAS) – though based on a system (cellular activity) which does not attend to the more predictable (and exploited) patterns of variation observed in genetics. Complexity aside, the high throughput, omic-scale, approach to metabolite analysis has led to the use of standard association testing in large-scale epidemiological studies -‘metabolome-wide association studies’ (MWAS). This approach, by which an experimental factor is tested for metabolite association against a hypothesis of ‘no effect’^4^ in a univariate testing framework, has as its output a list of single ‘associated’ metabolites determined by the presence of sufficient evidence to justifiably be more than a chance event alone. Since 2008, this approach to metabolomic profiling has been used to identify biomarkers associated with a range of diseases and to characterise metabolic risk factors^5,6^, but arguably has not fully exploited the analytical opportunities available in this field and extracting meaningful information from studies of the human metabolome remains challenging.

Univariate analyses disregard interrelationships between levels of different metabolites that may co-occur either due to biochemical pathways or because of shared genetic and/or environmental influences. This failure has been identified as a major limitation given the system-wide view provided by metabolomics data^7^. Furthermore, the coexistence of high throughput molecular measures of differing type and derived from the same biosamples, is presenting an opportunity to consider multi-omic causal complexes which are really only maximised when considered in multivariate analyses. Whilst existing multivariate approaches – typically ‘factor analysis’ methods, e.g. principal component analysis (PCA) – go some way to addressing this problem, these also fail to explicitly uncover changes in the relationships between metabolites under different conditions. Although not routinely implemented within the field of epidemiology, correlation network analysis has the potential to reveal differential connectivity patterns between features. To date, this approach has been applied to a limited extent within metabolomics, for example, in the context of demographic characteristics (age and sex)^8^, cardiovascular risk^9^, bacterial growth in soft tissue infections^10^ and mortality in acute myocardial infarction patients^11^. Whilst these studies have found evidence for potentially meaningful differences in connectivity, the limitations of the correlation network approach are well established^12^.

Metabolite-metabolite networks are typically underpinned by correlation matrices that capture the pairwise similarity between metabolites. However, it is not always clear how best to translate the correlation matrix into a network, with optimal approaches being dependent on the specific application. A key question is which relations to include in the network; the most common approach to determine this is to use thresholding such that only those pairwise relationships whose correlation coefficient exceeds some prespecified (and essentially arbitrary) threshold for relevance are represented in the network. Then the network itself may then be subject to weighting – where pairwise correlation coefficients are retained as the edge weights. However, there is no consensus regarding what an appropriate threshold for edge inclusion might be (and the choice is known to influence results). Challenges also exist in how to deal with negative correlations, edges with low correlations increasing uncertainty in the network, false positives resulting from indirect associations, varying edge density across individuals (or groups) and small sample sizes (relative to the number of measured metabolites)^12^. Methods continue to be developed to address these complexities including, proportional thresholding (to address varying edge density) and sparsity constraint, e.g. Gaussian graphical models^12,13^, however, an alternative approach proposed by Masuda *et al*.^12^ is to avoid transforming the correlation matrix data into a network all together. In line with this recommendation, here we explore the application of an approach designed to compare heterogeneous data using (dis)similarity as an alternative to network analysis in the context of metabolomics data.

When two datasets describe the same entities (typically samples or individuals), many scientific questions can be phrased around whether the (dis)similarities between entities are conserved across strata of interest (be it time, study, treatment, group). Here we implement CLARITY, a method that quantifies consistency across datasets, identifies where inconsistencies arise and aids in their interpretation^14^. Applications of this software to date have been limited to exploratory analysis for anomaly detection, and only synthetic data in a bioinformatics context. Here we evaluate what is detected when the shared entities are common omics – in this case a shared set of nuclear magnetic resonance (NMR)-derived metabolites, with the two datasets representing different sampling timepoints. This allows an integration with regression methods, and by matching simulated data to a real dataset we can characterise the underlying network in terms of “direct” and/or “indirect” or inter-relational changes in metabolites that occur following a clinical intervention.

## Methods

### Study overview

Metabolite data are complex and a minimal model risks oversimplification. NMR data in particular is characterised by relatively high redundancy and whilst this can lead to over conservative corrections for multiple testing in univariate frameworks it means that these data are well-suited to data compression techniques. The same metabolite observed at different times will systematically differ in a way that is not explainable using observed confounders, whilst groups of metabolites will be correlated due to both observed and unobserved confounders. The signal of an intervention effect sits on top of this complex structure and is also expected to be complex in the sense of affecting multiple metabolites. **Figure 1** illustrates the structure of this model.

**Figure 1.**
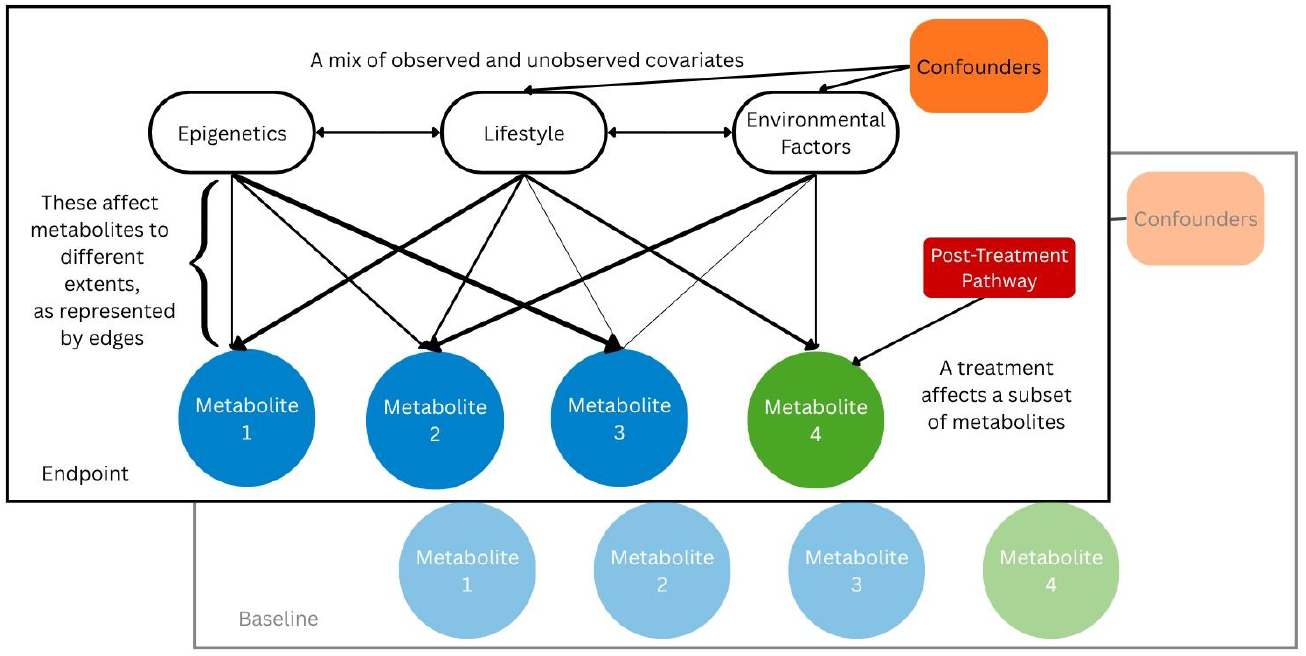
Conceptualisation of metabolite model. We consider two instances of the data (corresponding to before and after an intervention). Covariates - in our example epigenetics, lifestyle and environmental factors, affect the metabolites at strengths represented by edge widths. A cluster of covarying metabolites can be thought of as capturing part of a “metabolomic pathway”. In the endpoint, a post-treatment pathway variable is added which affects a subset of metabolites, which we are aiming to identify with our methods. Edge weights and confounders correlate, but differ, from baseline to endpoint. Our simulated data (see Methods) follows this model.

Detecting an anomaly after intervention is therefore not as simple as identifying a statistically robust signal of change. To improve classification of anomalies into their root causes, we apply CLARITY, a non-parametric multivariate comparison method designed to identify correlations that were not already found in the pre-intervention state. For example, doubling all metabolite levels would have no effect on test statistics derived from CLARITY; neither would doubling metabolite levels in any major clusters. Rather, doubling only a new set of metabolites would change the covariance structure in the data and therefore, be picked up by CLARITY. Here, CLARITY is used alongside a traditional linear mixed model (LMM) approach for identifying differences in means with a view to assessing the potential value added by the CLARITY analysis.

To test the interpretation of the joint analysis of the pre-intervention (‘baseline’) and post-intervention (‘endpoint’) state of metabolites, we introduce a simulation study that captures the most important features – also following the framework described in **Figure 1** - and for which the ground truth is known.

### Simulated metabolite data

To compare which anomalies the LMM and CLARITY approaches capture, we performed a simulation designed to represent a typical NMR-derived metabolite dataset with similar properties to our experimental data set. In this we simulated metabolites to represent shared causal pathways from covariates, whereby metabolites belong to clusters which are jointly activated by those covariates in a hierarchical model. Specifically, each covariate activates two clusters, each differently, and each metabolite differently within a cluster. Anomalies were then planted in the ‘endpoint’ observation that affect either a whole cluster or only half of a cluster.

Data was generated for *n* subjects and *M* metabolites, using an idealised notion of “pathways” - here defined as a set of metabolites that covary together – from observed confounders (*C*) which are included in the linear analysis and unobserved confounders (*U*) which are treated as unknown. We wish to detect an intervention “pathway” *C*^*^ for which coefficients are non-zero only in the endpoint. Then metabolite *M*_*j*_, at time t is given by:

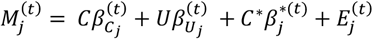

where the errors *E*^(1)^, *E*^(2)^ are generated independently according to a standard normal distribution. The important component of the model is the correlated effect sizes 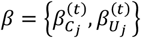, and the intervention pathway effects 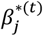.

Each metabolite is assigned to one of *c* evenly sized clusters, which informs how the *β*s are chosen to create a covariance structure representing critical features of clinical data. The metabolite data from both time points can be thought of as being generated by a multi-level hierarchical model which defines the covariance of the effect sizes *β*.

Specifically, let Σ^*l*^ define the covariance of all *β*s within a time point for metabolite *l*, for which we choose two clusters at random to be “activated”. 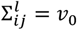 if *i* or *j* are outside of the activated clusters, *v*_1_ if they are in different activated clusters, *v*_2_ for the same activated cluster and *v*_3_ if *i* = *j* in an activated cluster. By choosing *v*_0_ < *v*_1_ < *v*_2_ < *v*_3_, the covariance is positive definite.

The joint effect sizes for each covariate across both time points are generated from a joint distribution Σ^*L*^, with correlation *ρ*_*t*_ across time, as:

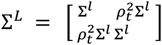

Finally, the intervention pathway 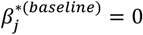, whilst a subset of metabolites {*M*_j_} has 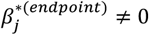 within two randomly selected clusters. For the first, say *c*_1_, the effects 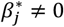 for the whole cluster, which we interpret as strengthening an existing pathway. In the case of the second, a random half of the metabolites denoted *c*_2_ has 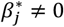, which we interpret as a new pathway. Further, we denote the remaining half *c*_3_, which has a second-order change as its relationship with similar metabolites has changed. 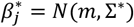 consists of a mean change *m* and a covariance increase parameterised as above.

Full details of how to generate *β*_*C*_, *β*_*U*_, *β*^*^ and *C*^*^ are provided in the **Supplementary Data 1**.

### Experimental data

Metabolomics data derived from samples collected during the By-Band-Sleeve (BBS) clinical trial were used. BBS is a pragmatic randomised controlled trial (RCT) which took place in the UK (registration number: ISRCTN00786323). The trial investigated the clinical and cost-effectiveness of three types of bariatric surgery: the Roux-en-Y gastric bypass (“Bypass”), laparoscopic adjustable gastric band (“Band”) and the sleeve gastrectomy (“Sleeve”). A full description of the trial can be found in published works; specifically, the trial protocol^15^, a description of the baseline data^16^ and the primary results^17^. In this study, serum samples that were collected at baseline (prior to surgery) and at 36-months after randomization were used to generate metabolomics data. The median (interquartile range) of time between baseline (randomization) and surgery was 2.7 (1.9, 4.5) months, and between surgery and 36-month follow-up was 33.4 (31.7, 34.8) months. Details of the sample collection and processing procedures have been previously described^18^. Included here are data from a pilot subset of 125 patients who were recruited at a single hospital site and had serum samples available for analysis from both baseline and 36-months post-randomisation by June 2021. The 250 samples were sent for NMR metabolomics analysis by Nightingale Health (Helsinki, Finland).

A high-throughput ^1^H-NMR metabolomics platform was used to quantify 250 metabolic biomarkers, 160 in absolute levels and 90 derived (ratio/percentage/total) measures. Details of experimental procedures have been described elsewhere^19,20^. Data were received in October 2022. The biomarkers include detailed measures of cholesterol metabolism, fatty acid compositions and various low-molecular weight metabolites, such as amino acids, ketones and glycolysis metabolites. The 14 lipoprotein subclass sizes are defined as follows: extremely large VLDL with particle diameters from 75 nm upwards and a possible contribution of chylomicrons, five VLDL subclasses (average particle diameters of 64.0 nm, 53.6 nm, 44.5 nm, 36.8 nm, and 31.3 nm), IDL (28.6 nm), three LDL subclasses (25.5 nm, 23.0 nm, and 18.7 nm) and four HDL subclasses (14.3 nm, 12.1 nm, 10.9 nm, and 8.7 nm). The following lipid components within the lipoprotein subclasses are quantified: phospholipids (PL), triglycerides (TG), cholesterol (C), free cholesterol (FC), and cholesteryl esters (CE). The mean size for VLDL, LDL and HDL particles is calculated by weighting the corresponding subclass diameters with their particle concentrations^19^.

The R package *metaboprep*^21^ was used to perform pre-processing and quality control (QC) of the NMR data. Any samples and features with extreme missingness (>80%) were excluded before recalculating sample and feature missingness. Then samples with more than 20% missingness and features with more than 20% missingness were removed from the dataset. PCA analysis was also performed and any samples that were more than five SDs from the mean of the first and/or second principal components were filtered from the data set. No imputation of the data was performed. A summary of dataset QC can be found in **Supplementary Data 2**. After QC filtering, 245 samples and 250 metabolites remained in the dataset. Finally, data for individuals who did not have surgery before the 36-mth post-randomization sample collection timepoint, were excluded from analyses. This left data for 237 samples from 121 unique individuals. Metabolite data were restricted to the 160 absolute measures (i.e., derived measures were excluded). Because the metabolomics data are measured on widely variable scales, to give equal weight in the algorithm they were jointly mean centred and scaled to have standard deviation (SD) 1 using the base R function *scale()*.

### Statistical analysis

#### Linear mixed model to detect mean differences

A linear mixed model (LMM) was implemented using the *lmer()* function from the *lmerTest* R package^22^ to identify metabolite levels whose mean differed between the two time points represented in the data.

The following formula was used:

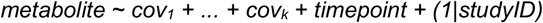

where for the simulated data *k=4* and cov_1_, …, cov_k_ were continuous normally distributed random variables. For the experimental data, covariates were *storage*.*time + age + sex*, where *storage*.*time* is the estimated length of time in months between sample collection and aliquoting for metabolomics analysis, *age* is participant age at baseline in years, *sex* is a binary variable (self-reported sex, male/female). *‘studyID’* is the identifier for each participant, fitted as a random effect to account for the repeated measures, and ‘*timepoint*’ is the independent (or predictor) variable. Model betas for ‘*timepoint*’ represent the estimated difference in metabolite abundance (in SD units) at the second time point (post-treatment) as compared to the first (pre-treatment). Benjamini-Hochberg (BH) corrected p-value^23^ was calculated using the “BH” method in the *p*.*adjust()* function from the ‘stats’ R package^24^.

#### CLARITY analysis to detect structural differences

CLARITY is a method to detect differences in structure – meaning components analogous to clusters or shared variance components - between two datasets. If we knew the causal pathways, we would regress these out to identify the changes, but these are very difficult to learn. CLARITY solves this problem by building a representation of the similarity between metabolites at baseline, which is used to predict the similarity at endpoint. In this way, change in the relative importance of metabolite groupings that are present in the baseline will not be identified, but metabolite groupings present in the second dataset alone are reported as anomalies. CLARITY uses a non-parametric approach in which increasingly complex representations of the first dataset are used to predict the second. An “anomaly” is therefore defined as a structure present in the second dataset and not in the first, that “persists” across a wide range of model complexities. In the current application, we reframe the CLARITY approach such that the shared entities are common omics – in this case a set of measured metabolites, rather than individuals or samples. The same metabolites have been measured at two different sampling timepoints.

All functions mentioned in this section form part of the R CLARITY package^14^. CLARITY was applied to the simulated and clinical datasets as follows. For this multivariate analysis, we are given feature matrices, denoted as *D*_1_ (the reference data) and *D*_2_ (the target data) each with *L* rows. From these, column-wise covariance matrices of the feature matrices of metabolites are taken, denoted as *Y*_1_and *Y*_2_. Note that this removes the requirement that the datasets contain the same individuals.

The first step is creating a rank *k* approximation of *Y*_1_ with the *Clarity_Scan()* function:

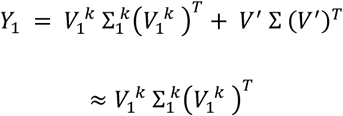

*Y*_1_ and *Y*_2_ are predicted using this rank *k* representation of the baseline *Y*_1_, implemented in *Clarity_Predict()*, giving:

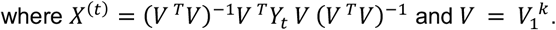

*Clarity_Persistence()* is then used to compute a test statistic, simply the squared row sums of residuals:

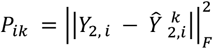

This test statistic is named “persistence” because the set of metabolites *i* that have large residuals remains highly consistent for a wide range of model complexities *k* by construction. Conceptually, the singular value decomposition (SVD) of the baseline data is learning structure (i.e. cluster-like objects) for small *k*, and noise (or structures that are not in the endpoint) as *k* grows. At very large *k* this allows overfitting to the endpoint data, so an appropriate choice of *k* is from the wide region where anomalies persist.

A cross-validation procedure for computing p-values for each metabolite is implemented in *Clarity_Compare()* and described below. Procedure:

1. For *r* = 1, …, *n*_*obs*_:
  a. Randomly split the features of *D*_1_ to create 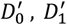.
  b. Sample *L*/2 rows from *D*_2_, call this 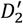.
  c. Create covariance matrices *Y*_0_, *Y*_1_, *Y*_2_ from 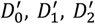.
  d. Perform CLARITY on pairs (*Y*_0_, *Y*_1_) and (*Y*_0_, *Y*_2_) with rank *k*.
  e. Compute test statistic f(persistences) for each row of *Y*_1_ and *Y*_2_.
2. Compute the final test statistic for each row:

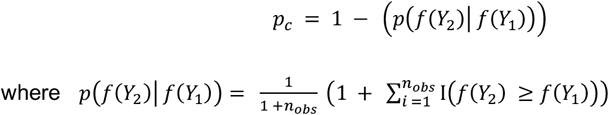

For the simulated data, the p-values were taken, with *k* to be the number of simulated clusters. In the analysis of the experimental data (where *k* is unknown), persistence values were computed for all *k*. As claimed from the name, it is observed that an almost identical set of anomalous metabolites “persist” i.e. have extreme test-statistic values, for a wide range of *k* from 10 to 40. We therefore discuss and interpret only anomalies that persisted over this range of *k*, whilst corresponding test statistics (i.e., p-values) are reported for rank *k=25*.

#### Ethical approval

This study was conducted in accordance with the principles of the Declaration of Helsinki. Ethical approval for BBS was granted by the Southwest Frenchay Research Ethics Committee (reference 11/SW/0248) and written informed consent was obtained from all participants. All samples were used, stored and disposed of in accordance with the Human Tissue Act 2004.

## Results

### Simulated data

We consider how each method performs in identifying multiple types of planted changes. We use the simulation framework described above with four observed and two unobserved covariates.

There were seven clusters of metabolites with correlation across time points *ρ* = 0.8, the minimum covariance was *v*_1_ = 0.0375, the within-activated cluster covariance was *v*_3_ = 0.12, and the between-activated cluster covariance *v*_2_ = (*v*_1_ + *v*_3_)/2. We explore the case with a large intervention pathway *C*^*^ mean change of *m=*0.5.

Figure 2. shows the covariance matrices and how well CLARITY predicts their structure when we set its complexity parameter *k* to be seven, the number of simulated clusters. The residual matrix for the simulated baseline is simply noise, suggesting this representation is complex enough to capture the structure of the baseline data. In the endpoint data, it is clear to see that there is still structure in the residual matrix. For these parameters, the persistence is highest for metabolites in clusters *c*_2_ and *c*_3_, with *c*_1_ also showing smaller but still substantial persistences.

While metabolites don’t fall into neatly defined groups, we can see that there are broad trends in where differently coloured points fall in **Figure 3**. We can see that the metabolites in *c*_1_and *c*_2_, which have experienced a first order change, show a consistently stronger signal in the univariate analysis. In contrast, CLARITY rates the change in *c*_2_ and *c*_3_ to be more significant on average, as the structure of that block has changed from one uniform cluster to two clusters within a larger block. Even though *c*_3_ was not directly affected, it’s relationship with *c*_2_ causes it to experience a second-order effect which can now be detected and classified.

**Figure 2.**
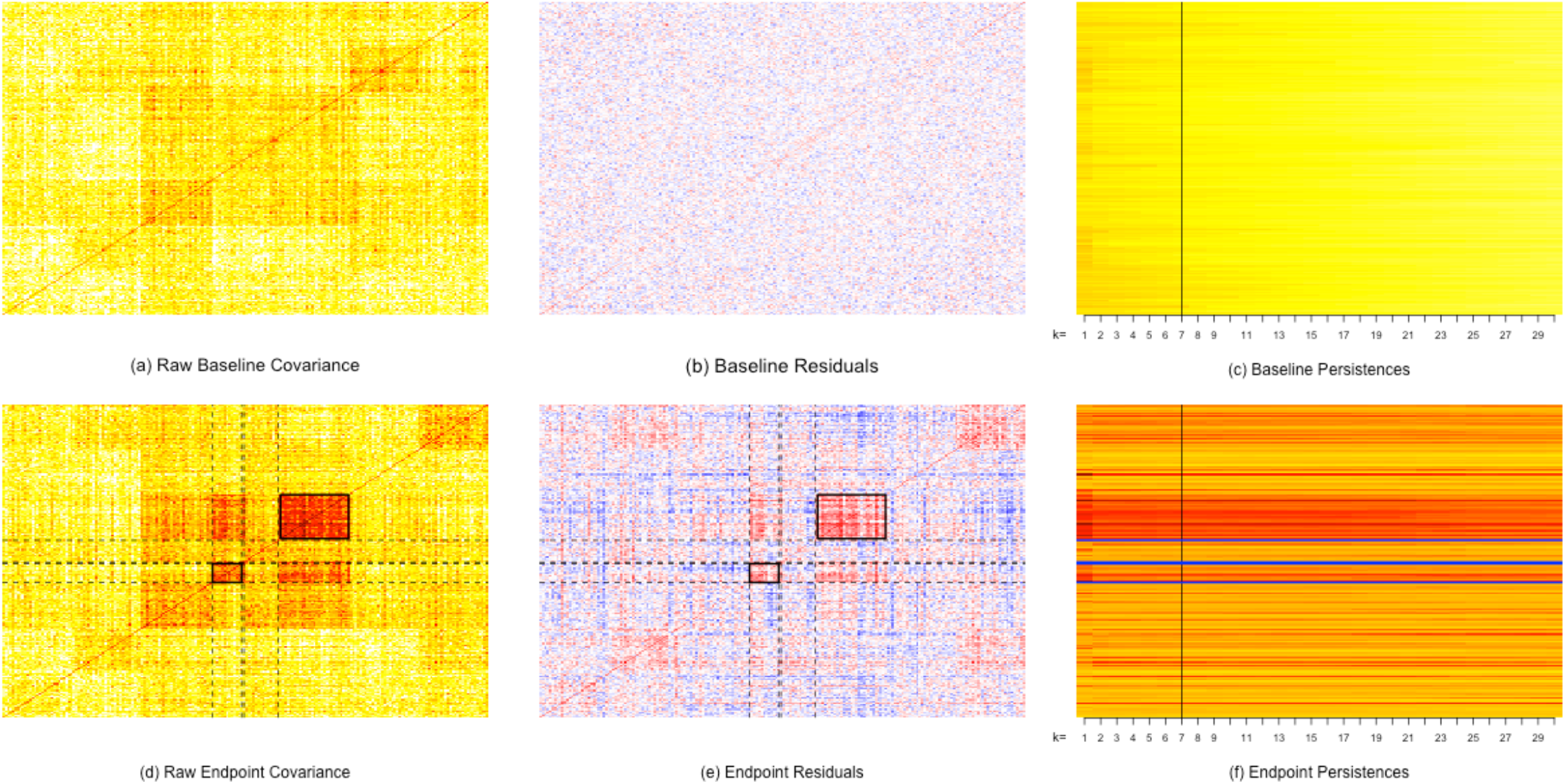
CLARITY plots for simulated data. A representation of the simulated data as it moves through the CLARITY pipeline. The first (a-c) and second (d-f) rows correspond to the baseline and endpoint data respectively. The first column (plots (a) and (d)) shows the raw covariance matrices. The second column (plots (b) and (e)) shows the residuals of the CLARITY model at k = 7. The third column (plots (c) and (f)) shows the persistences from k = 1, …, 30, with a black line showing the persistences for k = 7. In the endpoint plots, dashed lines and rectangles indicate the clusters that have a planted change. Scale: red: large and positive, decreasing through yellow and white=zero, to blue: large and negative.

**Figure 3.**
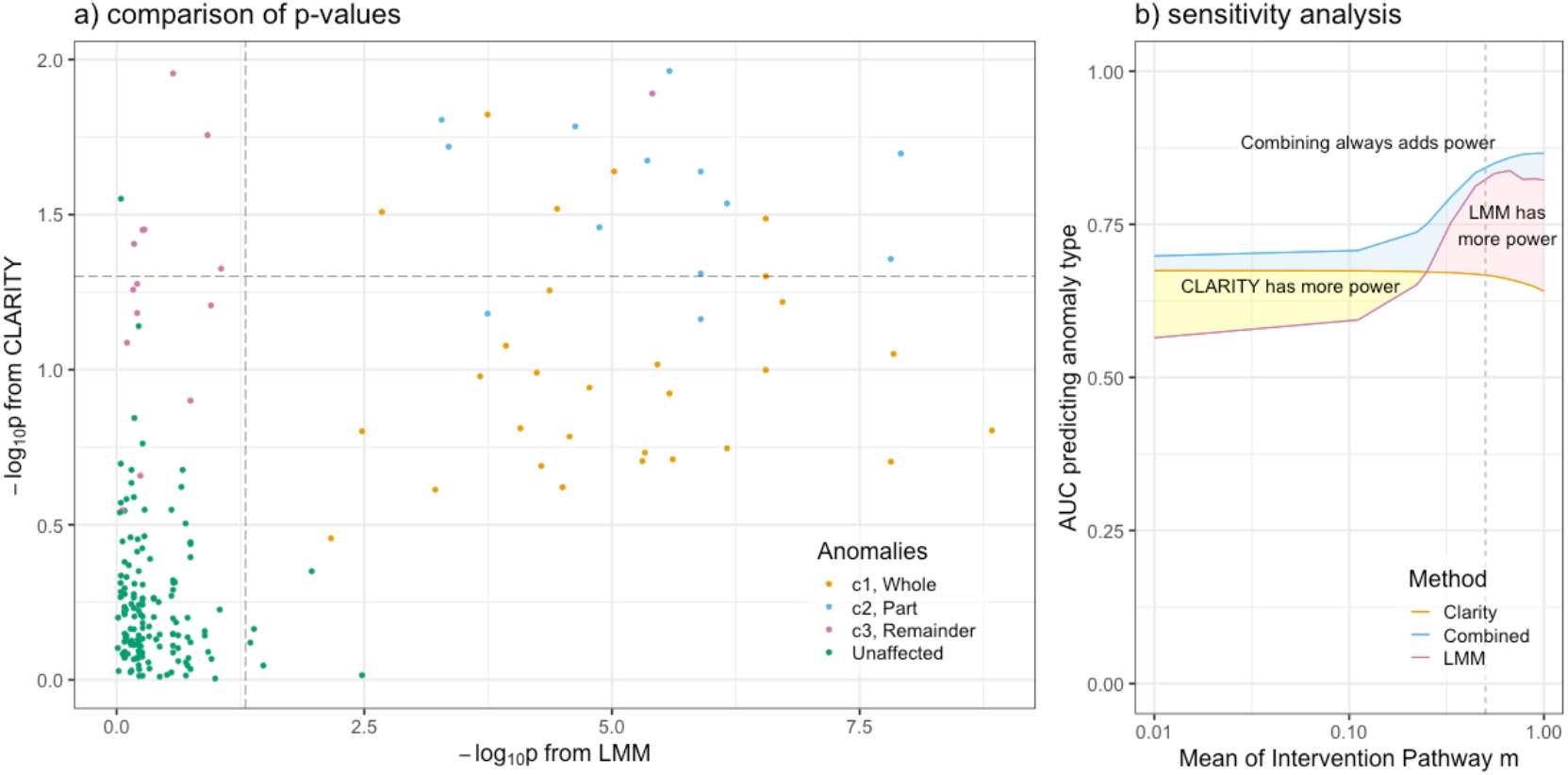
Linear mixed model (LMM) and CLARITY associations in simulated data. a) Association statistics from the two methods, with metabolites coloured to reflect the type of change they experienced. Grey dashed lines have been included to indicate p = 0.05. LMM = linear mixed model. b) Performance metric recovering the exact anomaly class type as measured by Area under the ROC curve (AUC), using a test statistic of the CLARITY persistence, LMM p-value, or Combined, as a function of intervention mean m. Grey dashed line at 0.5 shows the parameters from (a).

### Experimental data

Of the 160 NMR metabolites tested in the LMM, 30 (19%) were altered by the intervention (BH-corrected p<0.05) with 25 of those having higher levels at 36-months post-randomisation as compared to baseline (**Supplementary Table 1**). The strongest association was seen for triglycerides in large HDL (beta=0.78 SD, 95% CI: 0.44, 1.12, BH-adjusted p=0.002) with three other metabolites having BH-adjusted p=0.003 (average diameter of HDL particles (beta=0.69 SD, 95% CI: 0.36, 1.02), valine (beta=-0.75 SD, 95% CI: −1.11, −0.39) and isoleucine (beta=-0.71 SD, 95% CI: −1.06, −0.37)). There were relatively more metabolites from the ‘branched chain amino acid (BCAA)’ class in the LMM-associated subset as compared to all metabolites tested (13.3% compared to 5.6%) (**Figure 4**).

**Figure 4.**
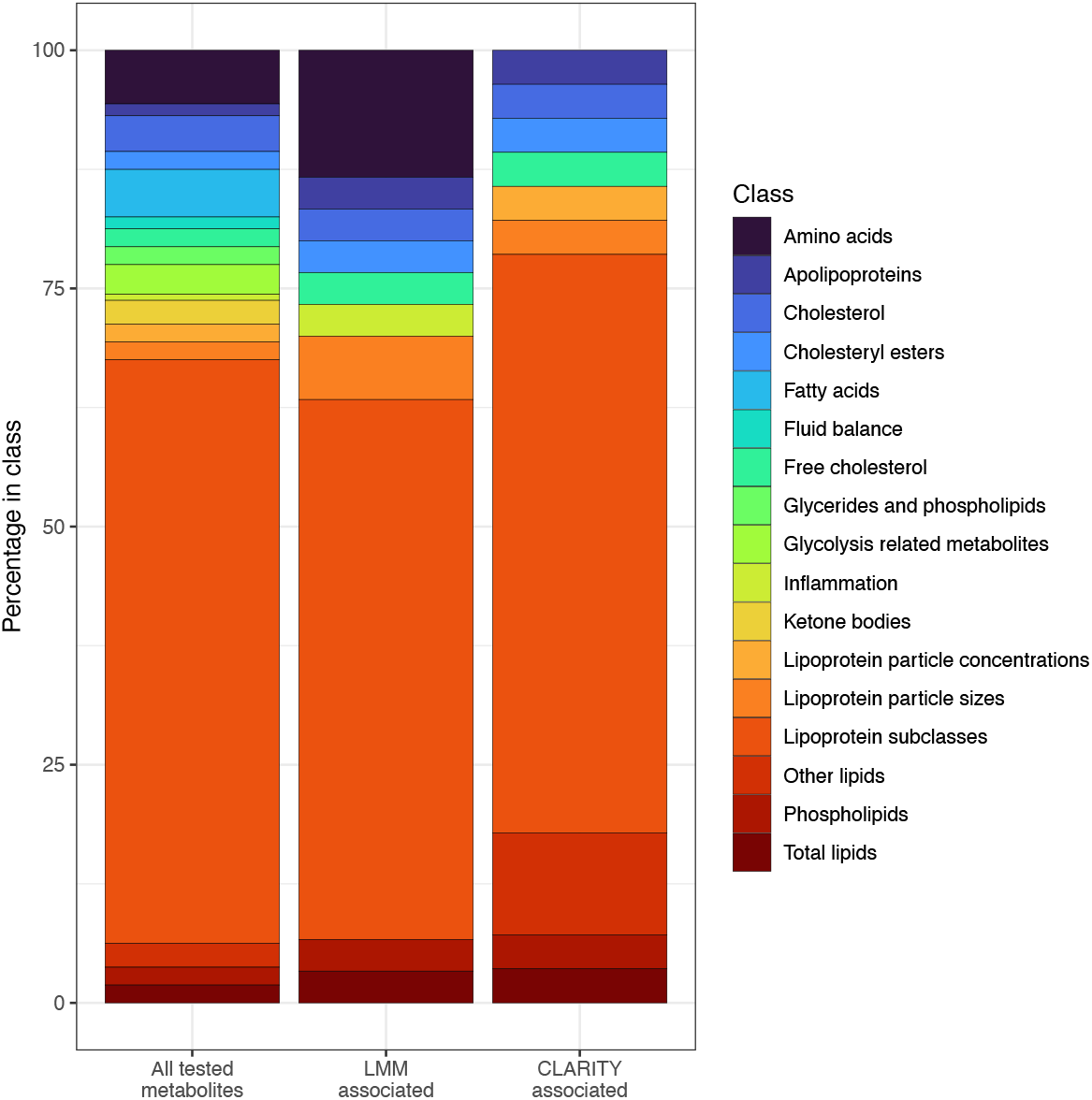
Distribution across classes of intervention-associated metabolites identified by the two different methods, as compared to all metabolites tested. LMM = linear mixed model.

In the CLARITY analysis, 28 metabolites (18%) had an empirically derived persistence p<0.05 (with k=25) suggesting that they could not be well-predicted at the 36-month timepoint based on baseline (reference) data structure, even in models with a relatively high degree of complexity (**Figure 5**). The five metabolites with the greatest persistence were Concentration of large HDL particles, Total cholesterol in large HDL, Cholesterol esters in large HDL, Total lipids in large HDL, and Phospoholipids in large HDL (**Supplementary Table 1**). There were relatively more metabolites from the ‘other lipids’ class (10.7% compared to 2.5%) in the subset with persistence p<0.05 as compared to all metabolites tested (**Figure 4**). No metabolites from the amino acids class had persistence p<0.05 in CLARITY. The distribution of pairwise correlations across metabolites differed (**Supplementary Data 3 & 4)** with those metabolites from the amino acid class typically showing a normal distribution centred on zero and those from the lipid classes frequently exhibiting bimodal distributions indicating moderate positive and negative correlations with many other metabolites. Visualising the between metabolite correlations at the two timepoints helps to elucidate the changes that have occurred in the case of metabolites that returned high persistence values (**Figure 6a & b**), whereas for metabolites that did not demonstrate persistence there is no obvious change in correlation structure (**Figure 6c & d**).

**Figure 5.**
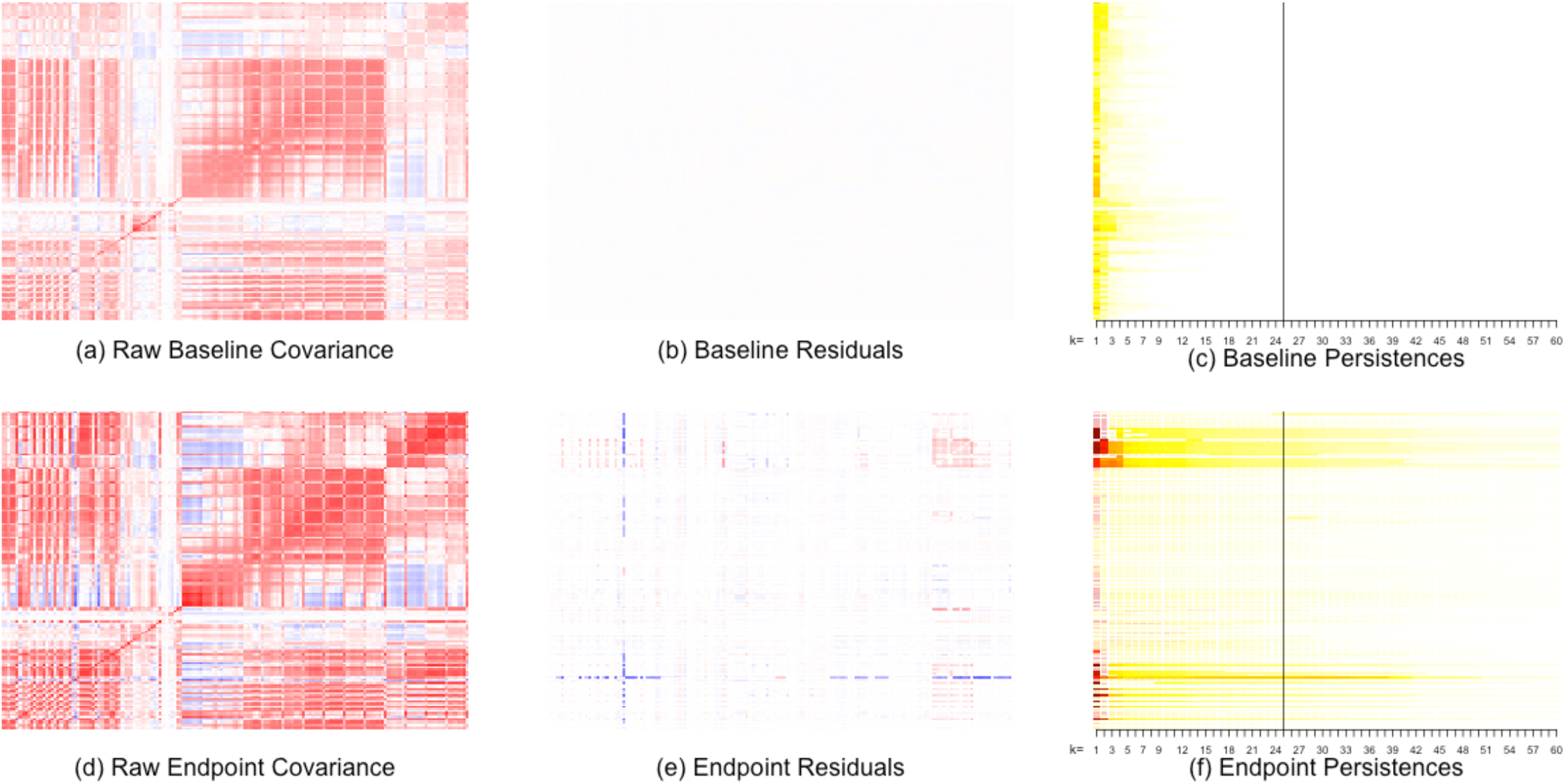
CLARITY plots for experimental data. A representation of the clinical data as it moves through the CLARITY pipeline. The first (a-c) and second (d-f) rows correspond to the baseline and endpoint data respectively. The first column (plots (a) and (d)) shows the raw covariance matrices. The second column (plots (b) and (e)) shows the residuals of the CLARITY model at k = 25 on the same scale, “fading” (making transparent) non-significant residuals. The third column (plots (c) and (f)) shows the persistences from k = 1, …, 60 on the same scale, with a black line indicating the persistences for k = 25 and “fading” p-values > 0.05. Scale: red: large and positive, decreasing through yellow and white=zero, to blue: large and negative.

**Figure 6.**
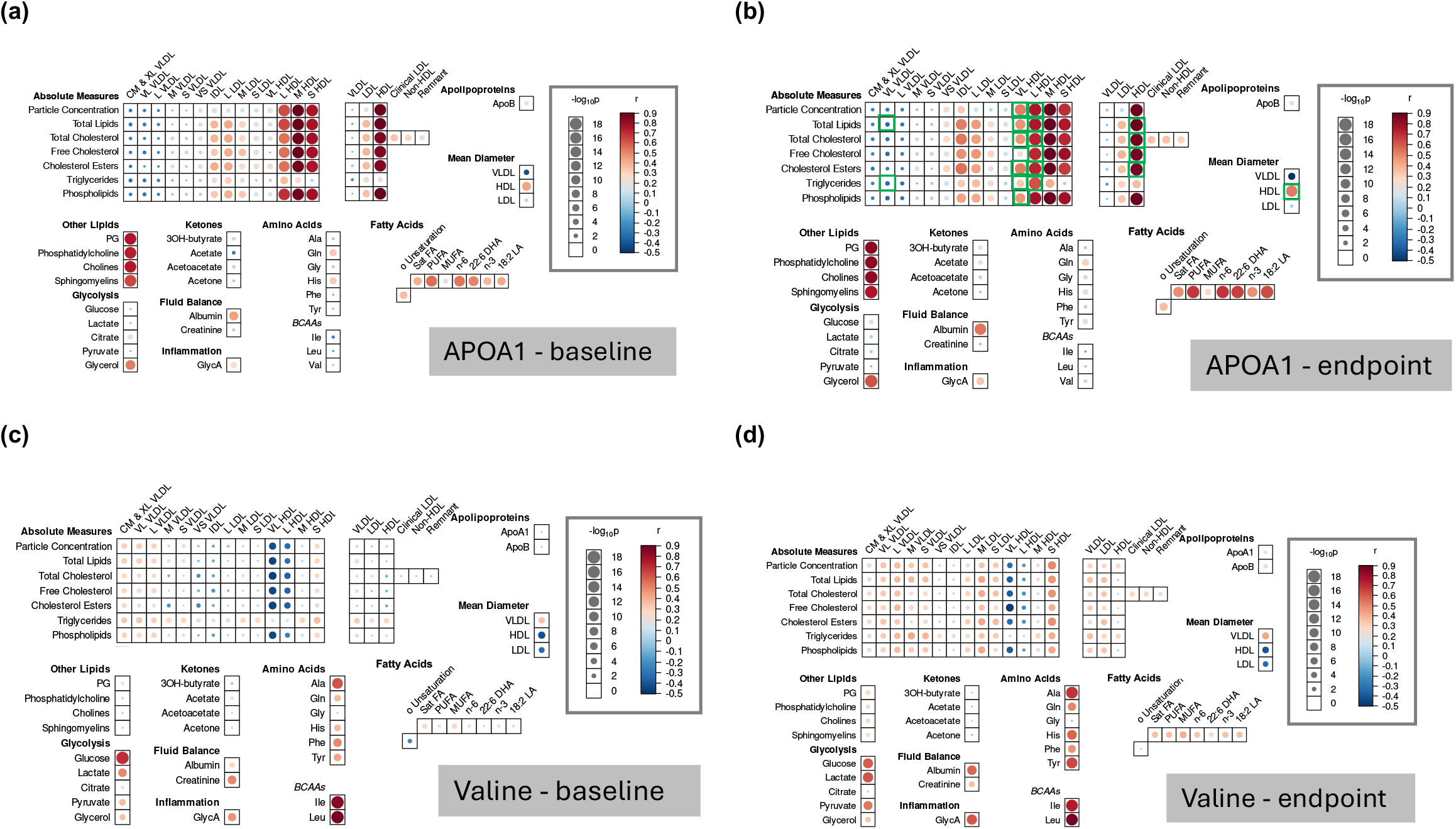
Correlation heat maps showing the relationship between APOA1 (a & b) and valine (c & d) and all other measured metabolites firstly at baseline (a & c) and then at endpoint (b & d). APOA1 showed persistence in the CLARITY analysis and green squares highlight which APOA1-metabolite relationships changed (p<0.05). Valine did not show persistence in the CLARITY analysis and no meaningful differences in valine-metabolite relationships were identified. Plots produced using a modified version of the R package ‘bubbleHeatmap’^25^. r = Pearson correlation; −log_10_p = minus log_10_ of p-value from test of Pearson’s correlation.

To check for simple explanations for our results, we computed the total correlation (sum of absolute pairwise correlations, *r*, values) for each metabolite first at baseline and then again at endpoint. Examining the difference, the metabolites with persistence p<0.05 exhibited differences in overall correlation across the full range (decrease, increase and no difference) (**Figure 7**), i.e. our findings cannot be explained by changes in the metabolite’s total correlation from baseline to endpoint. It is critical to consider the persistence value in the context of an empirically derived test statistic. Whilst a small number of metabolites returned high persistence values, these were found not to be statistically meaningful when put in the context of the empirically derived p-value (**Supplementary Data 5, top panel)**, likely because they were simply very variable between time points. There was additionally no strong relationship between total correlation at baseline and persistence (**Supplementary Data 5, middle and bottom panel)**. From these sanity checks we conclude that the anomalies indeed represent a change in the covariance between metabolites.

**Figure 7.**
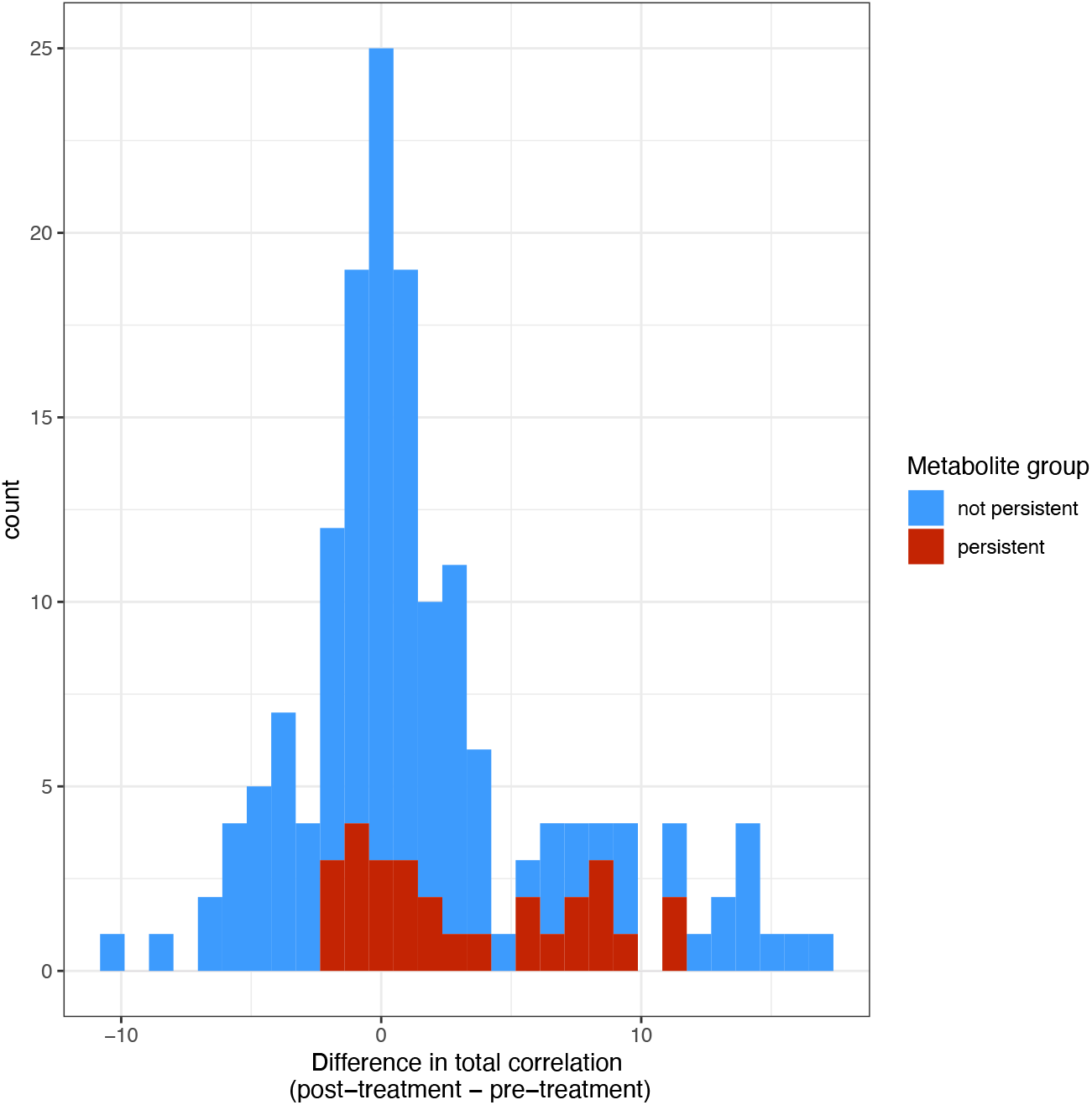
Distribution of change in total correlation for each metabolite (post-treatment – pre-treatment). Coloured according to whether metabolites showed persistence in CLARITY or not. A comparison of the two approaches shows a moderate degree of overlap. While some metabolites have an approximately proportional signal in the LMM and CLARITY analyses (based on respective p-values), others are only identified using one of the approaches (**Figure 8**). For example, the branched chain amino acids (BCAA) have a strong signal in the LMM analysis but do not show meaningful persistence in the CLARITY analysis. Extra-large HDL molecules predominate in the list of metabolites identified more strongly in the CLARITY analysis. And amongst those shown to differ in both approaches are several medium and large HDL molecules, as well as HDL particle size and APOA1 (**Figure 6a & b**).

**Figure 8.**
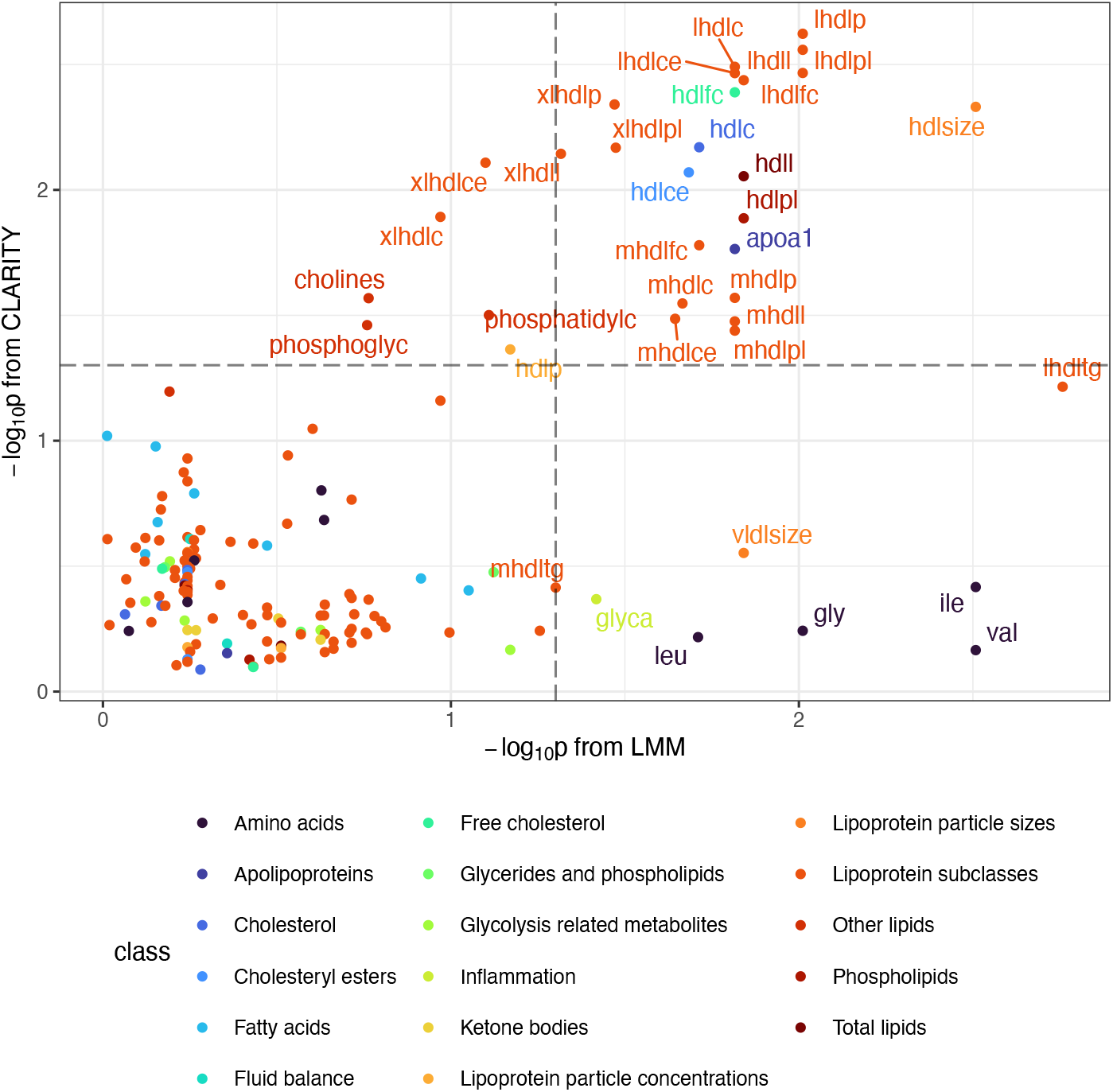
Association statistics from the two methods, linear mixed model (LMM) and CLARITY, applied to experimental data. Metabolites have been coloured according to their class. Grey dashed lines have been included to indicate p = 0.05 (BH-adjusted for LMM). Metabolites with p<0.05 in one or both analyses have been labelled (full names provided in Supplementary Table 1). LMM = linear mixed model. For a more detailed exploration of the pre- and post-intervention within metabolite relationships for APOA1 and valine (val), see Figure 6.

## Discussion

Metabolite profiles vary after an intervention for a variety of reasons. Many changes are technical as it is impossible to keep everything constant between observations of metabolites that vary due to many exogenous factors, including time of measurement, time since consumption of food, exercise, sleep and so on. These unobserved factors will be present at both time points but are expected to vary in relative impact, resulting in confounding for methods that look at before and after patterns of a single metabolite for each person.

The changes of interest in any given association analysis of longitudinal change are complex and involve a systematic change to the underlying (and largely unknown) metabolic pathways, therefore affecting many metabolites simultaneously. Information about these pathways is encoded in the co-occurrence patterns of metabolite profiles. We have shown that it is possible to identify sets of anomalous metabolites with new covariance structures after an intervention. Under our simulation model, this would be interpreted as the activation of a “metabolic pathway” in the sense of a group of metabolites responding together under this treatment condition. This was detected using CLARITY, a non-parametric analytical approach robust to noise and focused on evaluating the relative conservation of (dis)similarities.

We have shown using simulated data that if there were truly multiple metabolic pathways detectable as covarying metabolites and these varied in importance between baseline and endpoint, the CLARITY approach can correct for this and identify sets of metabolites involved in a pathway specific to the endpoint (here post-intervention) dataset – i.e. that changed because of intervention. Conversely, traditional LMM approaches based on independent regressions identify a combination of covariance changes, mean changes, and confounding by technical changes. The metric used by CLARITY to quantify the degree of novelty (or unpredictability) of metabolites in the endpoint is a ‘persistence’ score. By considering the persistence score (and its test statistic) alongside results from traditional univariate analyses we can gain information regarding how (not just if) metabolites are changing post-intervention making this a complementary analysis to LMM.

In the CLARITY analysis of our experimental dataset, we observed meaningful persistence for 28 metabolites in total, six of which did not exhibit a strong effect in the LMM analysis (i.e., did not differ in their mean between baseline and endpoint based on a BH-adjusted *p* > 0.05) including HDL particle concentration, cholines and phosphoglycerides. The sets of anomalous metabolites identified partition quite neatly into two classes, which we can interpret using the simulation analysis and by understanding the types of anomalies each method tries to identify. Firstly, metabolite anomalies identified by both methods (which CLARITY has more power to detect) are suspected of being involved in a pathway active only in the post-intervention state. Secondly, metabolite anomalies identified by the LMM only are likely to belong to changes in pathways active in both timepoints. LMM approaches robustly detect mean changes which CLARITY is not designed to detect, but follow-up (experimental or otherwise) is needed to assess which type of change is most likely to lead to actionable insights. What is clear is the ability to annotate metabolites by whether they appear in a novel pathway compared to a baseline, is informative of the underlying biology and likely to be an important tool in the analysis of massively multivariate ‘omics data.

Our contribution is primarily methodological, and specific results should be seen as exploratory. We found that branched chain amino acids (BCAAs) (leucine, isoleucine, valine) showed a strong effect in the LMM but little persistence in CLARITY suggesting an effect of the intervention on mean levels via a pre-existing pathway, but no new relationship with other measured metabolites; indeed, this change in mean is supported by existing literature^26^. In general, and across both timepoints, the BCAAs exhibit a strong positive correlation with each other, a moderate positive correlation with other amino acids but a low correlation with most of the lipid metabolites that dominate our dataset (see **Figure 6c & d** for valine as an example). To investigate whether the low overall correlation of the BCAAs with other measured metabolites was the reason they did not exhibit high persistence, we considered the relationship between total correlation and persistence. There was little indication that an overall low (or high) level of relatedness with other metabolites unduly influenced the CLARITY results. This suggests the cross-validation procedure used to generate empirical test statistics effectively accounts for such features in the data.

Metabolites identified with greater power in the CLARITY analysis were all lipids or lipid components with a predominance of very large HDL molecules. It is important to note that these metabolites fall into a continuum that includes metabolites that were identified by both methods, therefore showing both differences in mean and differences in their relationship with other metabolites. These included APOA1 (**Figure 6a & b**), the principal protein component of HDL, as well as several medium and large HDL molecules. The identification of HDL molecules by CLARITY is in keeping with recent literature that points towards the relevance of HDL quality and functionality (determined by properties such as the composition of lipid and protein, and particle shape and number) and not just quantity to health^27^. Investigating this group of molecules further is an important activity for future work.

CLARITY is an exploratory data analysis tool for approaching high-dimensional statistical comparisons and so is limited for drawing formal conclusions. Because it corrects for structure learned from the baseline dataset using arbitrary linear combinations in the test dataset, it has limited power to detect and quantify changes in factors already present. There are many situations we would expect these to change meaningfully despite the issues around confounding that are expected, particularly for metabolites. Methods to quantify this class of change would be welcomed, for example Unfolded Spectral Embedding^28^ provides embeddings in which distances between multivariate embeddings (before vs after) are meaningful, and which can be extended to similarities^29^. Other limitations include a lack of theoretical guidance for how to pre-process the data, which is currently an implicit part of choosing a similarity measure.

There is a need to develop a wide range of tools that can detect different types of structural change in ‘omics data, which should ultimately lead to the ability to identify specific biological pathways impacted by an intervention. The CLARITY approach we have explored here identifies structural changes in metabolomic covariances that provides complementary information to traditional regression. It has the advantage of requiring no training data and because it uses covariances only, could be applied even if the two datasets were observed on different individuals. Next steps include comparison to a “healthy normal”, comparison of different treatments, and baseline validation where experimental work is available to probe the actual pathways activated.

## Data Availability

At the time these data were generated, participants were not asked for their permission to share data beyond the immediate project team. However, anonymised individual patient data from the By-Band-Sleeve trial will be made available upon request to the chief investigator (Jane Blazeby) for secondary research, conditional on assurance from the secondary researcher that the proposed use of the data is compliant with the Medical Research Council Policy on Data Sharing regarding scientific quality, ethical requirements, and value for money, and is compliant with the National Institute for Health and Care Research policy on data sharing. A minimum requirement with respect to scientific quality will be a publicly available prespecified protocol describing the purpose, methods, and analysis of the secondary research (e.g., a protocol for a Cochrane systematic review), approved by a UK Research Ethics Committee or other similar, approved ethics review body. Participant identifiers will not be passed on to any third party.

## Supplementary Data Files

*Supplementary Data 1:* Extended methods – simulation pipeline

*Supplementary Data 2:* Extended results – *metaboprep* data preparation summary report

*Supplementary Data 3:* Extended results – distribution of metabolite correlations at baseline

*Supplementary Data 4:* Extended results – distribution of metabolite correlations at endpoint

*Supplementary Data 5:* Extended results – exploration of CLARITY results.

Top figure: Plot of −log10(p) on persistence score (CLARITY)

Middle figure: Plot of −log10(p) from CLARITY on total metabolite correlation at baseline.

Bottom figure: Plot of persistence score from CLARITY on total metabolite correlation at baseline.

*Supplementary Table 1:* Full results from linear mixed model and CLARITY for all metabolites tested.

## Acknowledgments

The By-Band-Sleeve trial is led by JM Blazeby (Chief Investigator) and CA Rogers (lead methodologist). We acknowledge the support of all other members of the NIHR By-Band-Sleeve Management Group: Robert C Andrews, John Bessant, James P Byrne, Nicholas Carter, Caroline Clay, Jenny L Donovan, Eleanor Gidman, Graziella Mazza, Mary O’Kane, Barnaby C Reeves, Nicki Salter, Janice L Tompson, Richard Welbourn and Sarah Wordsworth. We also thank all the By-Band-Sleeve contributors including the investigators, research dieticians and nurses, the independent trial steering committee and data monitoring and safety committee. We are grateful to all the patients who participated in this trial.

We thank the technical team at the Bristol Bioresource Laboratories at the University of Bristol for sample management once samples had been processed.

## Funding

RMW was supported by the EPSRC Centre for Doctoral Training in Computational Statistics and Data Science through a PhD studentship (Compass, EPSRC Grant number EP/S023569/1). NJT is the PI of the Avon Longitudinal Study of Parents and Children (MRC & WT MR/Z505924/1), is supported by the University of Bristol NIHR Biomedical Research Centre (BRC-1215-2001), the MRC Integrative Epidemiology Unit (MC_UU_00032/1) and works within the CRUK Integrative Cancer Epidemiology Programme (C18281/A29019). LJC is supported by the MRC Integrative Epidemiology Unit (MC_UU_00032/1). JMB is an NIHR Senior Investigator. CAR was funded by the British Heart Foundation until 2016.

By-Band-Sleeve was funded by National Institute of Health and Care Research (NIHR) Health Technology Assessment Programme (HTA 09/127/53). We also acknowledge funding from the MRC ConDuCT-II Hub for Trials Methodology Research and the NIHR Biomedical Research Centre (BRC) at the University of Bristol (IS-BRC-1215-20011). This trial was designed and delivered in collaboration with the Bristol Trials Centre, a UKCRC registered clinical trials unit (CTU), which is in receipt of NIHR CTU support funding. The views expressed are those of the author(s) and not necessarily those of the NIHR or the Department of Health and Social Care.

Metabolomics data generation in By-Band-Sleeve was supported by a Wellcome Trust Investigator Award held by NJT (202802/Z/16/Z).

This research was funded in whole, or in part, by the Wellcome Trust [202802/Z/16/Z, 218495/Z/19/Z]. For the purpose of Open Access, the author has applied a CC BY public copyright licence to any Author Accepted Manuscript version arising from this submission.

This work was carried out using the computational facilities of the Advanced Computing Research Centre, University of Bristol - “http://www.bristol.ac.uk/acrc/”.

## Author contributions

Conceptualisation: LJC, NJT, DJL; Data curation: LJC; Formal analysis: LJC, RJW, DJL; Funding acquisition: NJT, JMB, CAR; Resources: JMB, CAR; Supervision: NJT, DJL; Visualisation: LJC, RMW; Writing – original draft preparation: LJC, RMW, DJL; Writing – reviewing & editing: LJC, RMW, DJL, NJT, CAR, JMB.

