## Supplementary Data 1 for "Multivariate analysis of metabolomic data to identify biological pathways modified by a clinical intervention"

### Simulation Pipeline

Input:

- $n$: sample size
- $n_X$: number of metabolites
- $n_U$: number of unobserved confounders
- $n_C$: number of observed confounders
- $m$: mean effect
- $c$ : number of clusters
- $\sigma_c$: correlation of effects within a cluster
- $\sigma_x$: correlation of effects outside clusters
- $\rho_t^2$: shared covariance between effects across times

1. **Generate observed ($C$ ), unobserved ($U$) confounders and anomalous confounder**:
    1. Get vector of means $\mu \in \mathbb{R}^{n_C+n_U}$
    2. Set covariance matrix

    $$\Sigma_{ij}^C = \begin{cases} 1 & \text{if } i = j \\ 0.5 & \text{else} \end{cases}$$

    3. Obtain $n$ samples of covariates:

    $$UC_l \sim_{iid} \mathcal{N}(\mu, \Sigma^C) \qquad \text{for } l = 1, \ldots, (n_C + n_U)$$

    to get matrix $UC \in \mathbb{R}^{n \times (n_C+n_U)}$
    4. Generate variable $C^* \in \mathbb{R}^n$ which is only active in $t = 2$ for anomalous metabolites:

    $$C_i^* \sim \mathcal{N}(m, 1)$$

2. **Divide metabolites into evenly sized clusters**, denote cluster memberships as $Z_i \in \{1, \ldots, c\}$ for $i = 1, \ldots, n_X$.
3. **Create coefficient matrices** $\boldsymbol{\beta}^{(1)}, \boldsymbol{\beta}^{(2)} \in \mathbb{R}^{nX \times (nC+nU)}$ in a way such that each confounder activates a two clusters (mimicking pathways). For every $l = 1, \ldots, (n_U + n_U)$, we consider the coefficients corresponding to the $l^{\text{th}}$ column of $UC$:
    1. Sample 2 clusters to be activated, say $c_l^1, c_l^2$
    2. Set matrix $\Sigma_{ij}^{(l)}$ to be the marginal covariance matrix for coefficients within a time point as follows:

$$\mathbf{\Sigma}_{ij}^{(l)} = \begin{cases} \sigma_c & \text{if } Z_i = Z_j \in \left\{c_l^1, c_l^2\right\} \\ \frac{(\sigma_c + \sigma_x)}{2} & \text{if } Z_i \neq Z_j, \text{ and } Z_i, Z_j \in \left\{c_l^1, c_l^2\right\} \\ \sigma_x & \text{else} \end{cases}$$

i.e. $\sigma_c$ within an activated clusters, $\frac{\sigma_c + \sigma_x}{2}$ across activated clusters and $\sigma_c$ otherwise.

3. Create covariance matrix to sample $t = 1, 2$ coefficients jointly:

$$\mathbf{\Sigma}^L = \begin{bmatrix} \mathbf{\Sigma}^{(l)} & \rho_t^2 \mathbf{\Sigma}^{(l)} \\ \rho_t^2 \mathbf{\Sigma}^{(l)} & \mathbf{\Sigma}^{(l)} \end{bmatrix}$$

4. Draw $l^{\text{th}}$ column of coefficient matrices as follows

$$\left[\beta_l^{(1)} \; \beta_l^{(2)}\right] \sim \mathcal{N}\left(\mathbf{0}, \mathbf{\Sigma}^L\right)$$

4. **Create anomalies**:
1. Sample 2 clusters to be anomalies, say $c_1^A, c_2^A$ and set anomaly status vector $A$ to be $1$ for half of the metabolites in $c_1^A$ and all metabolites in $c_2^B$
2. Obtain coefficients for $C^*$ as $\boldsymbol{\beta}^* \in \mathbb{R}^{nX}$:

$$\boldsymbol{\beta}_i^* \sim \begin{cases} U[0.5, 1] & \text{if } A_i = 1, 2 \\ 0 & \text{else} \end{cases}$$

5. **Produce metabolite data** $X_1, X_2$:
   1. Generate error matrices $E^{(1)}, E^{(2)} \in \mathbb{R}^{n \times nX}$ according to a standard normal:

$$E_{ij}^{(t)} \sim \mathcal{N}(0, 1)$$

   2. For each metabolite $j$ at time $t$, get the vector $X_{t^j} \in \mathbb{R}^n$ of observed metabolites:

$$X_{t^j} = \begin{cases} C\boldsymbol{\beta}_j^{(t)} + E_j^{(t)} + C^*\boldsymbol{\beta}^* & \text{if } i \in A, t = 2 \\ C\boldsymbol{\beta}_j^{(t)} + E_j^{(t)} \end{cases}$$
