## Supplementary Data 3 for "Multivariate analysis of metabolomic data to identify biological pathways modified by a clinical intervention"

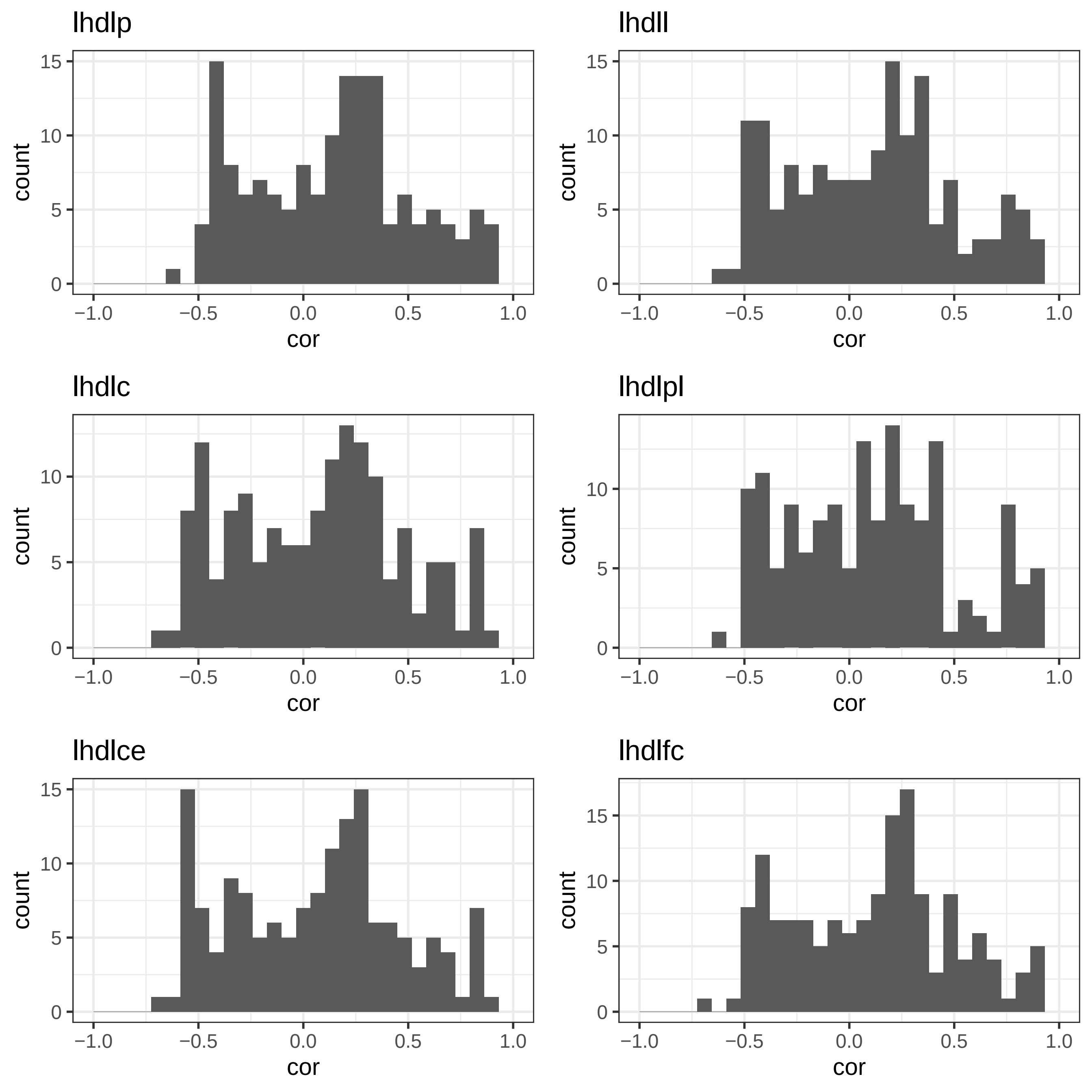
lhdlp
lhdll
lhdlc
lhdlpl
lhdlce
lhdlfc
count
cor
0
5
10
15
−1.0
−0.5
0.0
0.5
1.0

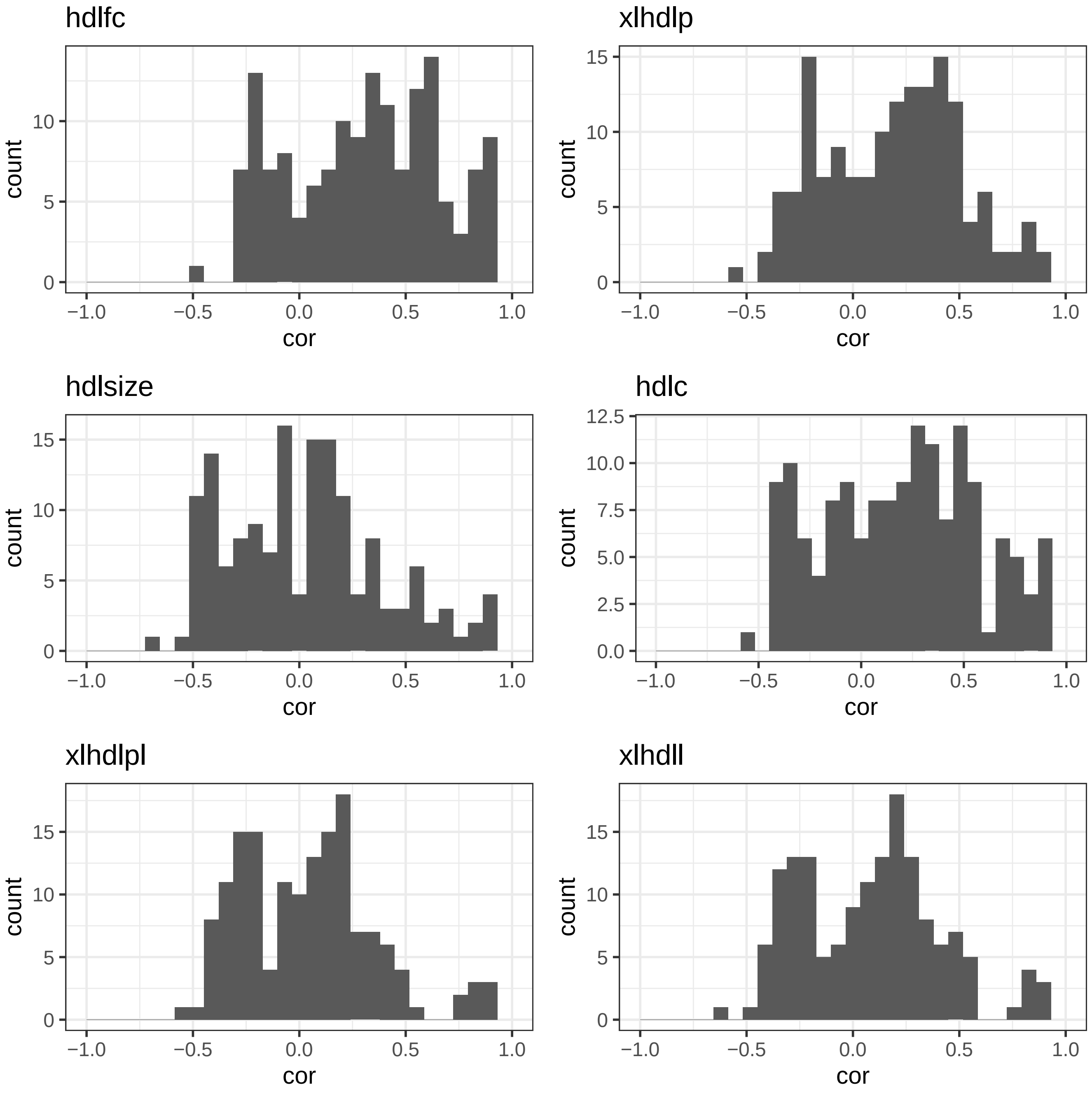
hdlfc
xlhdlp
hdlsize
hdlc
xlhdlpl
xlhdll
count
cor
0
5
10
15
0.0
2.5
5.0
7.5
10.0
12.5
−1.0
−0.5
0.0
0.5
1.0

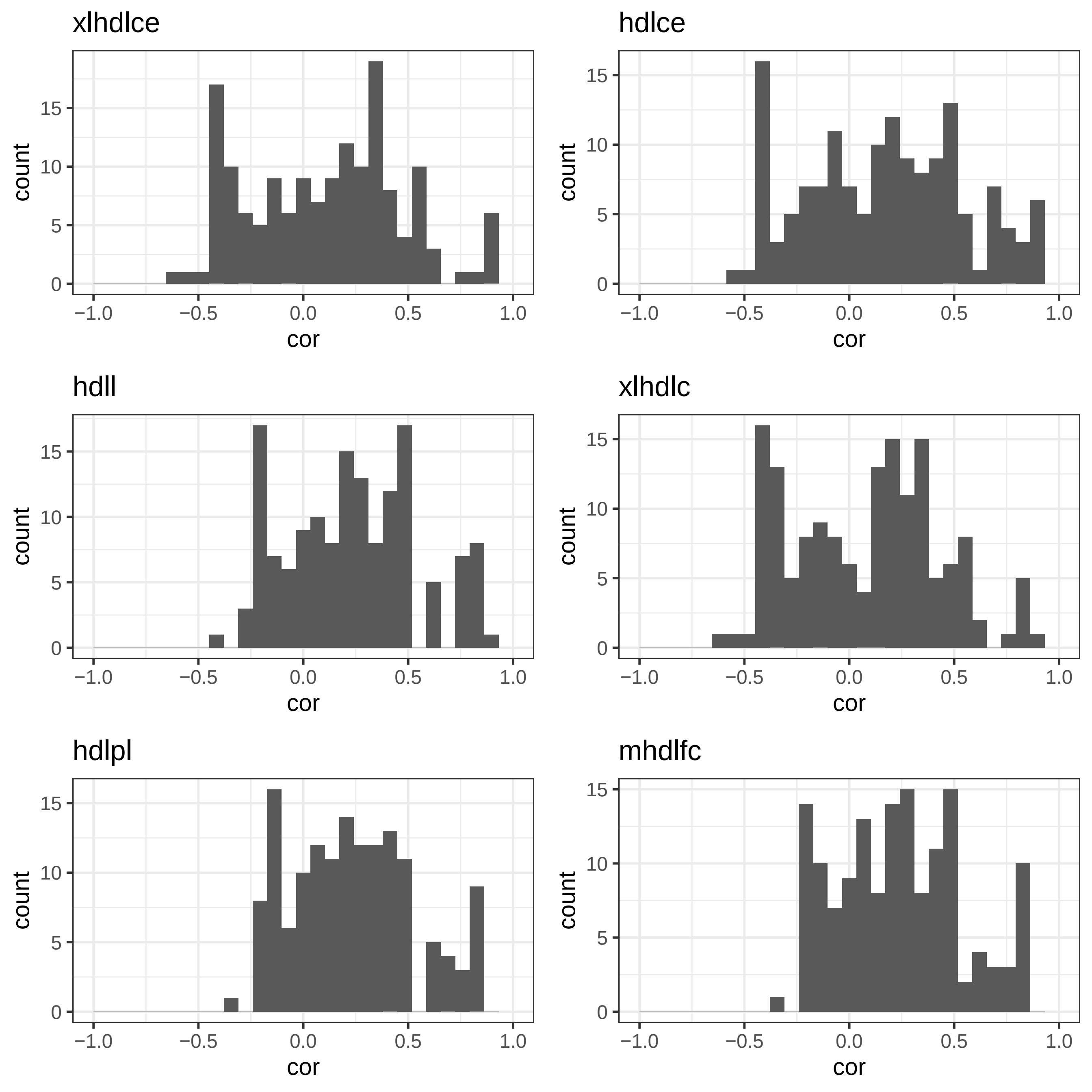

xlhdlce
hdlce
hdll
xlhdlc
hdlpl
mhdlfc
count
cor
0
5
10
15
−1.0
−0.5
0.0
0.5
1.0

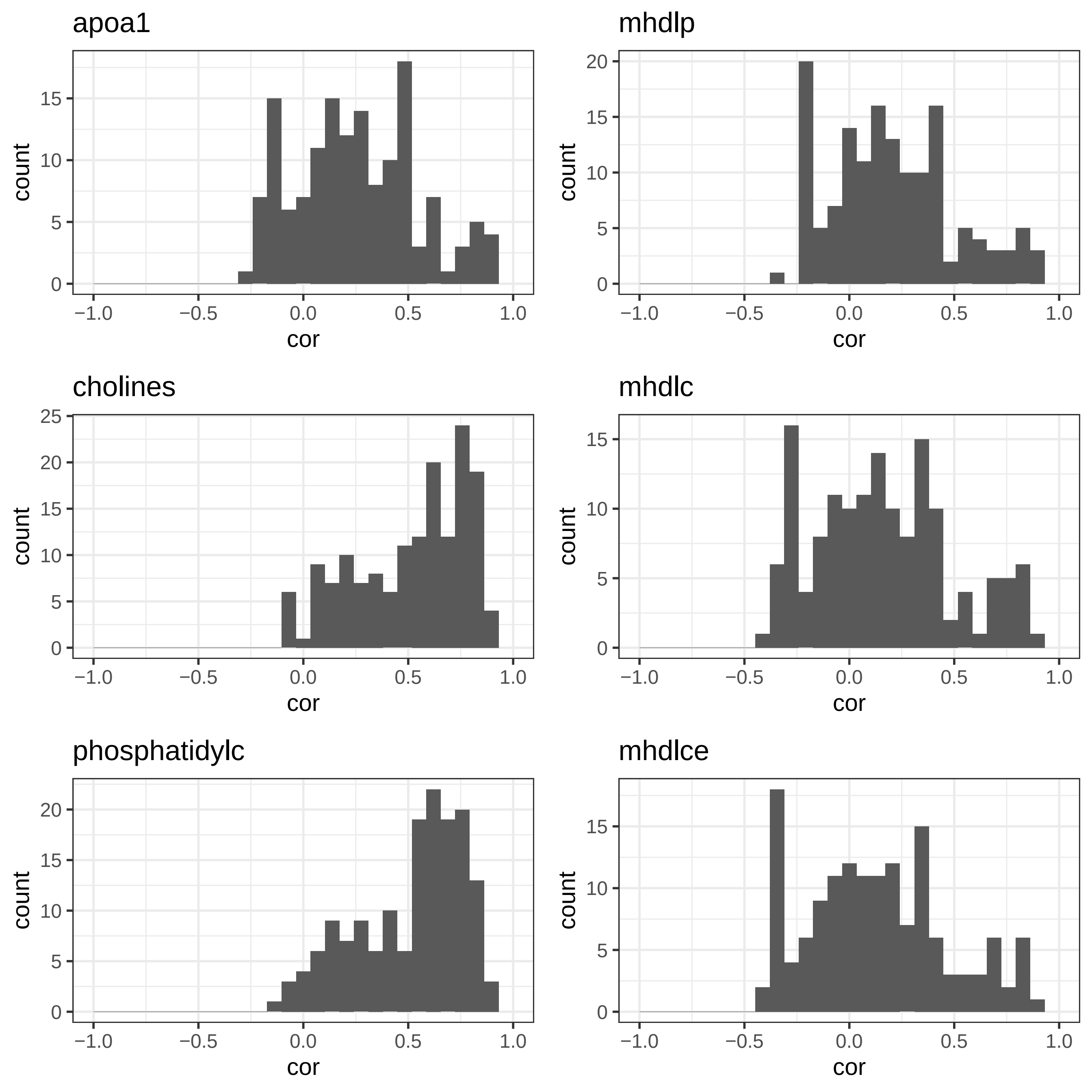

apoa1
mhdlp
cholines
mhdlc
phosphatidylc
mhdlce
count
cor

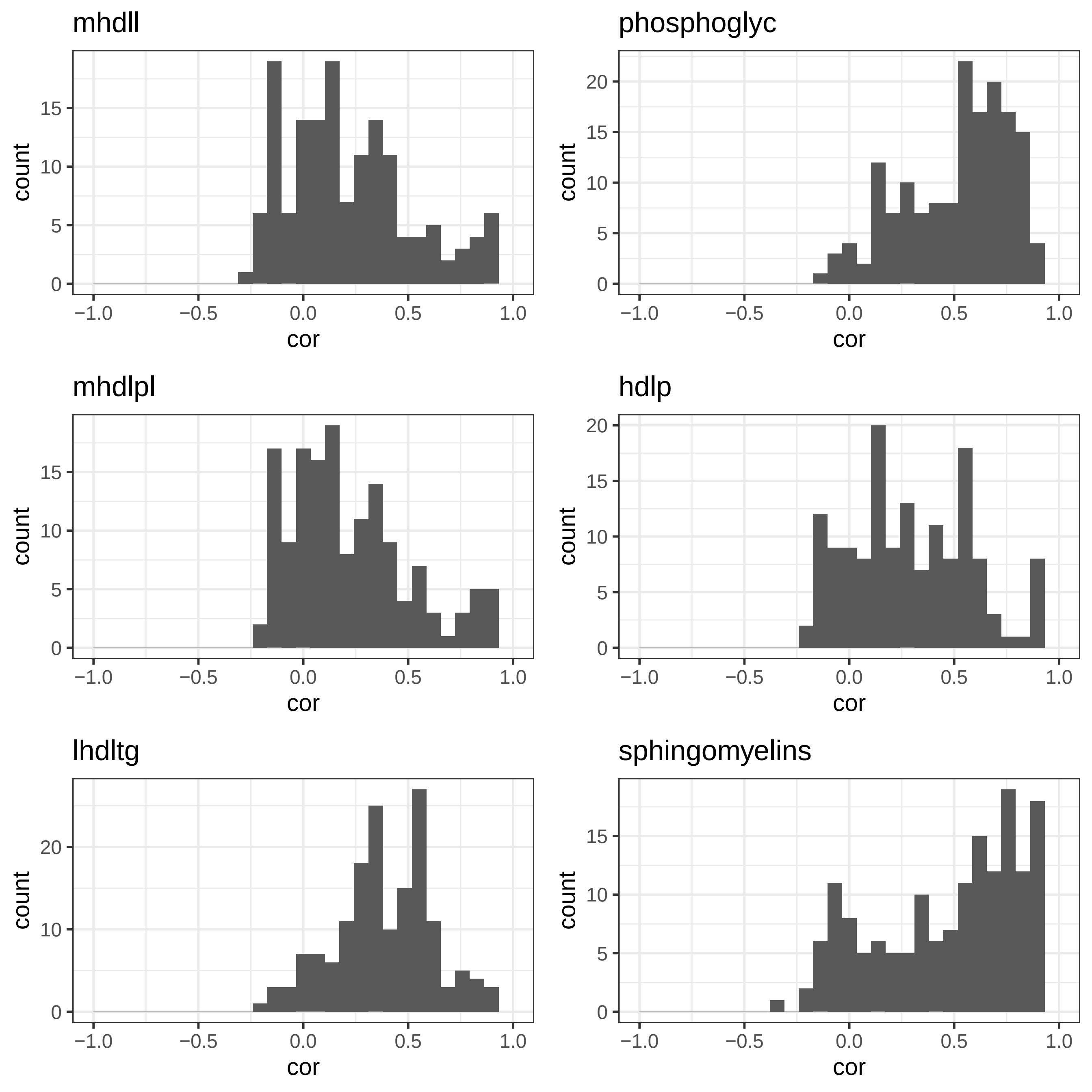

mhdll
phosphoglyc
mhdlpl
hdlp
lhdltg
sphingomyelins
count
cor

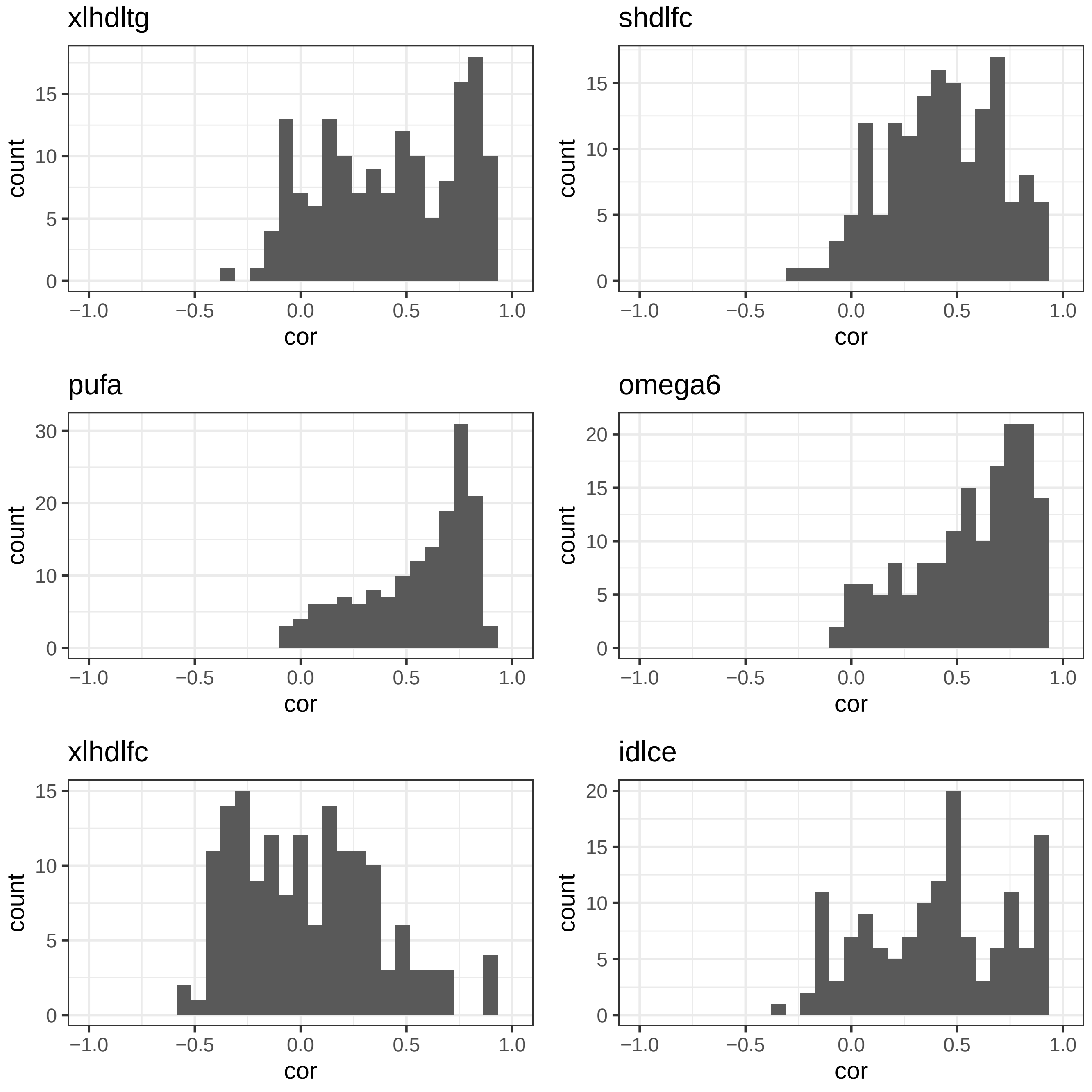

xlhdltg
shdlfc
pufa
omega6
xlhdlfc
idlce
count
cor

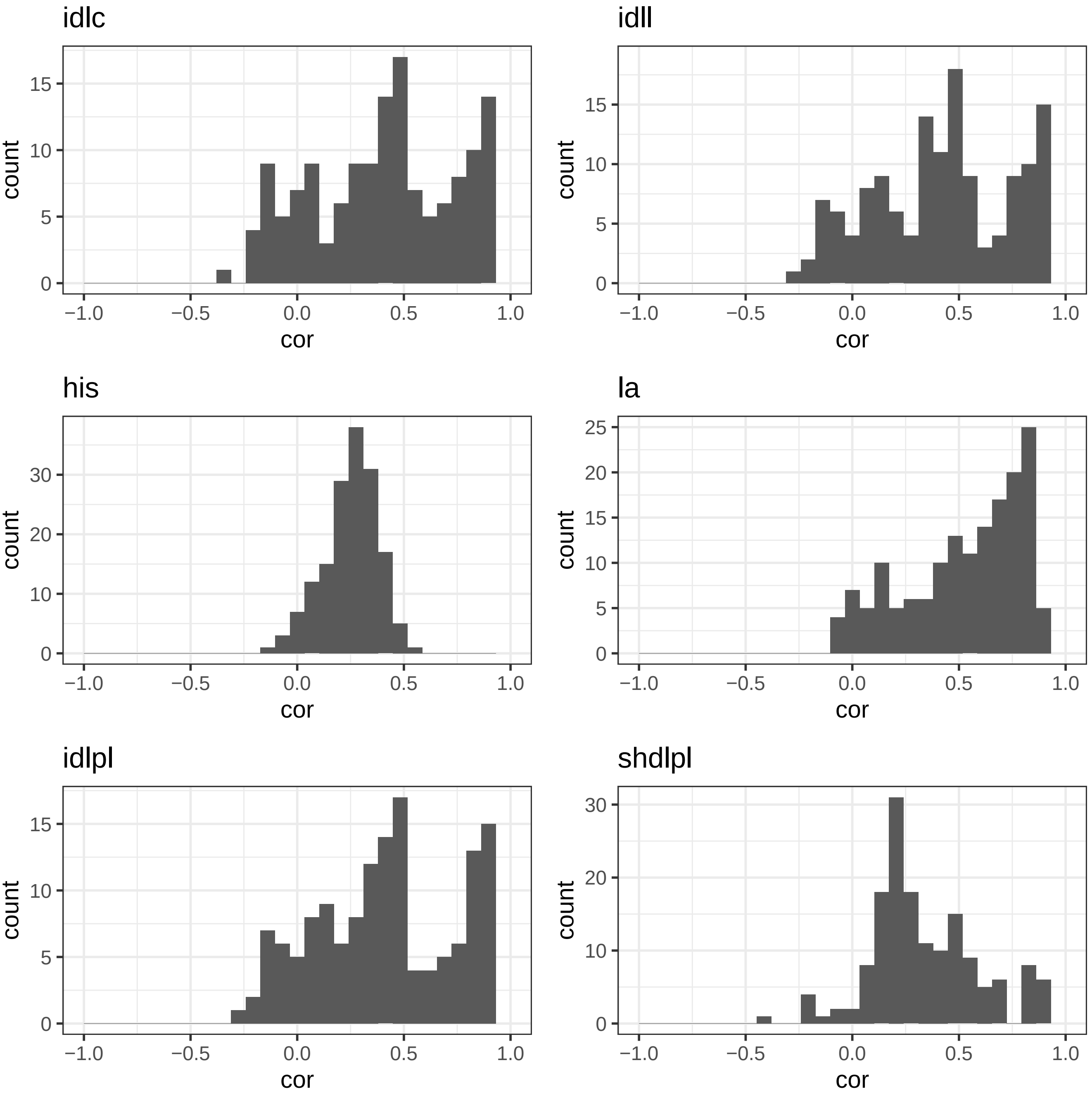
idlc
idll
his
la
idlpl
shdlpl
count
cor
0
5
10
15
20
25
30
−1.0
−0.5
0.0
0.5
1.0

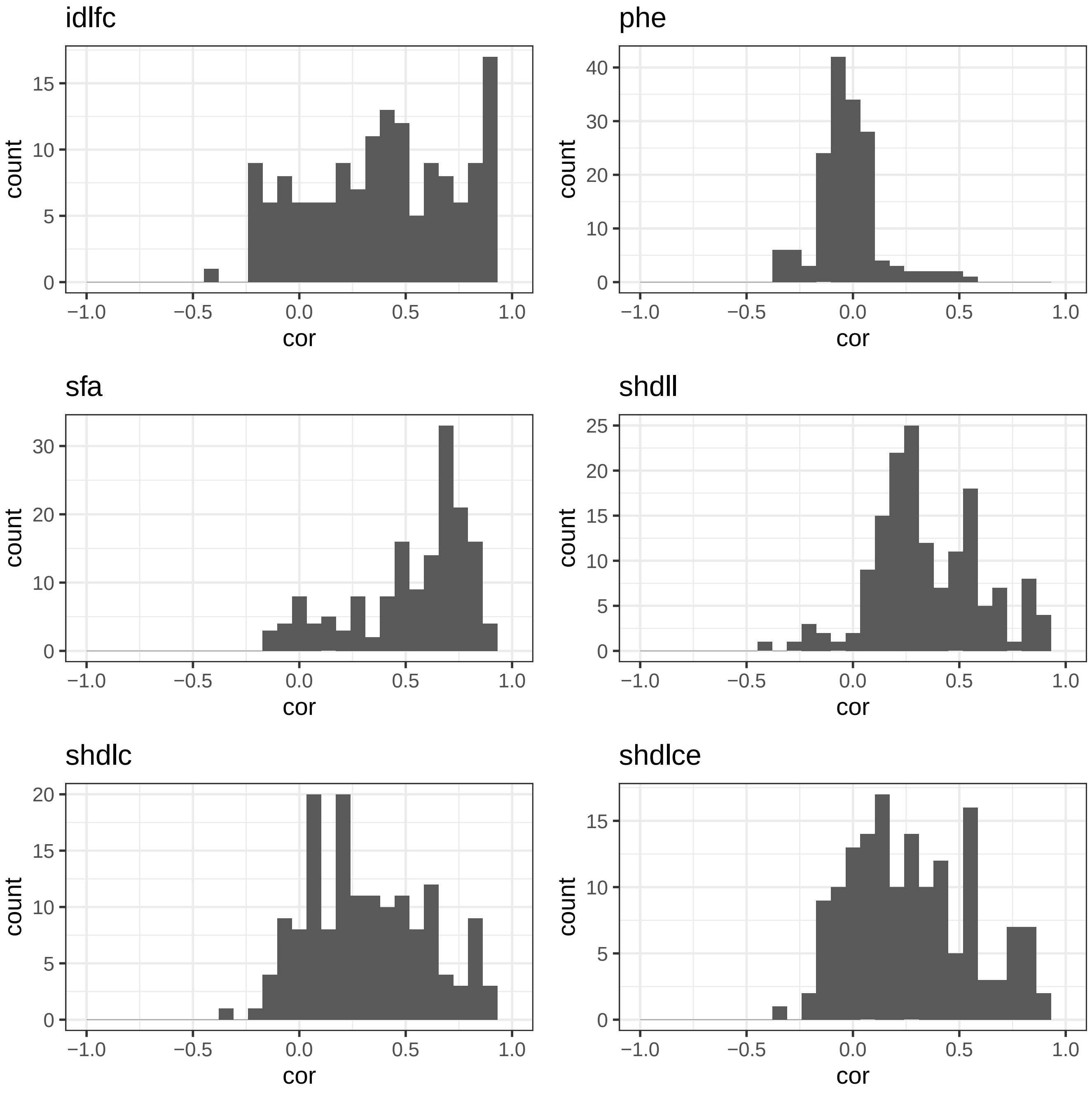
idlfc
phe
sfa
shdll
shdlc
shdlce
count
cor
−1.0
−0.5
0.0
0.5
1.0

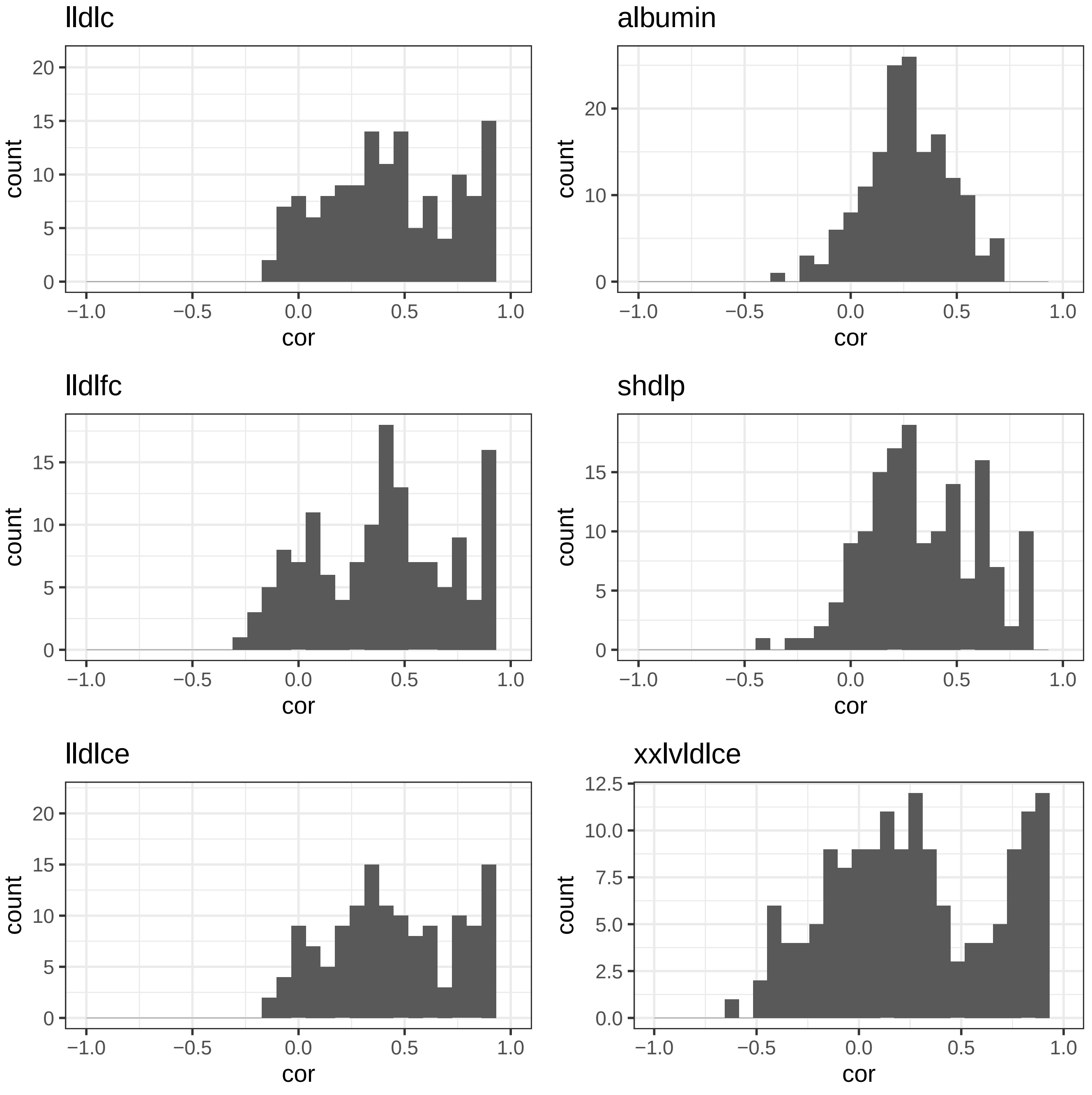
lldlc
albumin
lldlfc
shdlp
lldlce
xxlvldlce
count
cor
-1.0
-0.5
0.0
0.5
1.0
0
5
10
15
20
2.5
5.0
7.5
10.0
12.5

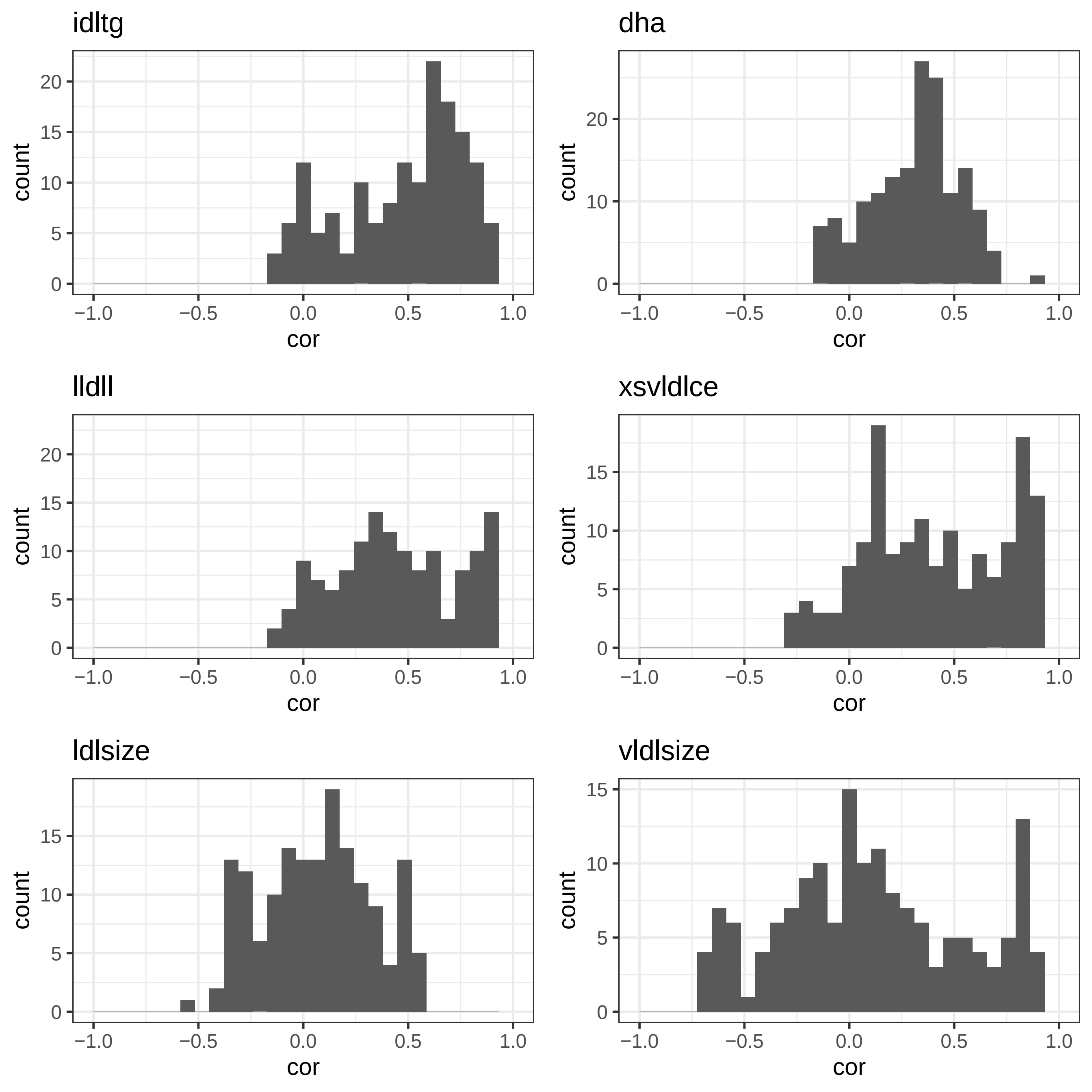
idltg
dha
lldll
xsvldlce
ldlsize
vldlsize
count
cor
20
15
10
5
0
−1.0
−0.5
0.0
0.5
1.0

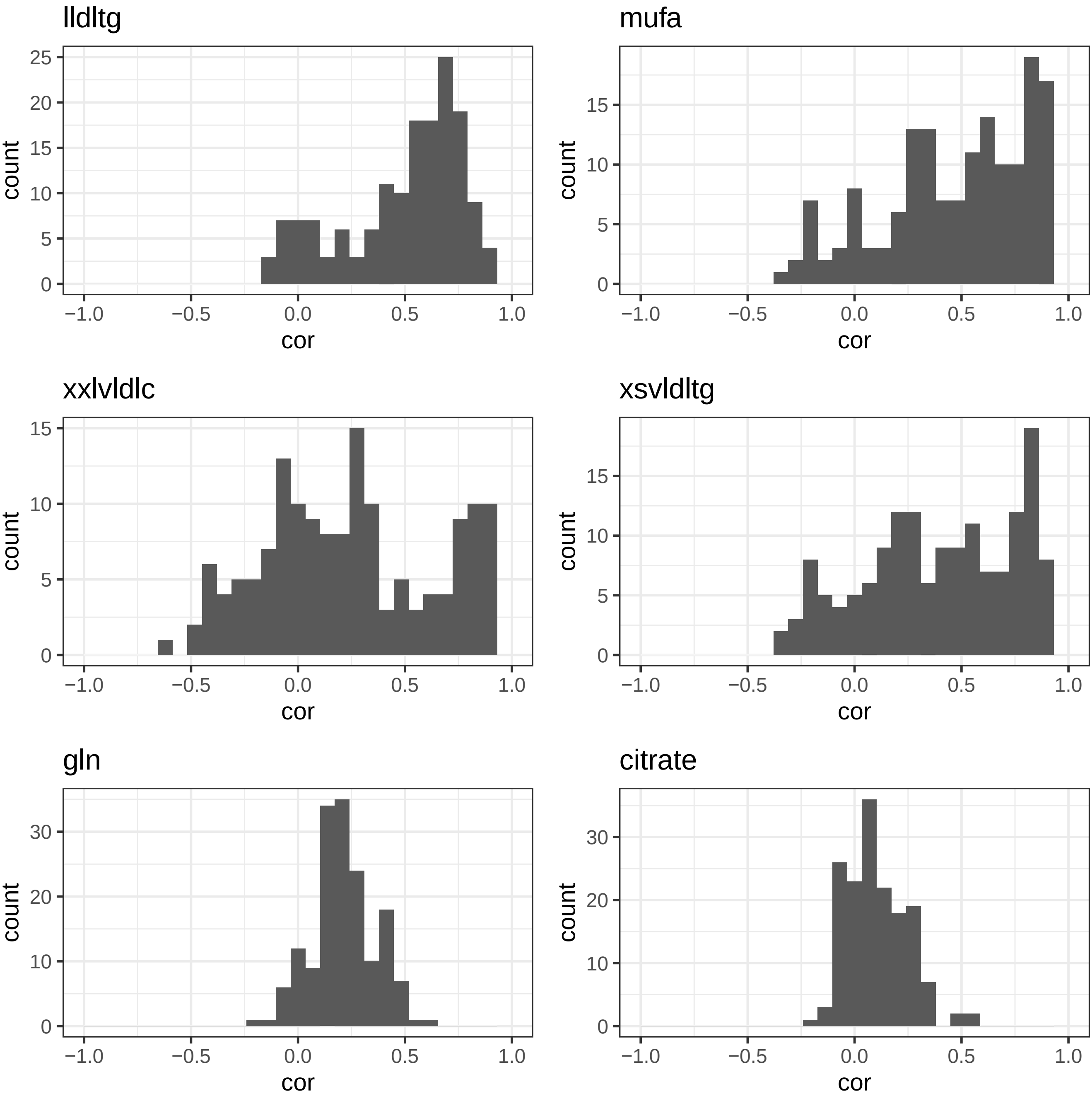
lldltg
mufa
xxlvldlc
xsvldltg
gln
citrate
count
cor
−1.0
−0.5
0.0
0.5
1.0

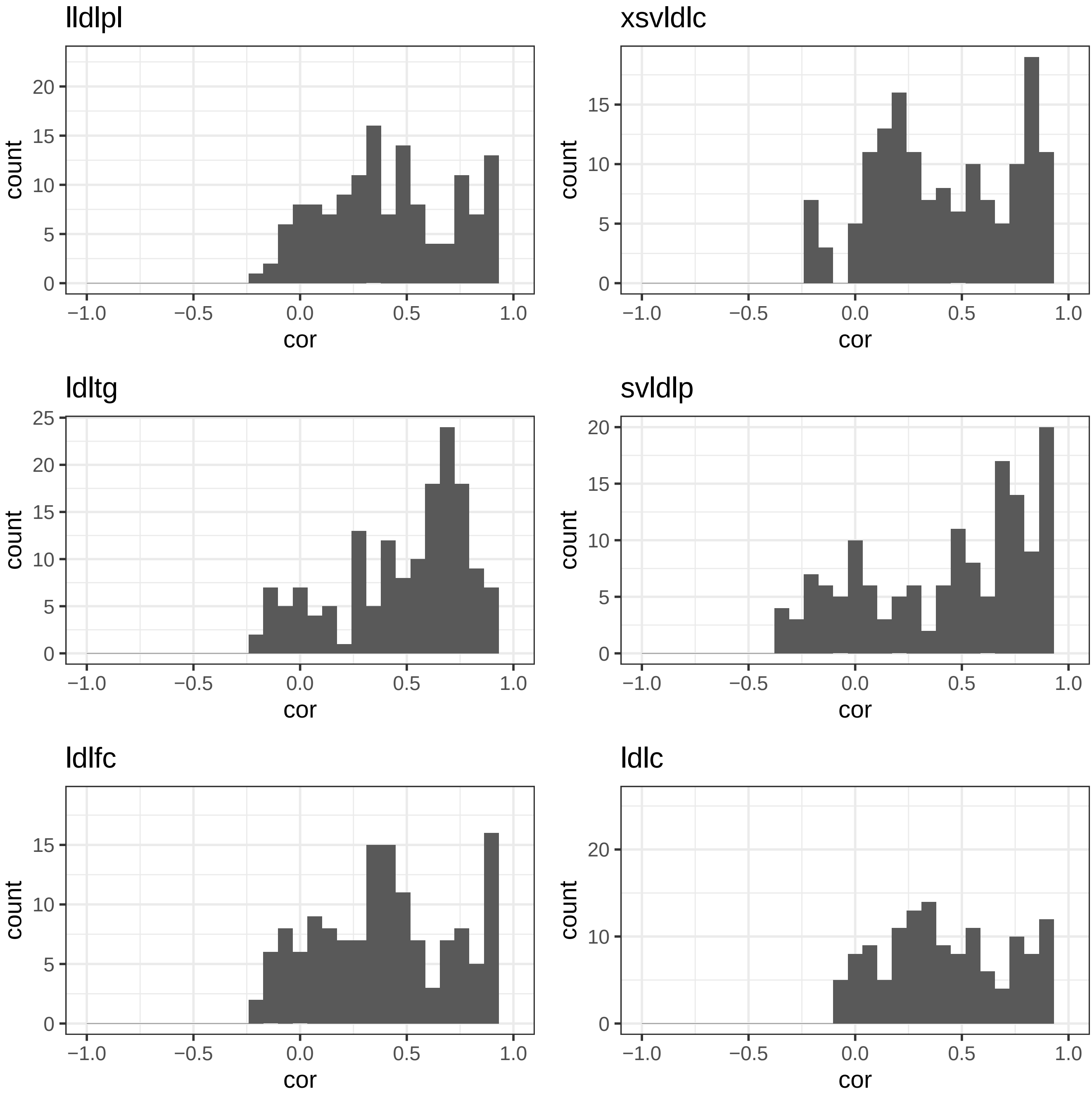
lldlpl
xsvldlc
ldltg
svldlp
ldlfc
ldlc
count
cor
−1.0
−0.5
0.0
0.5
1.0
0
5
10
15
20
25

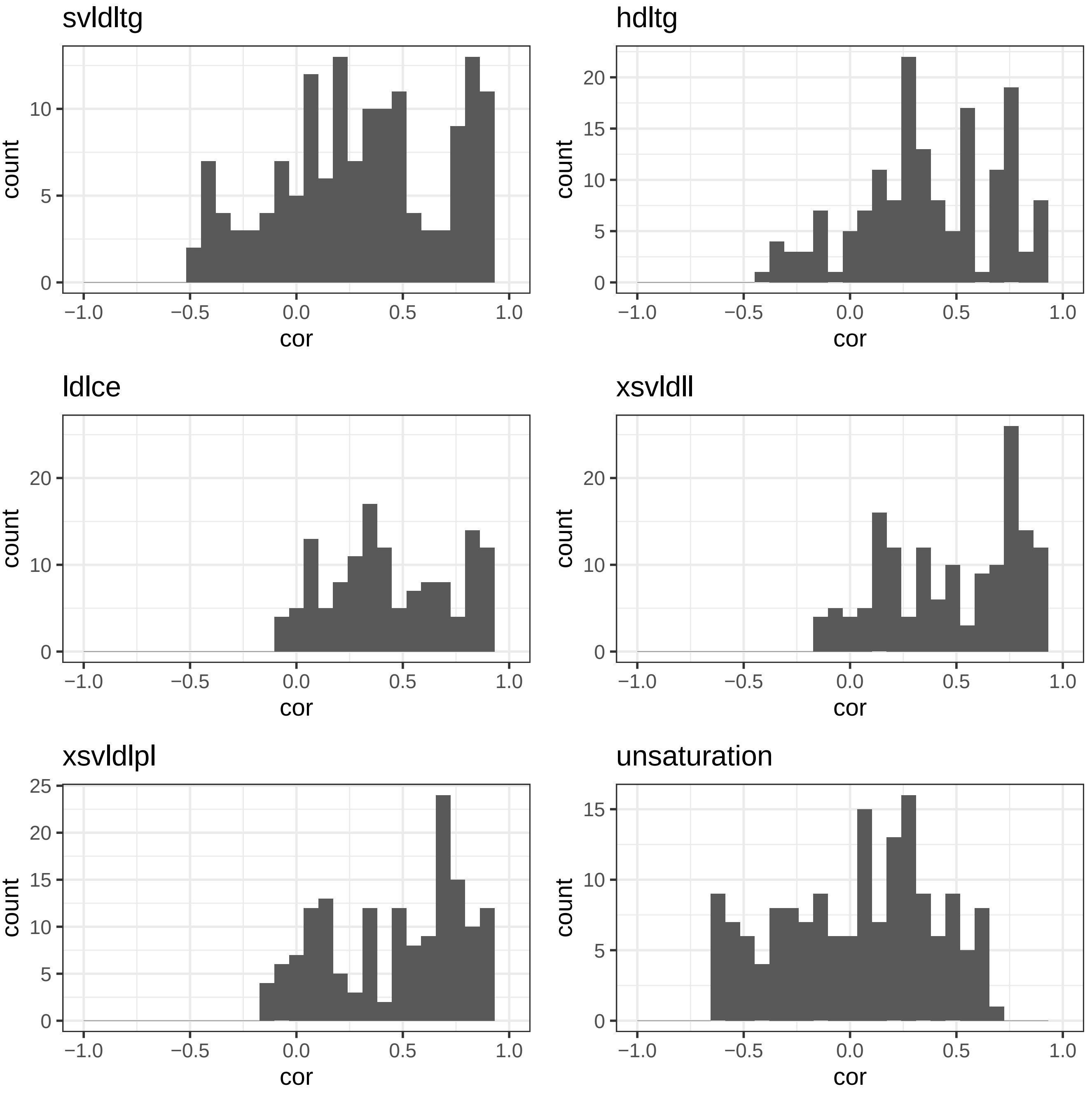
svldltg
count
cor
hdltg
count
cor
ldlce
count
cor
xsvldll
count
cor
xsvldlpl
count
cor
unsaturation
count
cor
−1.0
−0.5
0.0
0.5
1.0
0
5
10
15
20
25

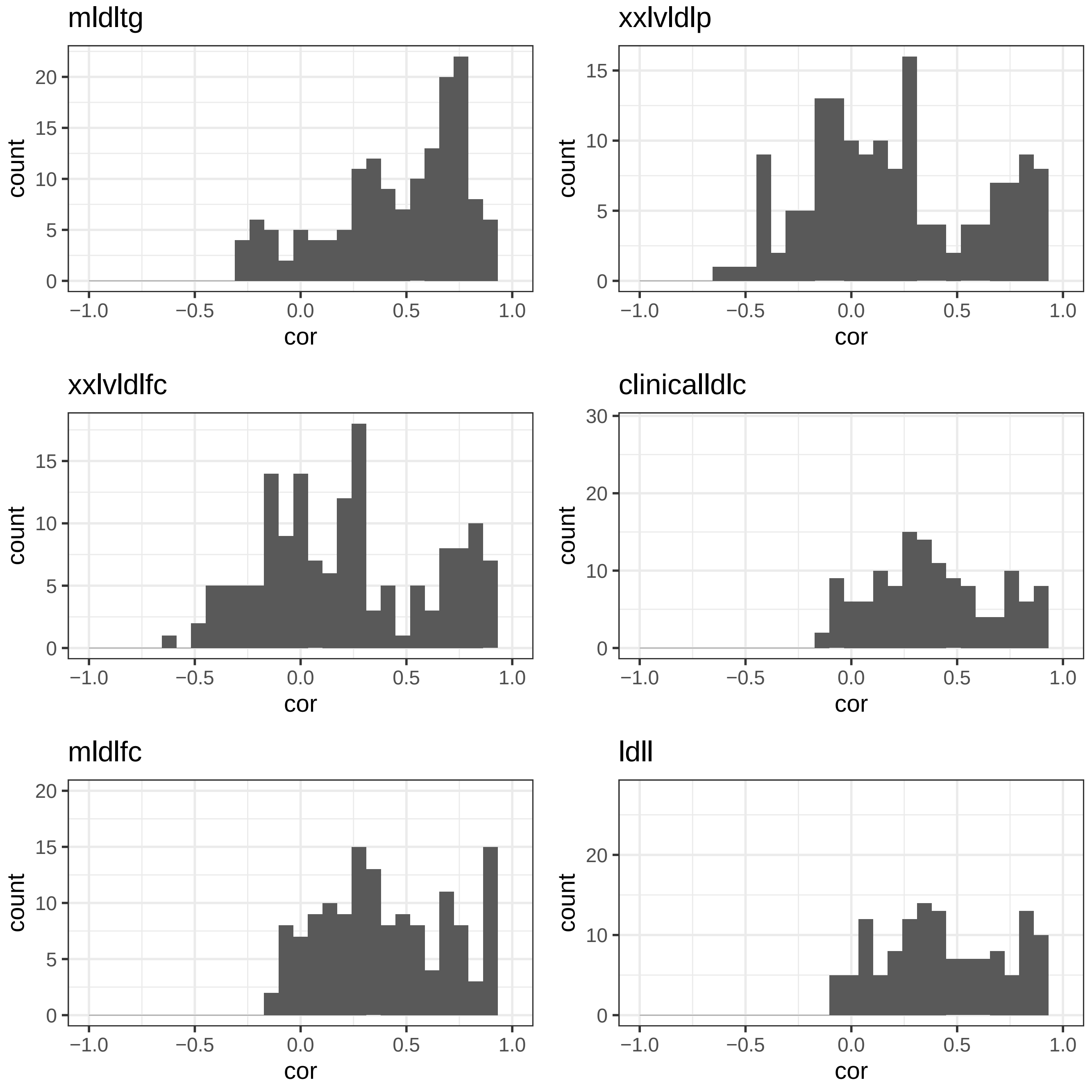
mldltg
xxlvldlp
xxlvldlfc
clinicalldlc
mldlfc
ldll
count
cor
−1.0
−0.5
0.0
0.5
1.0

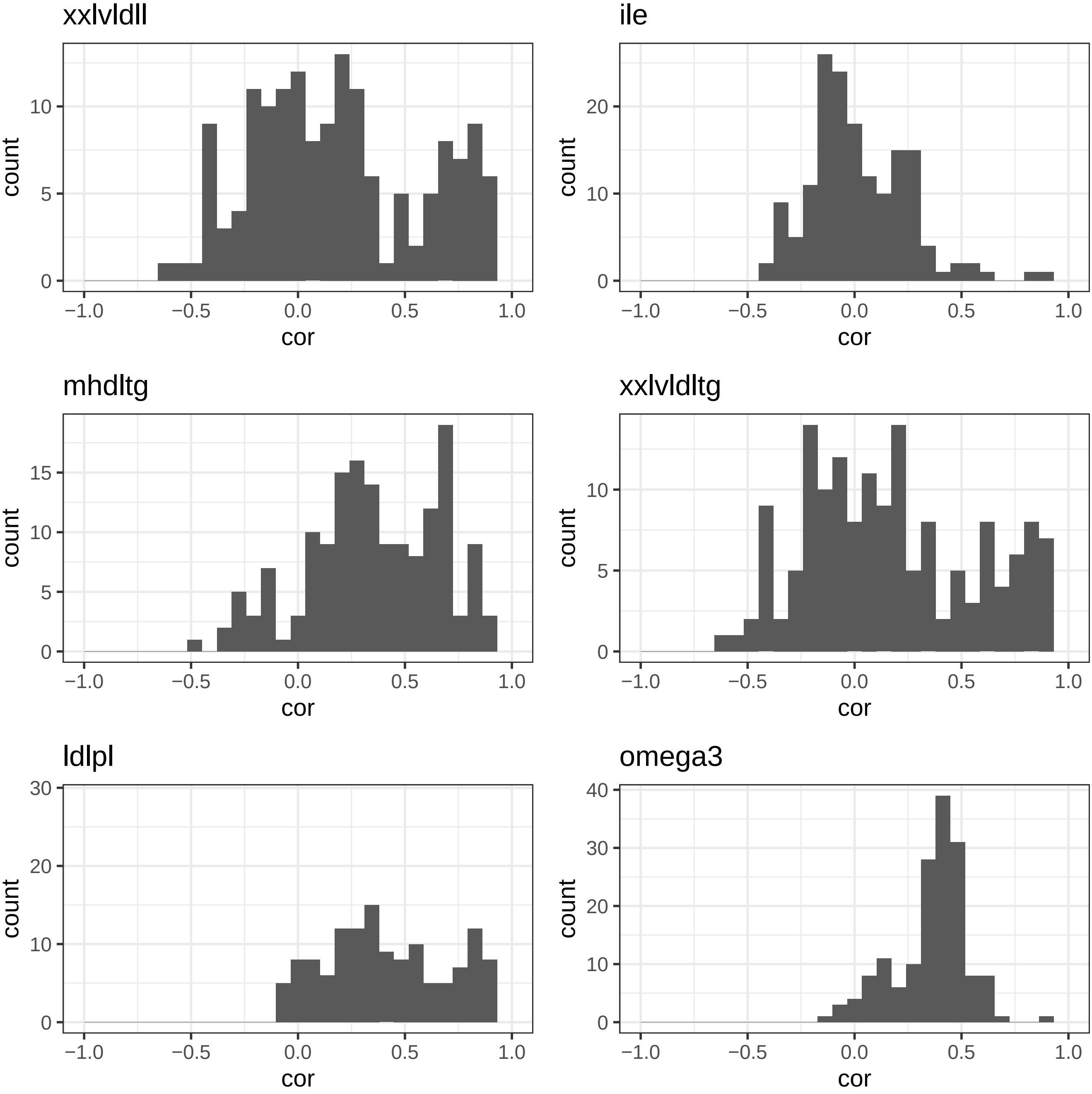

xxlvldll
ile
mhdltg
xxlvldltg
ldlpl
omega3
count
cor
−1.0
−0.5
0.0
0.5
1.0
0
5
10
15
20
30
40

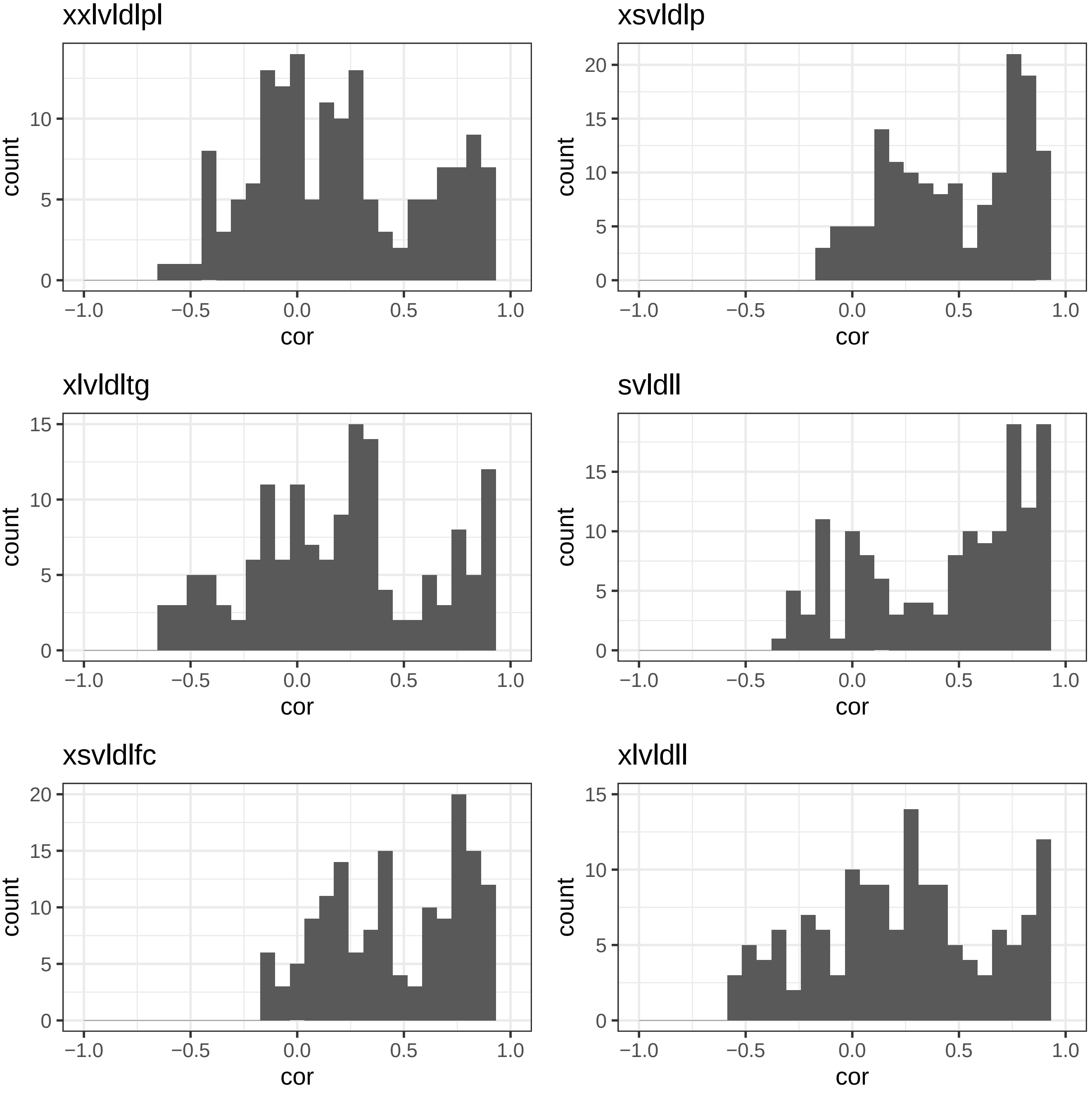
xxlvldlpl
xsvldlp
xlvldltg
svldll
xsvldlfc
xlvldll
count
cor
0
5
10
15
20
−1.0
−0.5
0.0
0.5
1.0

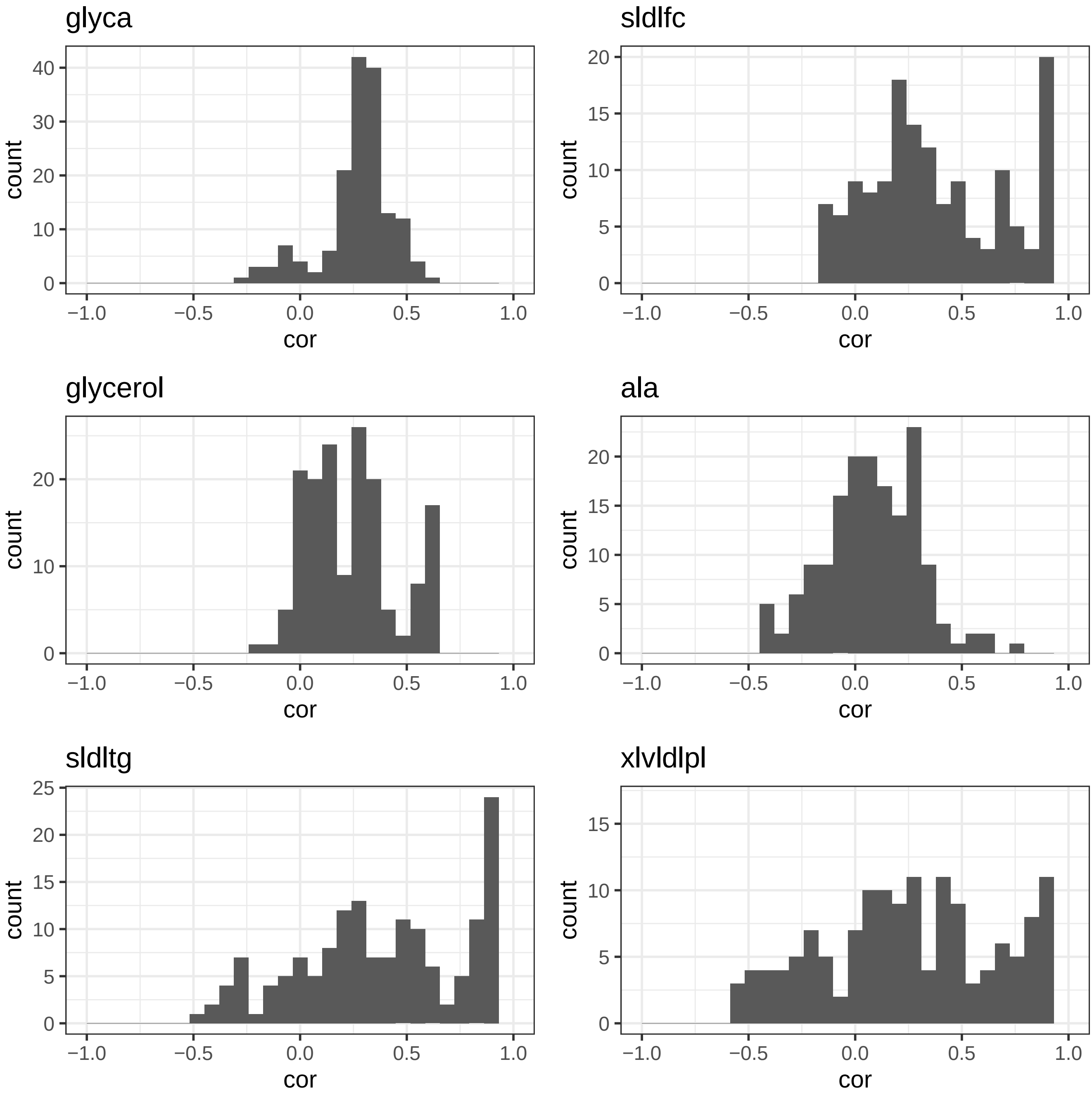
glyca
sldlfc
glycerol
ala
sldltg
xlvldlpl
count
cor
−1.0
−0.5
0.0
0.5
1.0
0
5
10
15
20
25
30
40

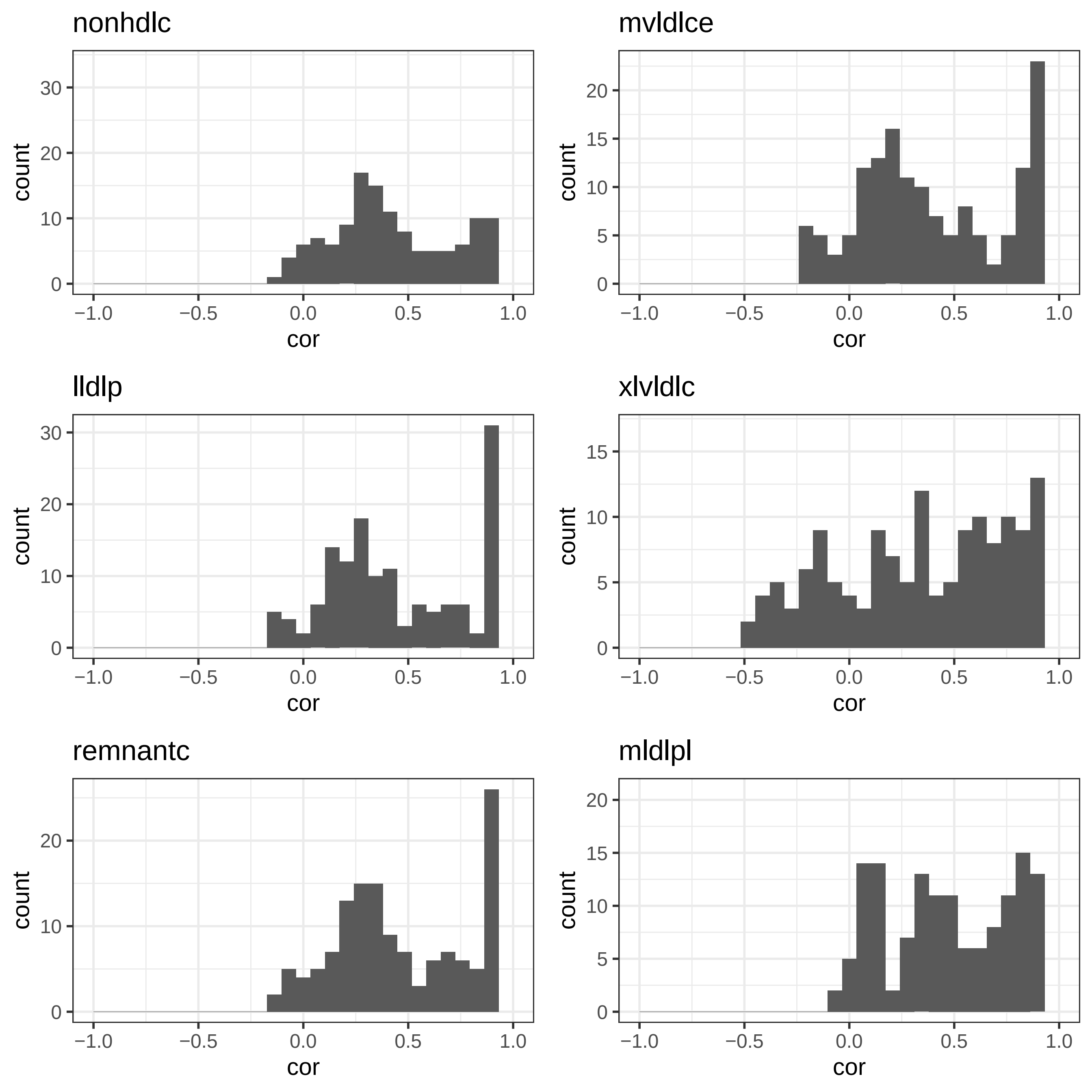

nonhdlc
mvldlce
lldlp
xlvldlc
remnantc
mldlpl
count
cor
-1.0
-0.5
0.0
0.5
1.0
0
5
10
15
20
30

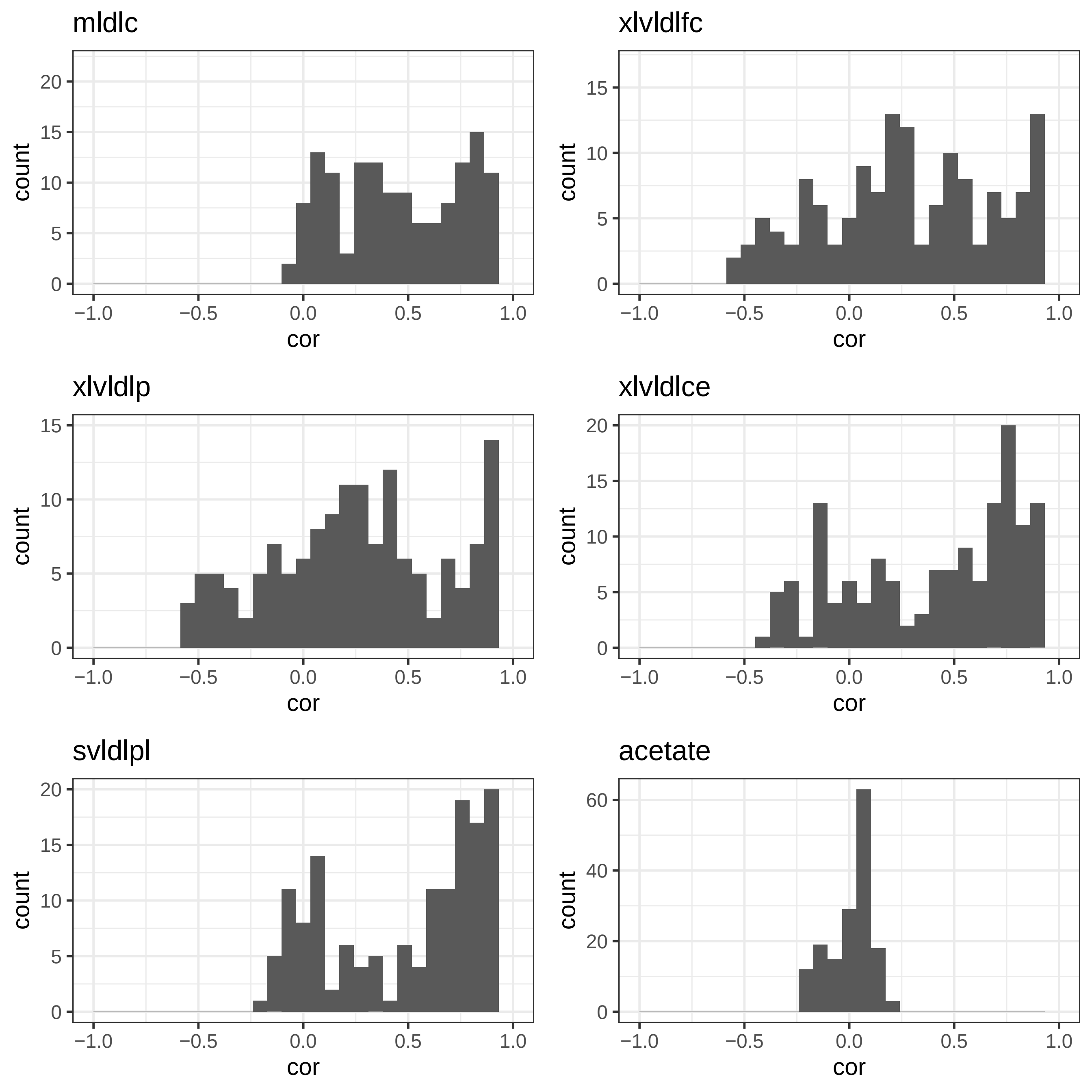

mldlc
xlvldlfc
xlvldlp
xlvldlce
svldlpl
acetate
count
cor
−1.0
−0.5
0.0
0.5
1.0
0
5
10
15
20
40
60

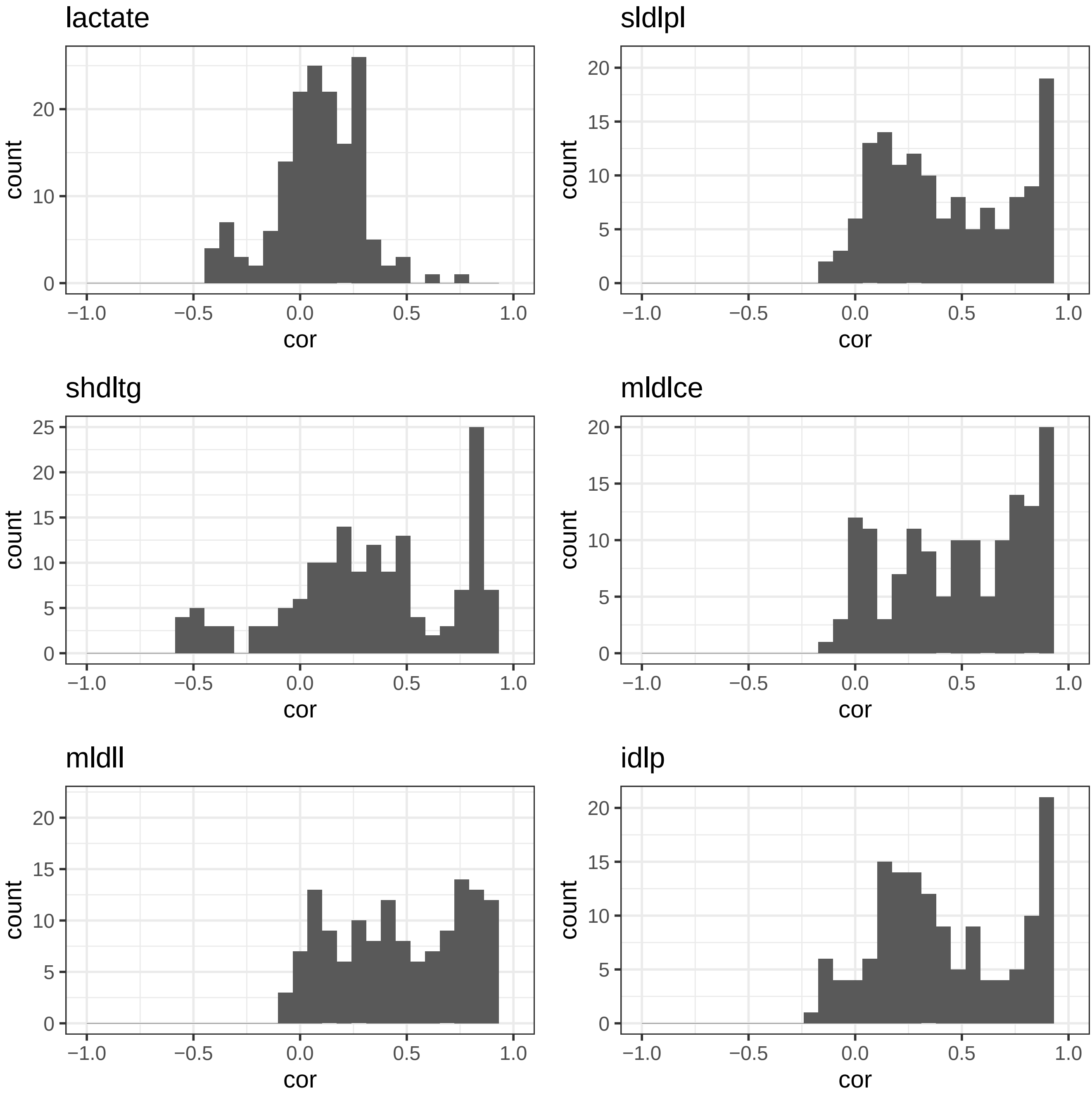
lactate
sldlpl
shdltg
mldlce
mldll
idlp
count
cor
−1.0
−0.5
0.0
0.5
1.0
0
5
10
15
20
25

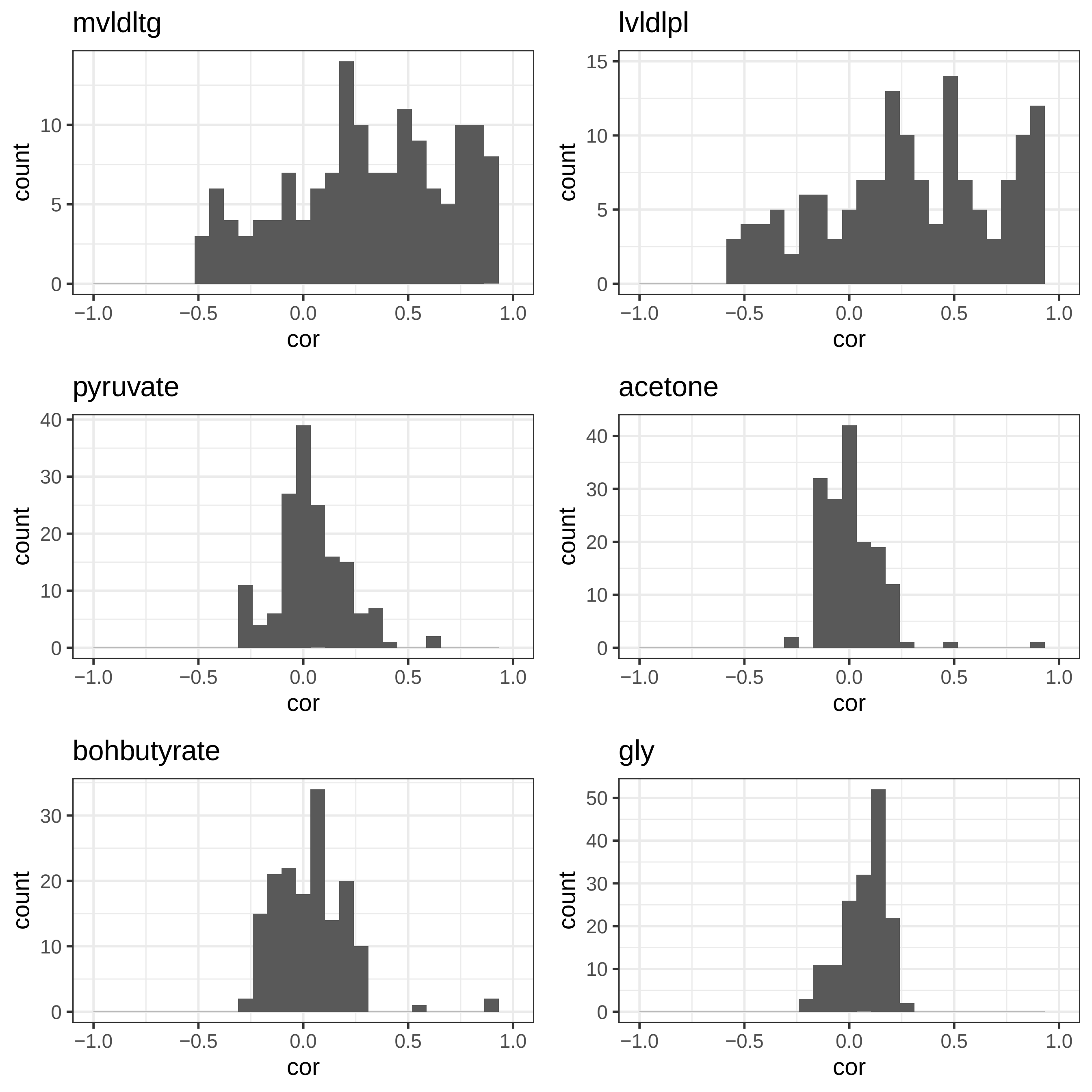
mvldltg
lvldlpl
pyruvate
acetone
bohbutyrate
gly
count
cor
−1.0
−0.5
0.0
0.5
1.0
0
5
10
15
20
30
40
50

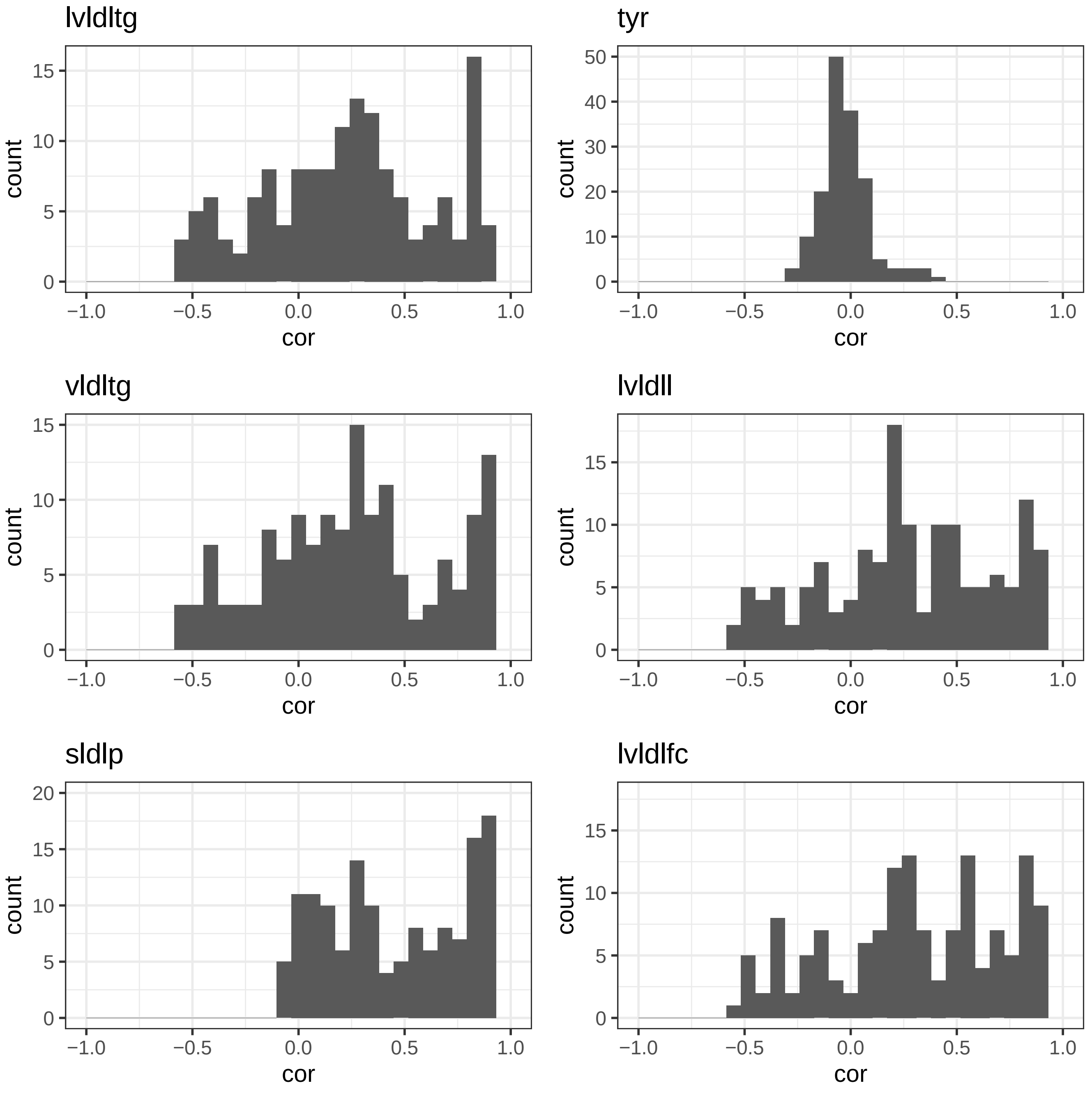

lvldltg
tyr
vldltg
lvldll
sldlp
lvldlfc
count
cor

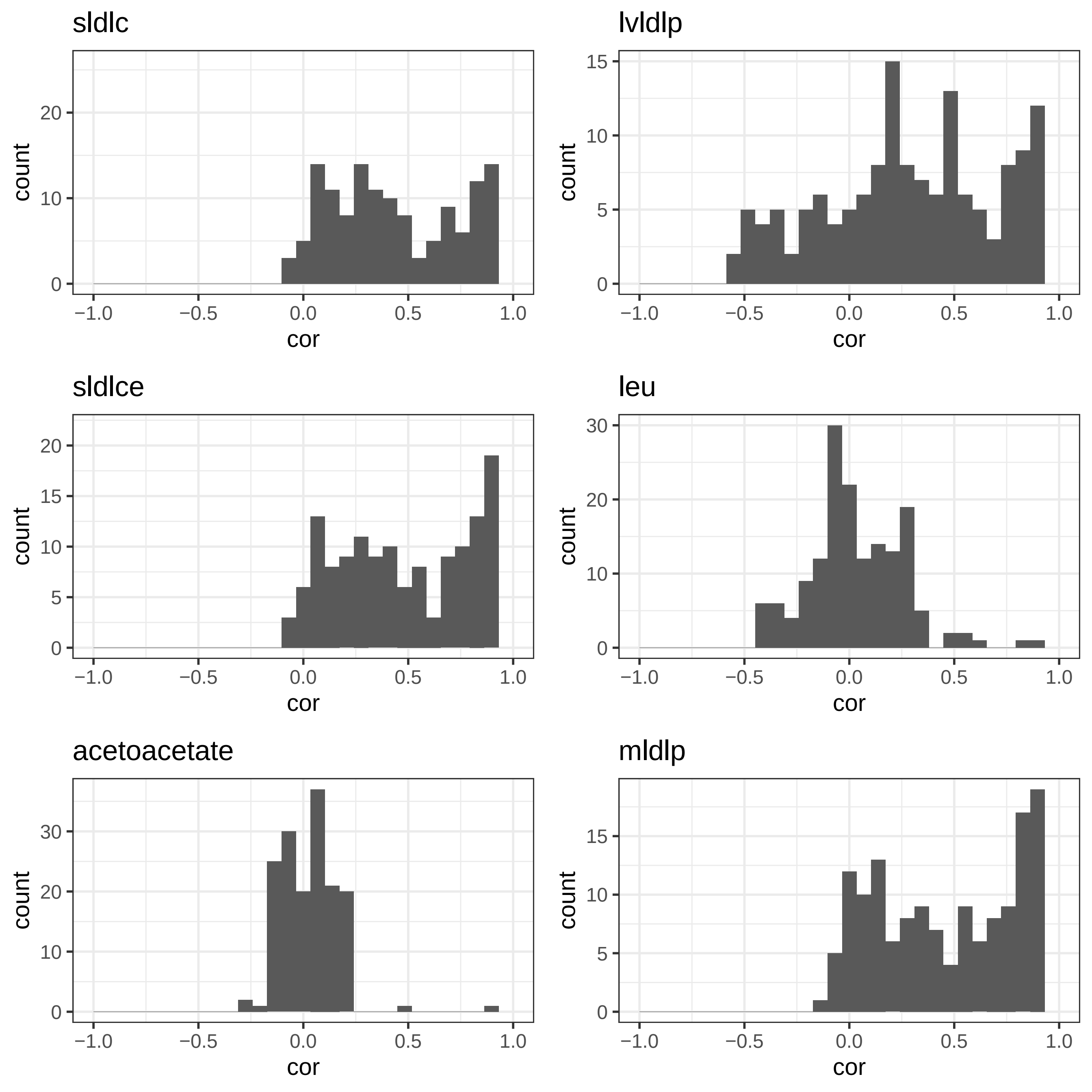
sldlc
lvldlp
sldlce
leu
acetoacetate
mldlp
count
cor
−1.0
−0.5
0.0
0.5
1.0
0
5
10
15
20
30

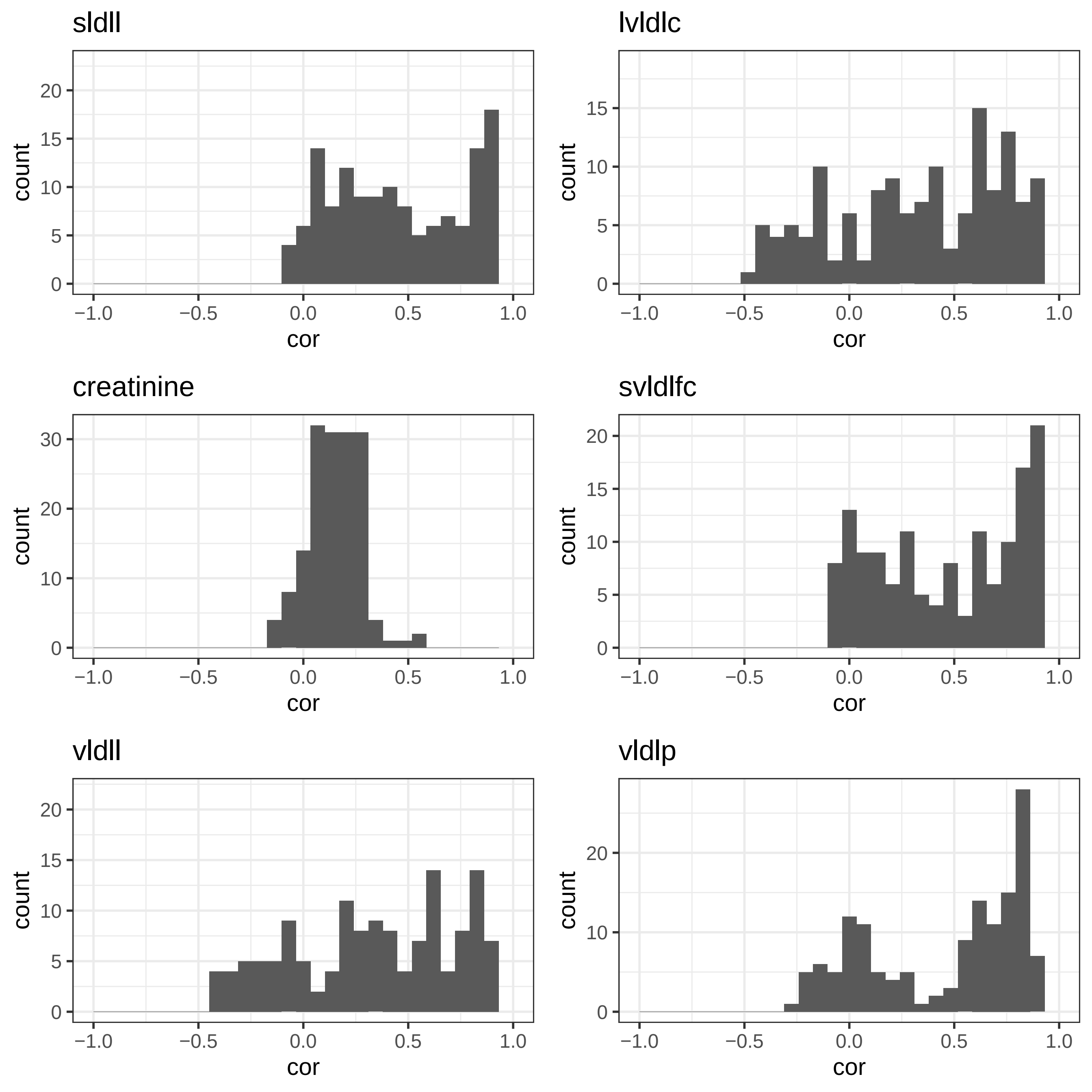

sldll
lvldlc
creatinine
svldlfc
vldll
vldlp
count
cor

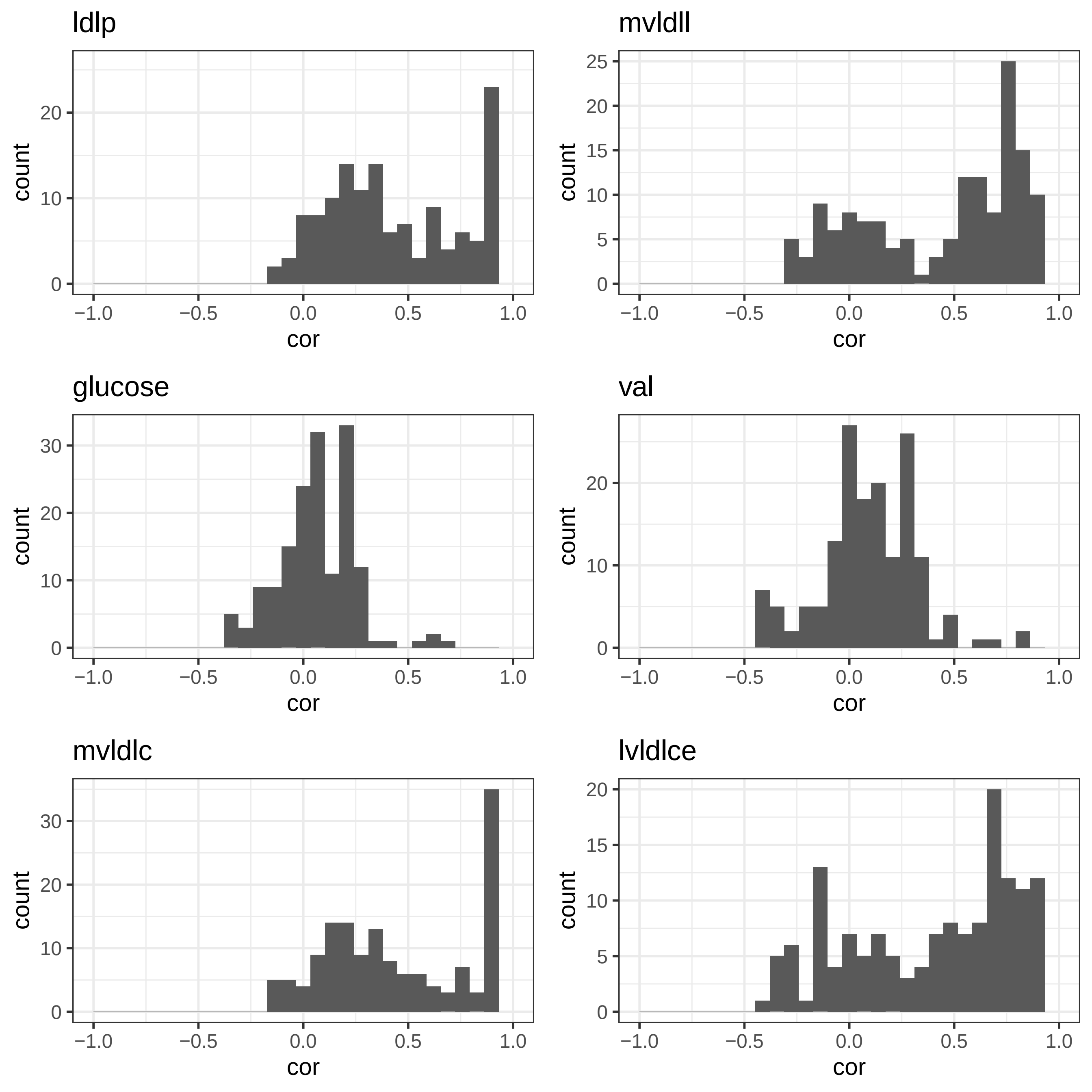

ldlp
mvldll
glucose
val
mvldlc
lvldlce
count
cor

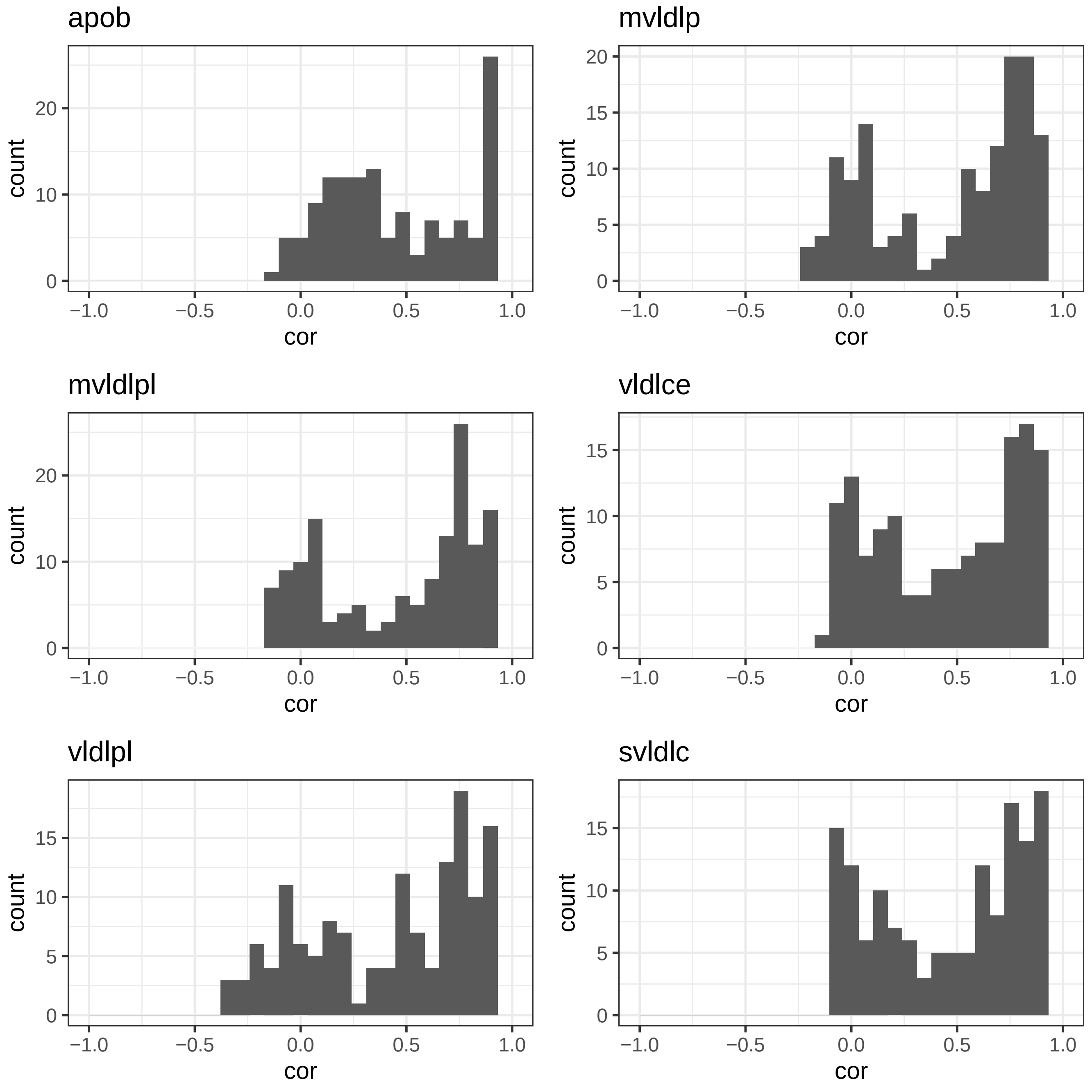
apob
count
cor
20
10
0
−1.0
−0.5
0.0
0.5
1.0
mvldlp
15
5
mvldlpl
vldlce
vldlpl
svldlc

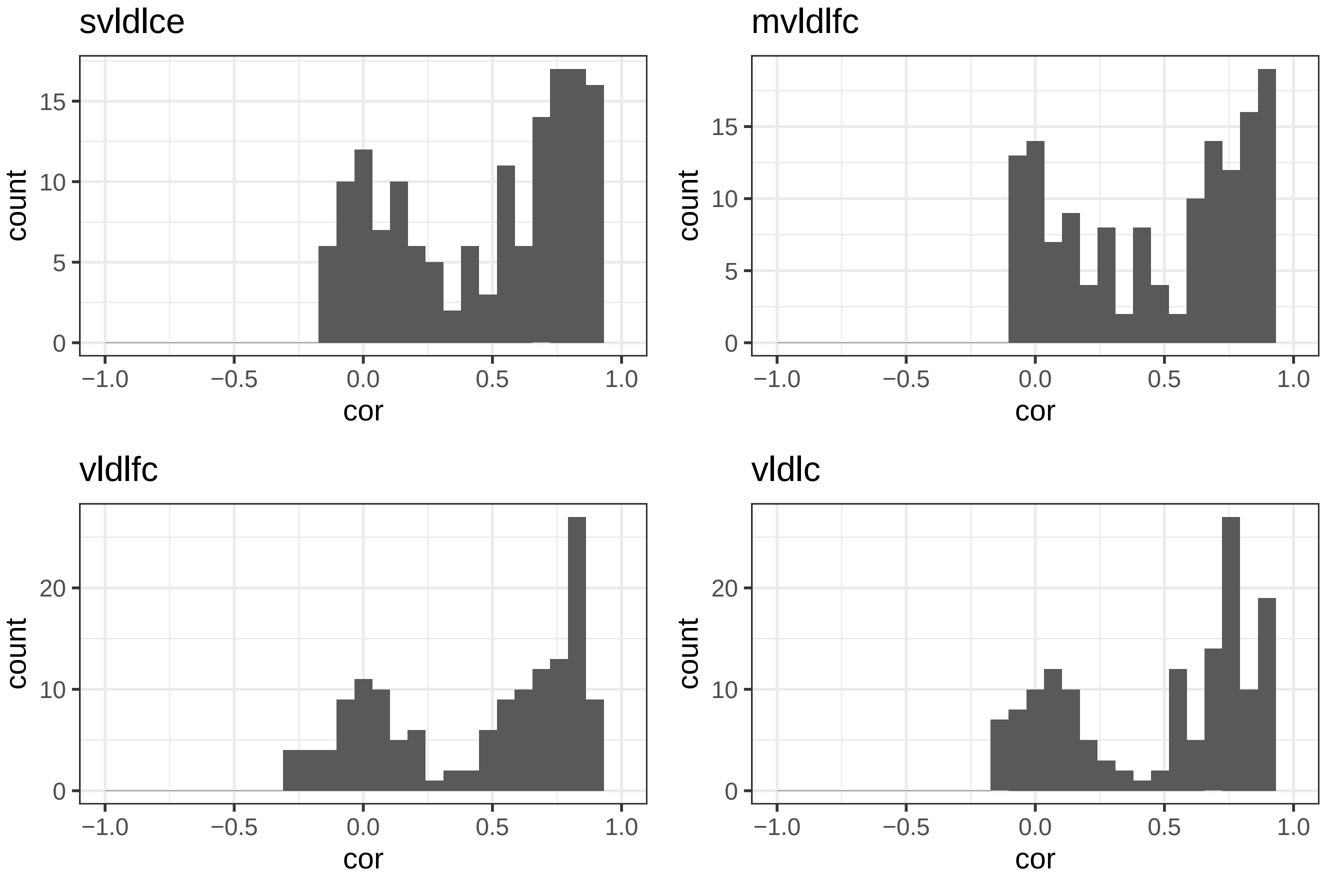

svldlce
mvldlfc
vldlfc
vldlc
count
cor
