## Supplementary Data 4 for "Multivariate analysis of metabolomic data to identify biological pathways modified by a clinical intervention"

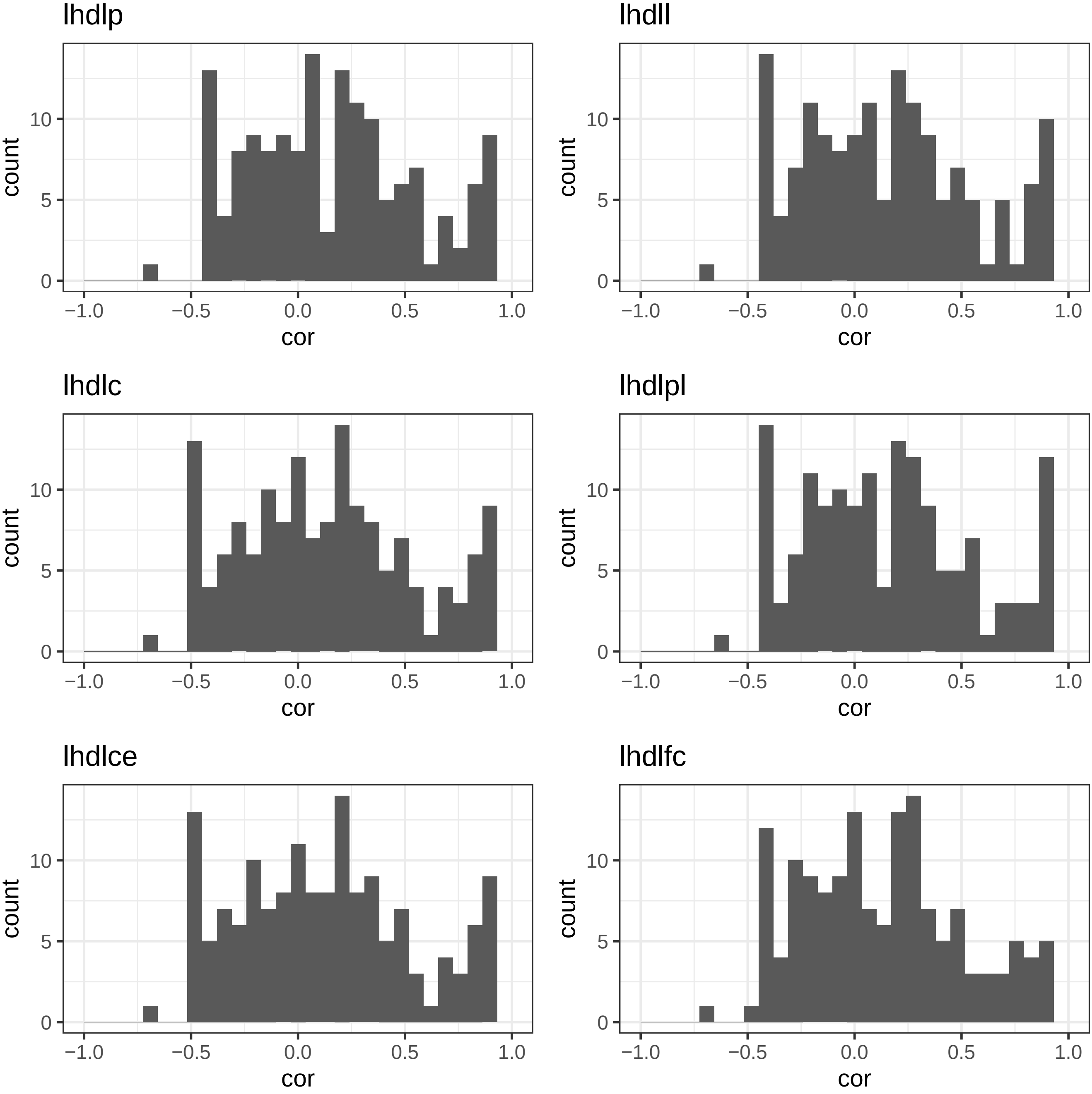
lhdlp
lhdll
lhdlc
lhdlpl
lhdlce
lhdlfc
count
cor
0
5
10
−1.0
−0.5
0.0
0.5
1.0

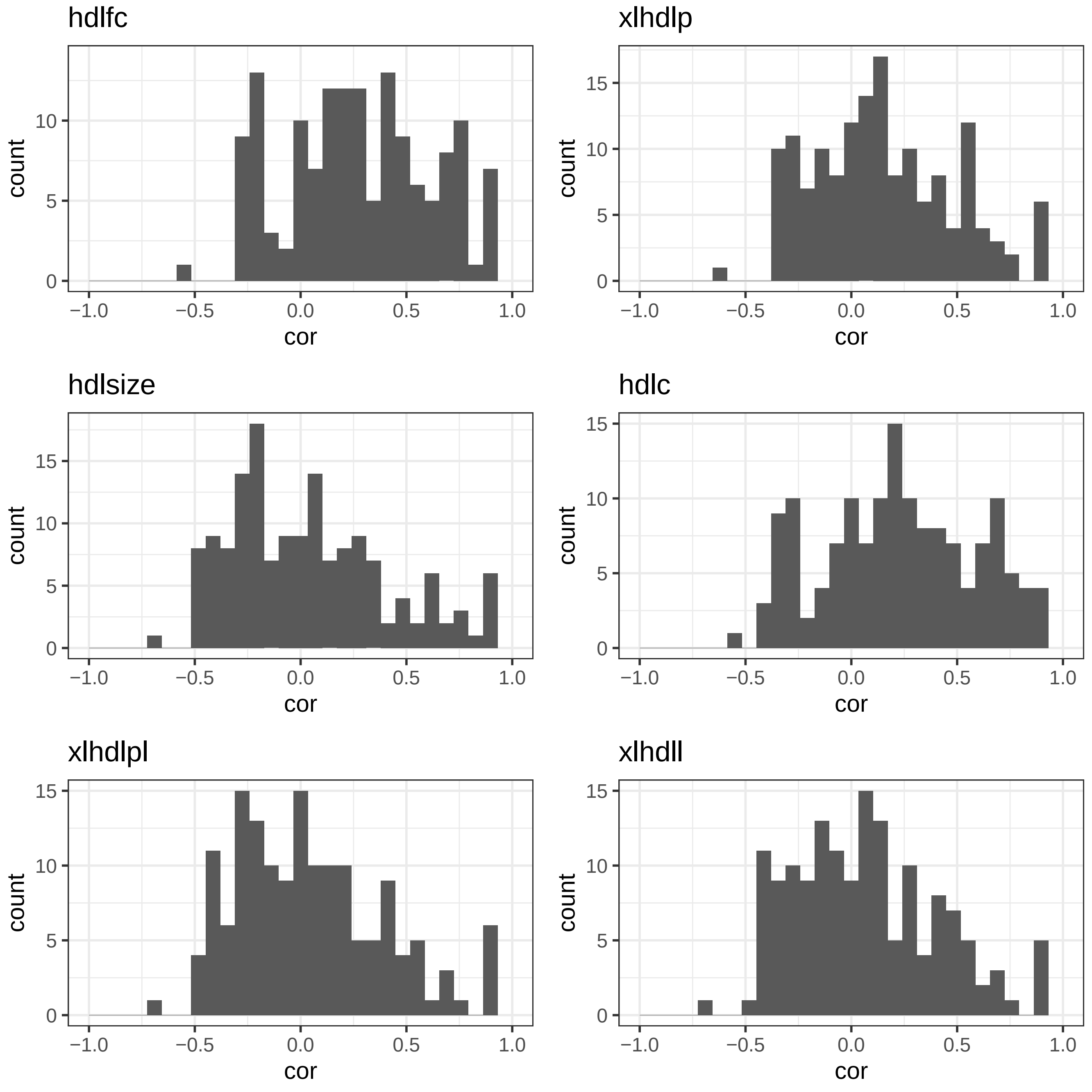
hdlfc
xlhdlp
hdlsize
hdlc
xlhdlpl
xlhdll
count
cor
0
5
10
15
−1.0
−0.5
0.0
0.5
1.0

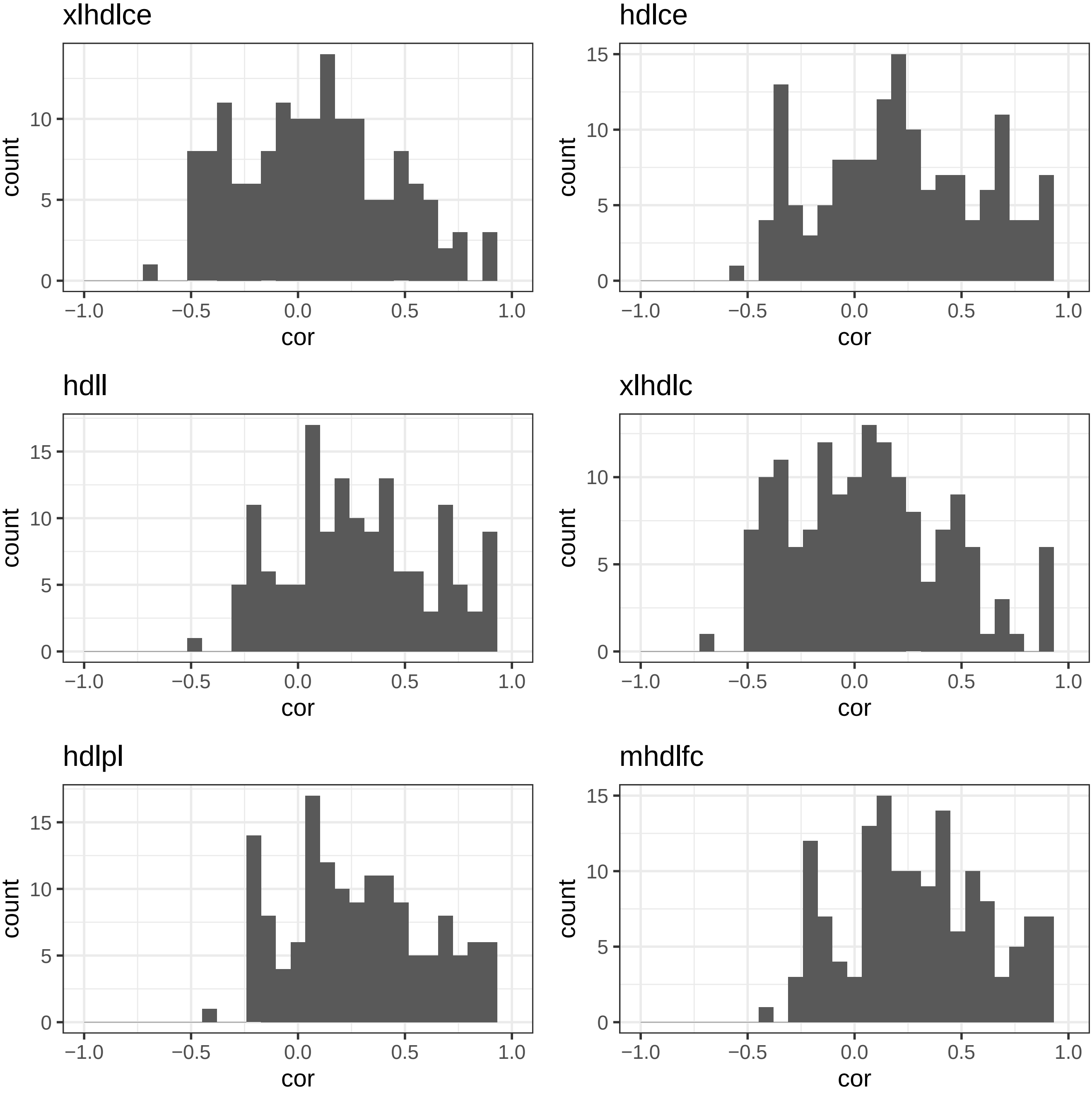
xlhdlce
count
cor
0
5
10
−1.0
−0.5
0.0
0.5
1.0
hdlce
count
cor
0
5
10
15
−1.0
−0.5
0.0
0.5
1.0
hdll
count
cor
0
5
10
15
−1.0
−0.5
0.0
0.5
1.0
xlhdlc
count
cor
0
5
10
−1.0
−0.5
0.0
0.5
1.0
hdlpl
count
cor
0
5
10
15
−1.0
−0.5
0.0
0.5
1.0
mhdlfc
count
cor
0
5
10
15
−1.0
−0.5
0.0
0.5
1.0

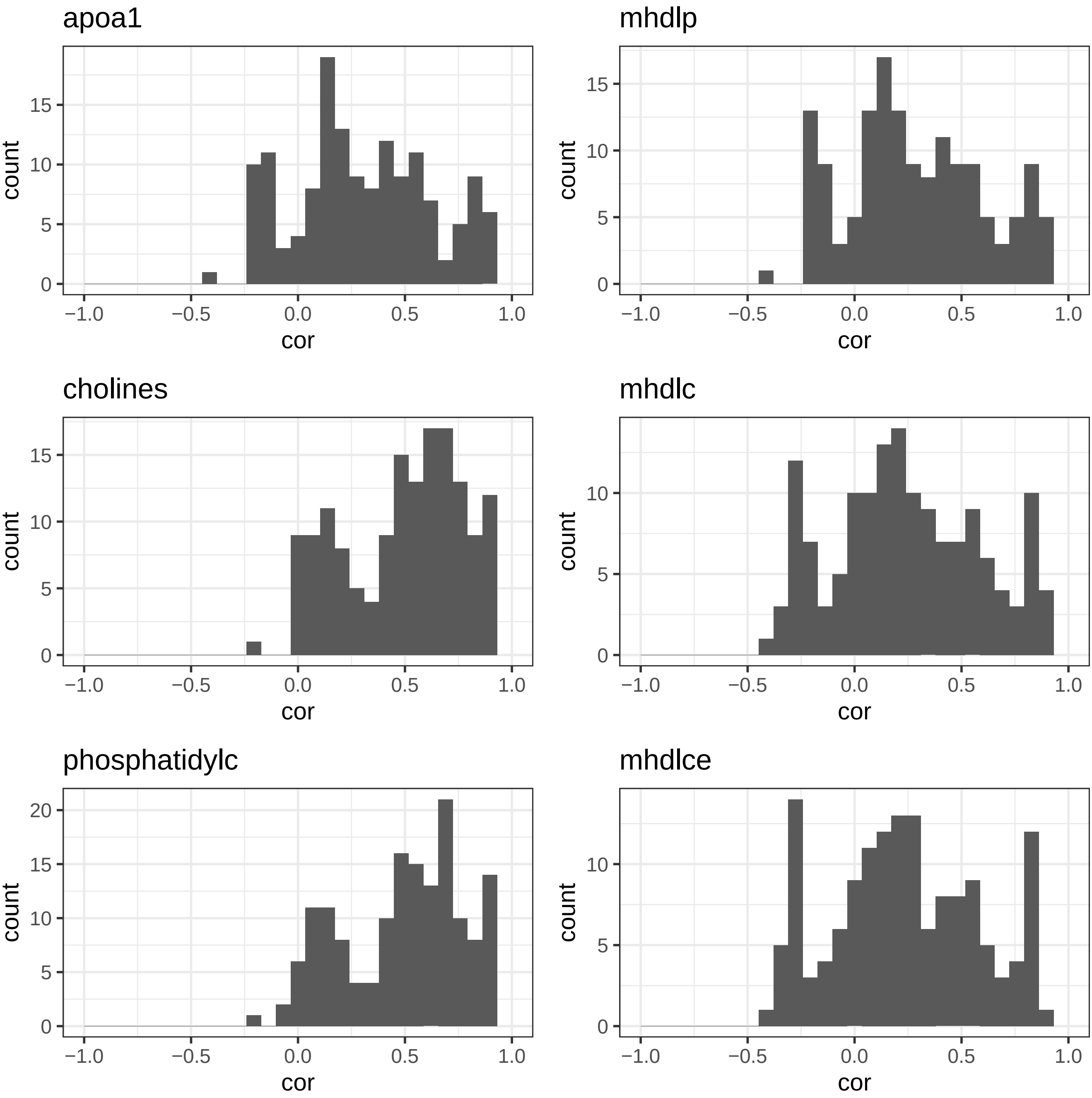
apoa1
mhdlp
cholines
mhdlc
phosphatidylc
mhdlce
count
cor
−1.0
−0.5
0.0
0.5
1.0
0
5
10
15
20

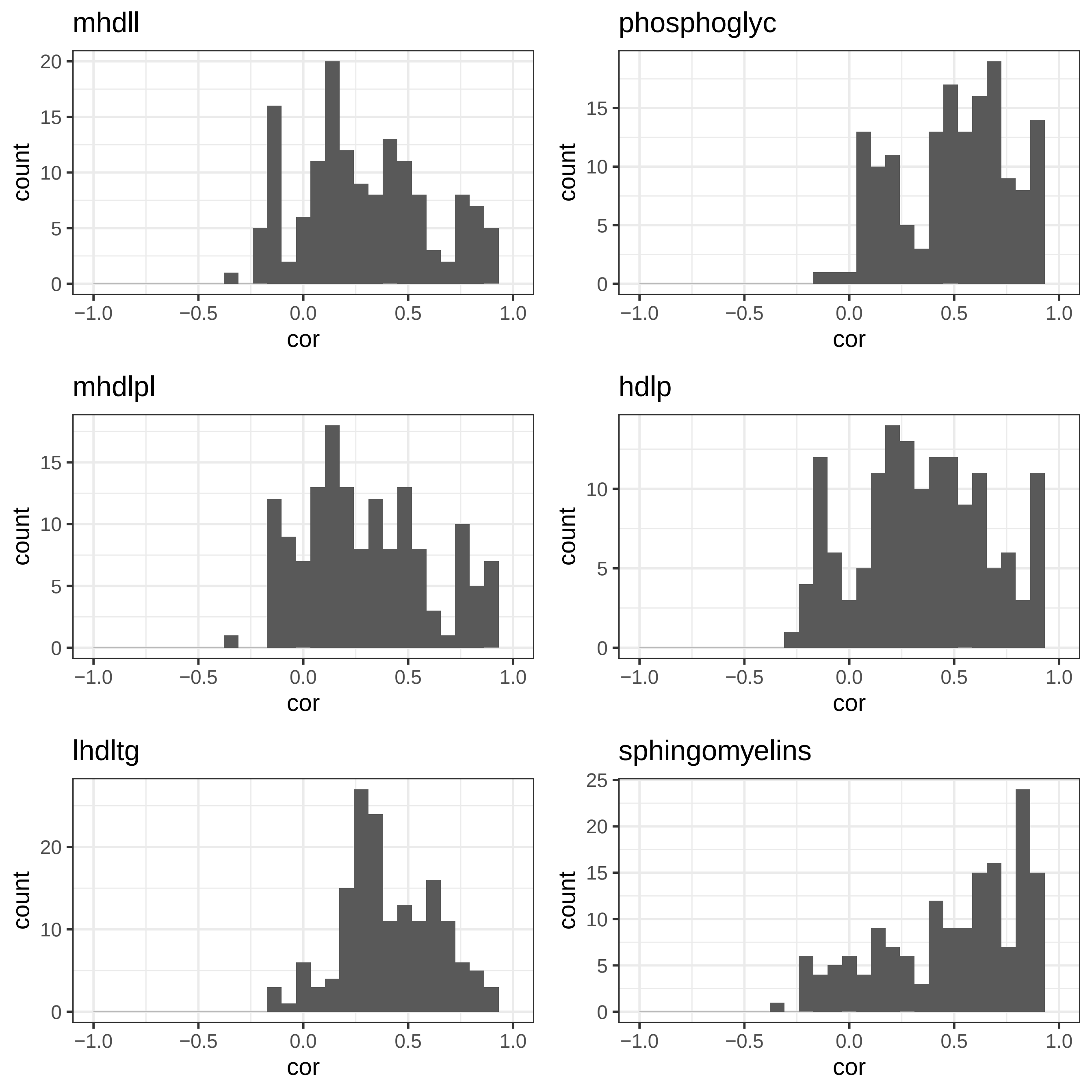
mhdll
phosphoglyc
mhdlpl
hdlp
lhdltg
sphingomyelins
count
cor
−1.0
−0.5
0.0
0.5
1.0

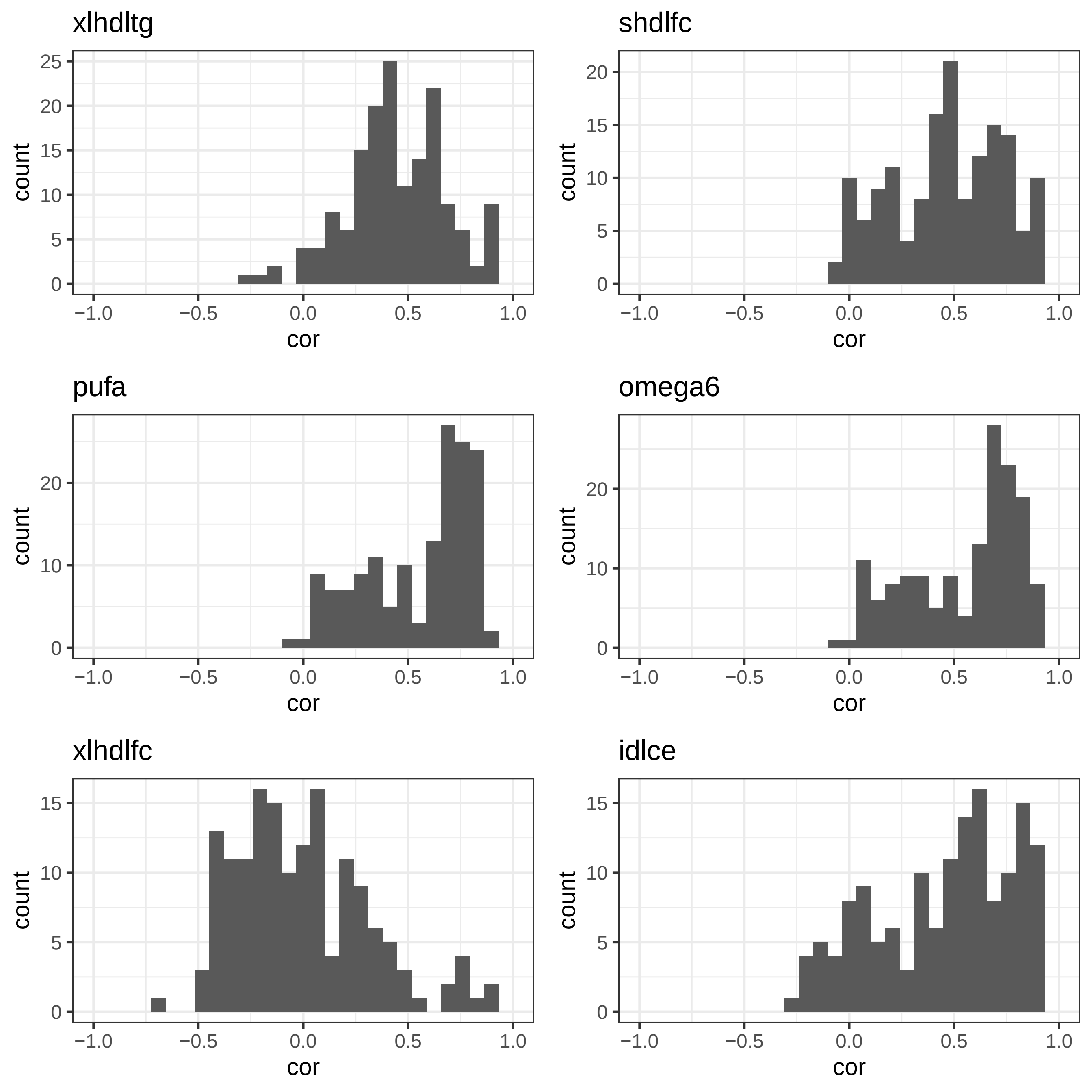
xlhdltg
shdlfc
pufa
omega6
xlhdlfc
idlce
count
cor
-1.0
-0.5
0.0
0.5
1.0

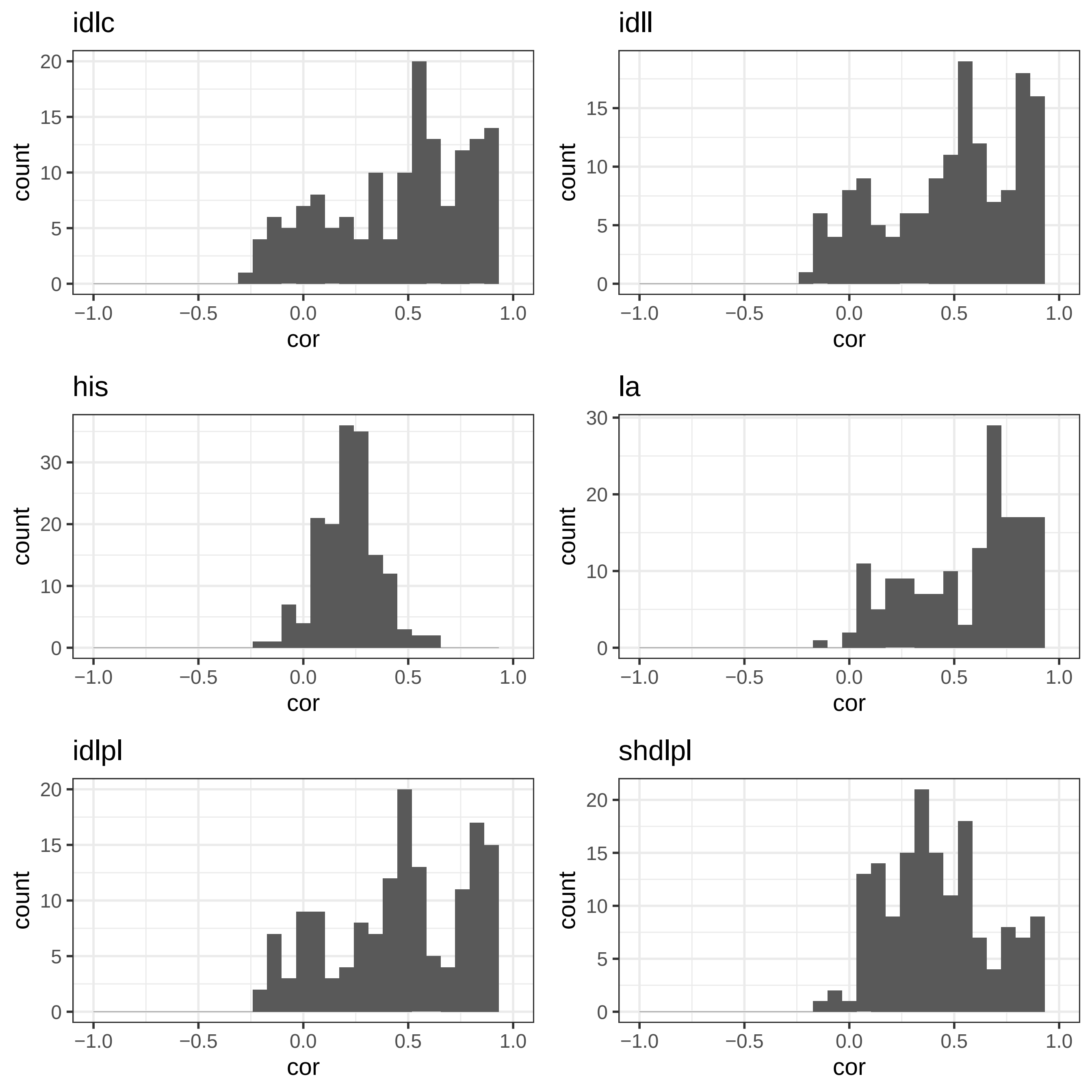

idlc
idll
his
la
idlpl
shdlpl
count
cor
−1.0
−0.5
0.0
0.5
1.0

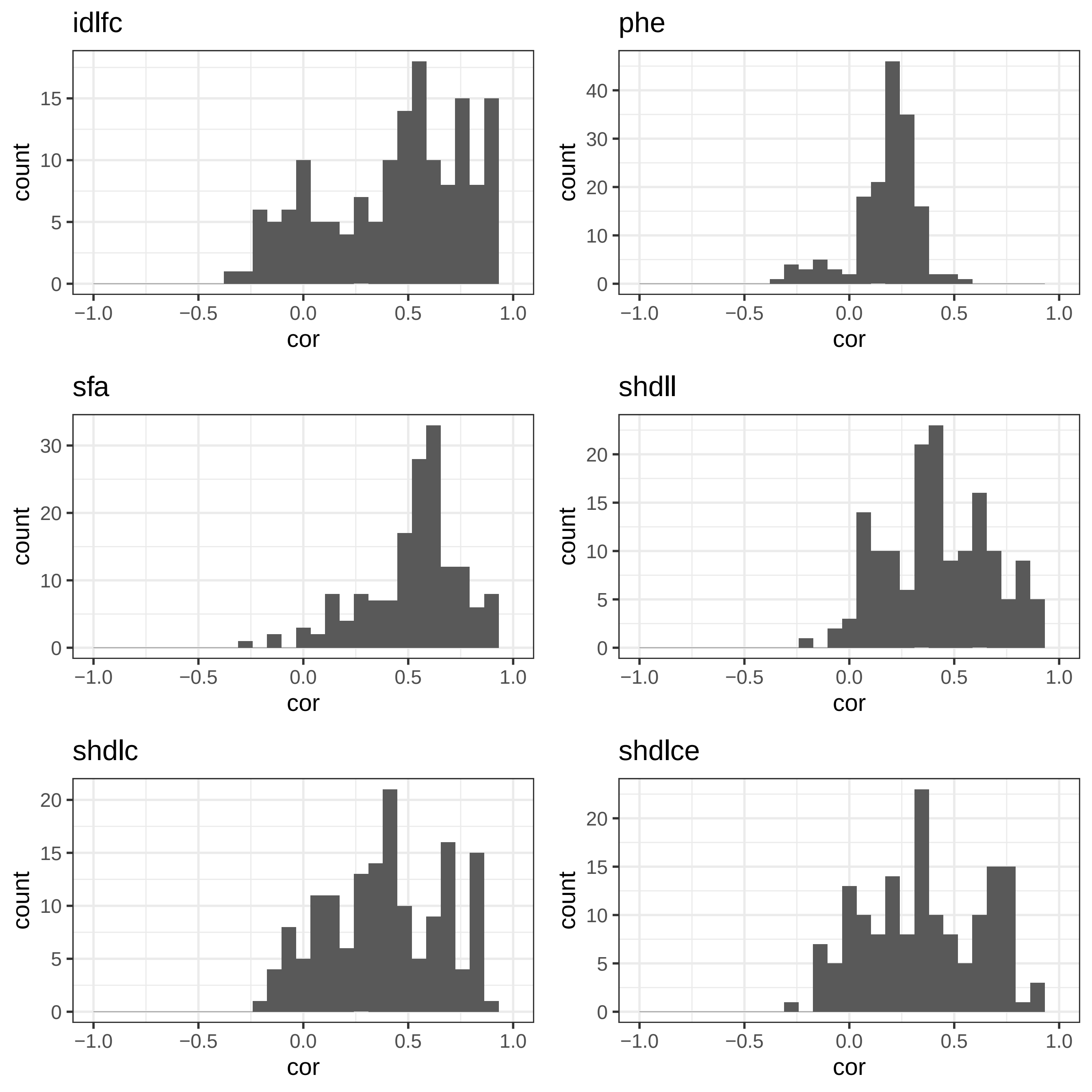

idlfc
phe
sfa
shdll
shdlc
shdlce
count
cor

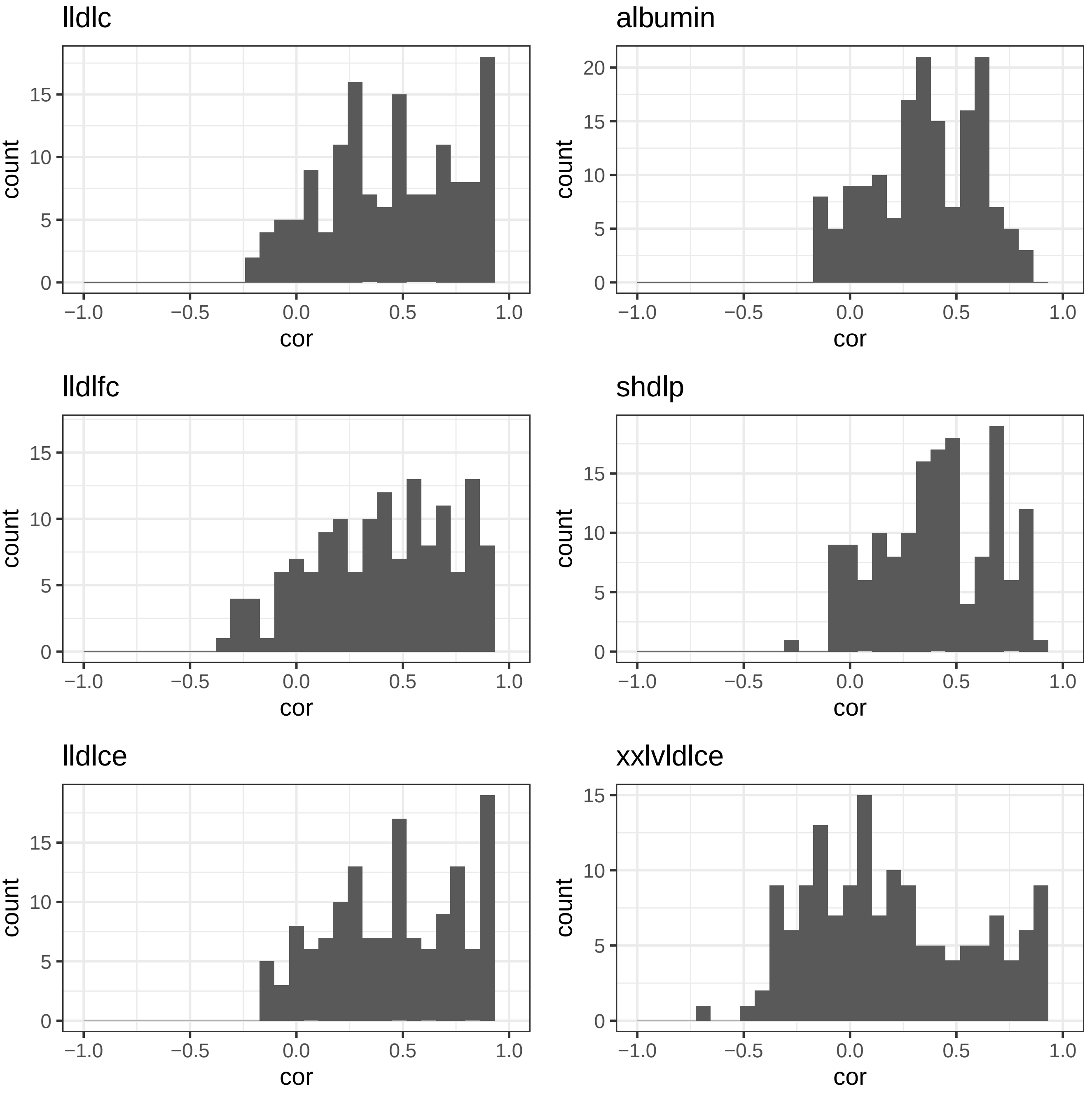

lldlc
albumin
lldlfc
shdlp
lldlce
xxlvldlce
count
cor
0
5
10
15
20
−1.0
−0.5
0.0
0.5
1.0

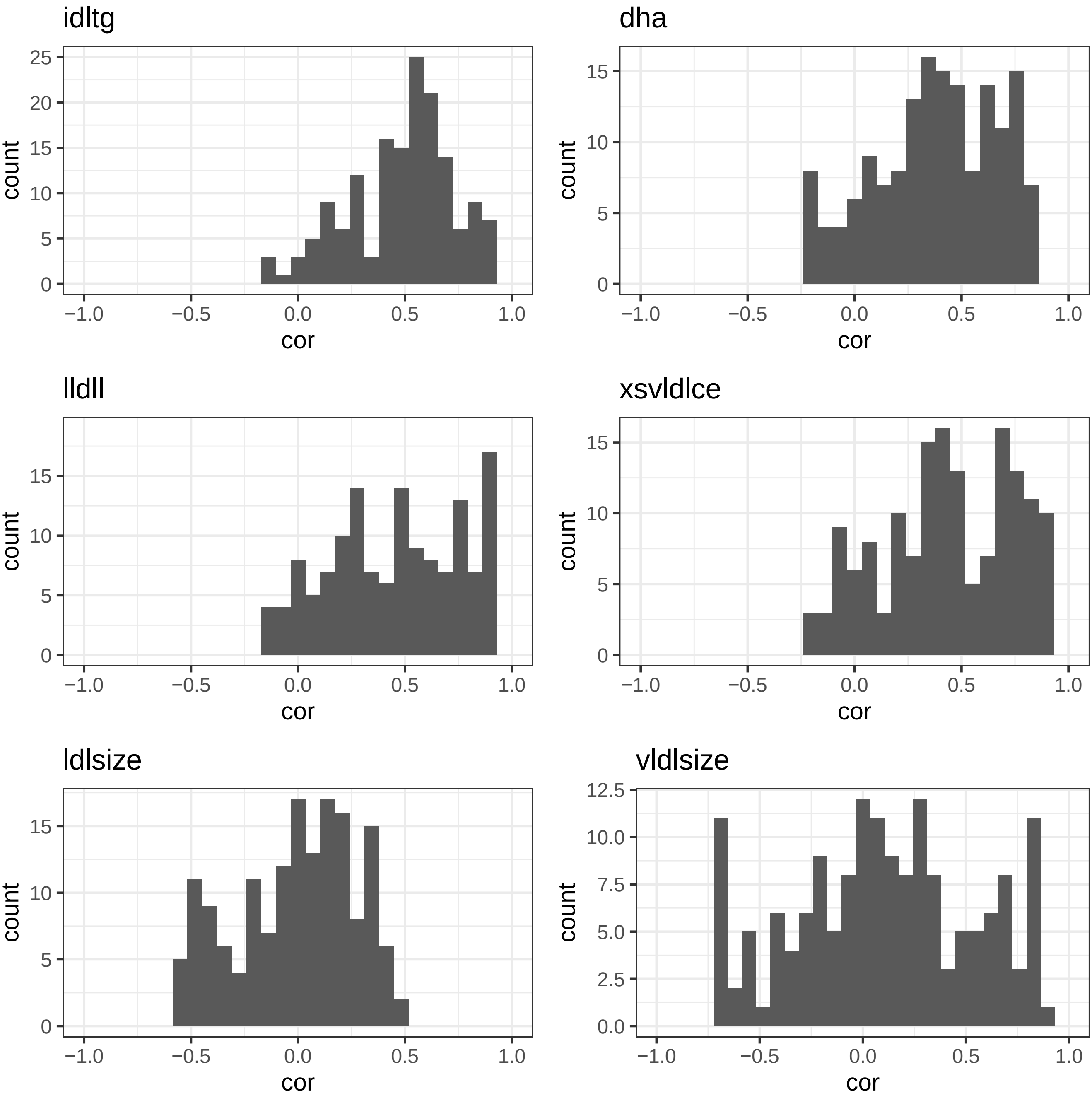
idltg
dha
lldll
xsvldlce
ldlsize
vldlsize
count
cor
−1.0
−0.5
0.0
0.5
1.0
0
5
10
15
20
25
0.0
2.5
5.0
7.5
10.0
12.5

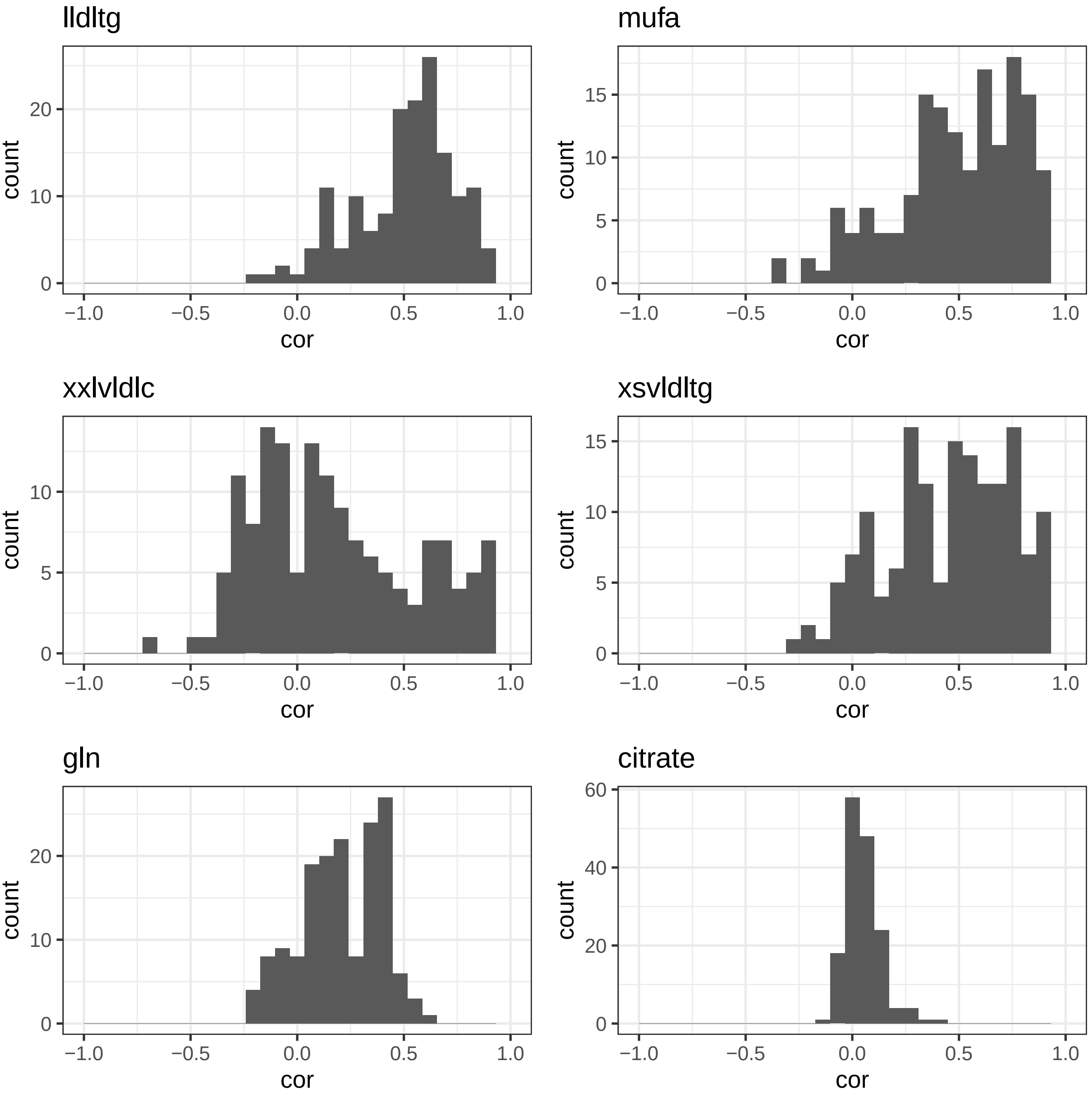
lldltg
mufa
xxlvldlc
xsvldltg
gln
citrate
count
cor
−1.0
−0.5
0.0
0.5
1.0
0
10
20
5
15
40
60

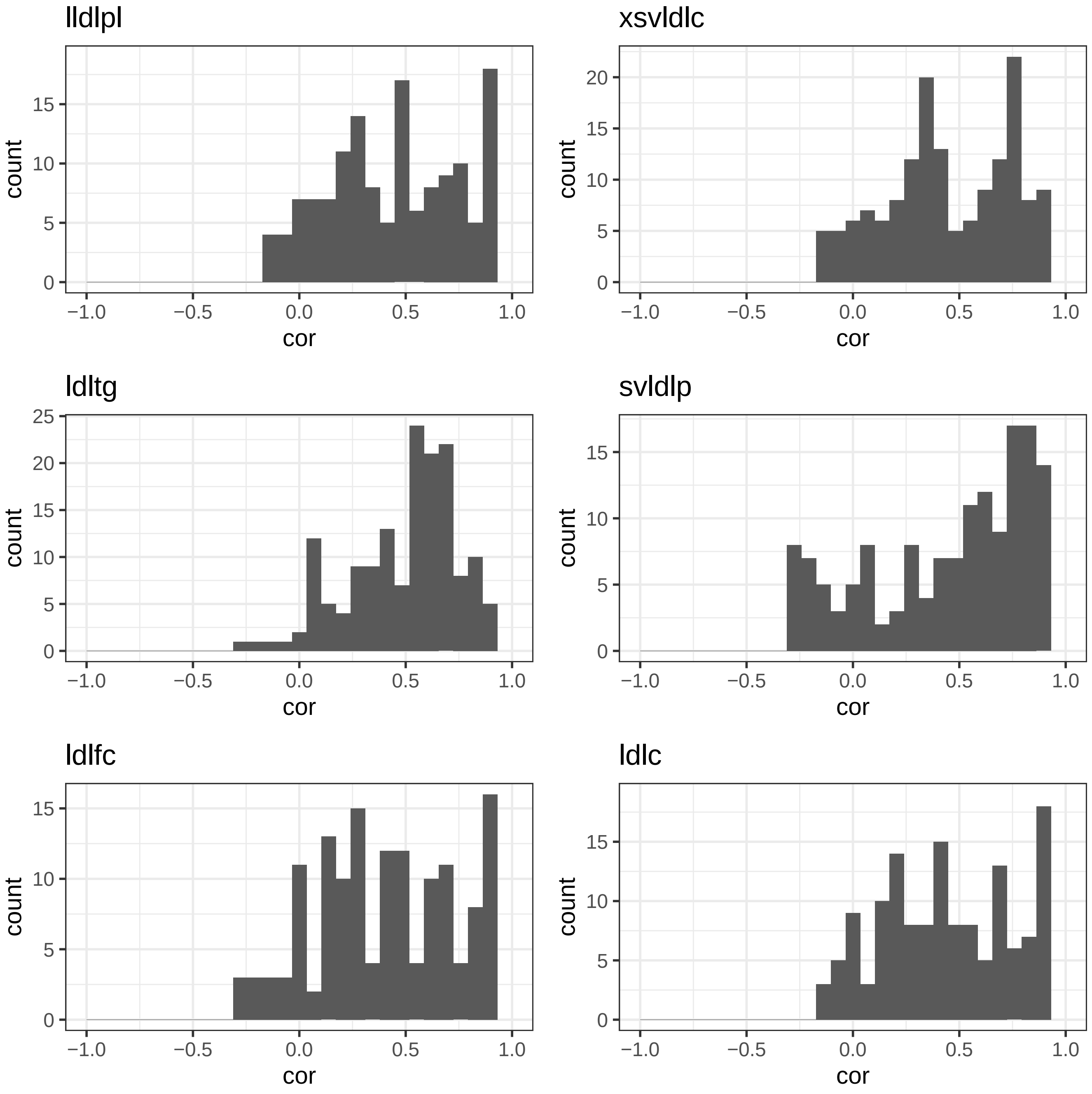

lldlpl
xsvldlc
ldltg
svldlp
ldlfc
ldlc
count
cor

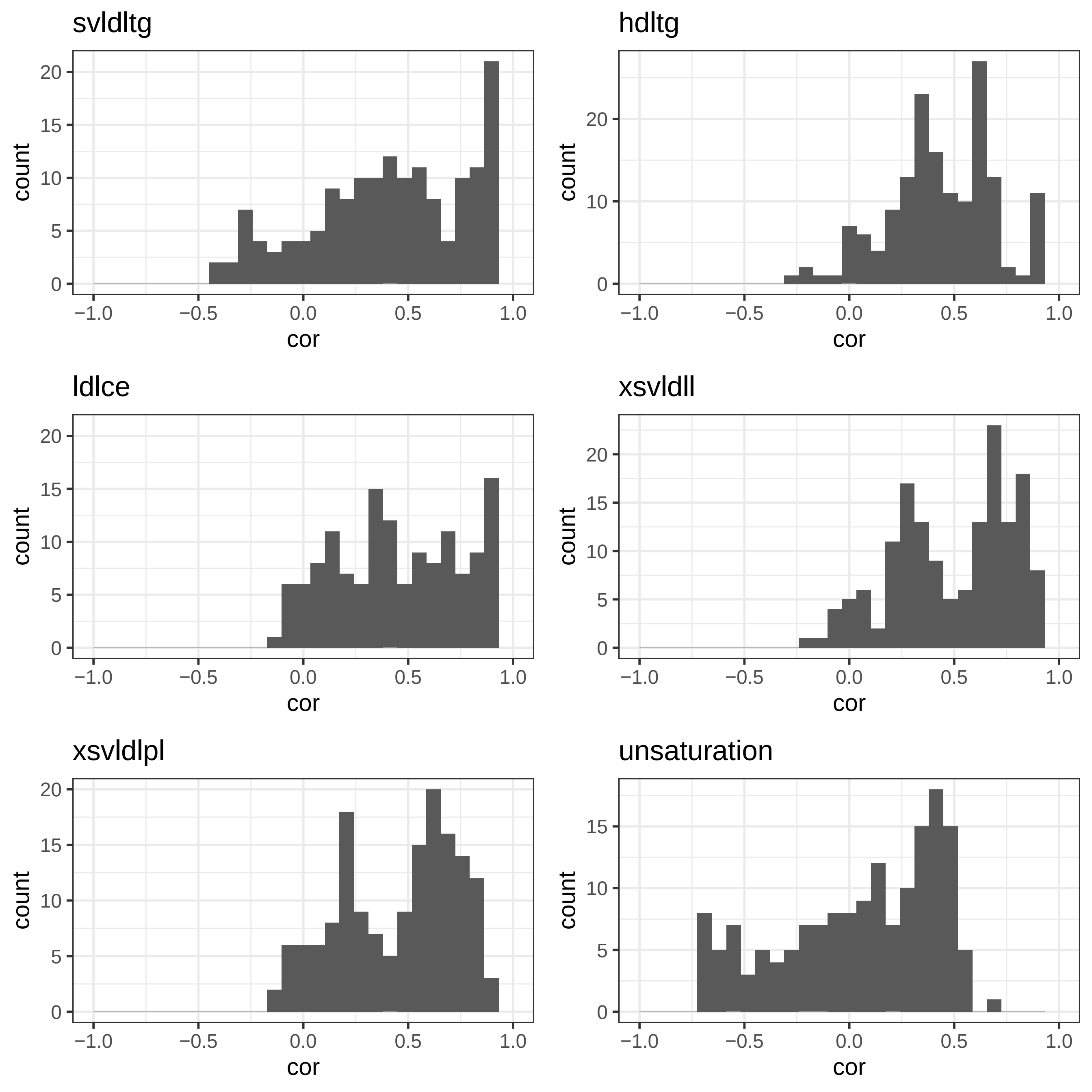

svldltg
hdltg
ldlce
xsvldll
xsvldlpl
unsaturation
count
cor
0
5
10
15
20
−1.0
−0.5
0.0
0.5
1.0

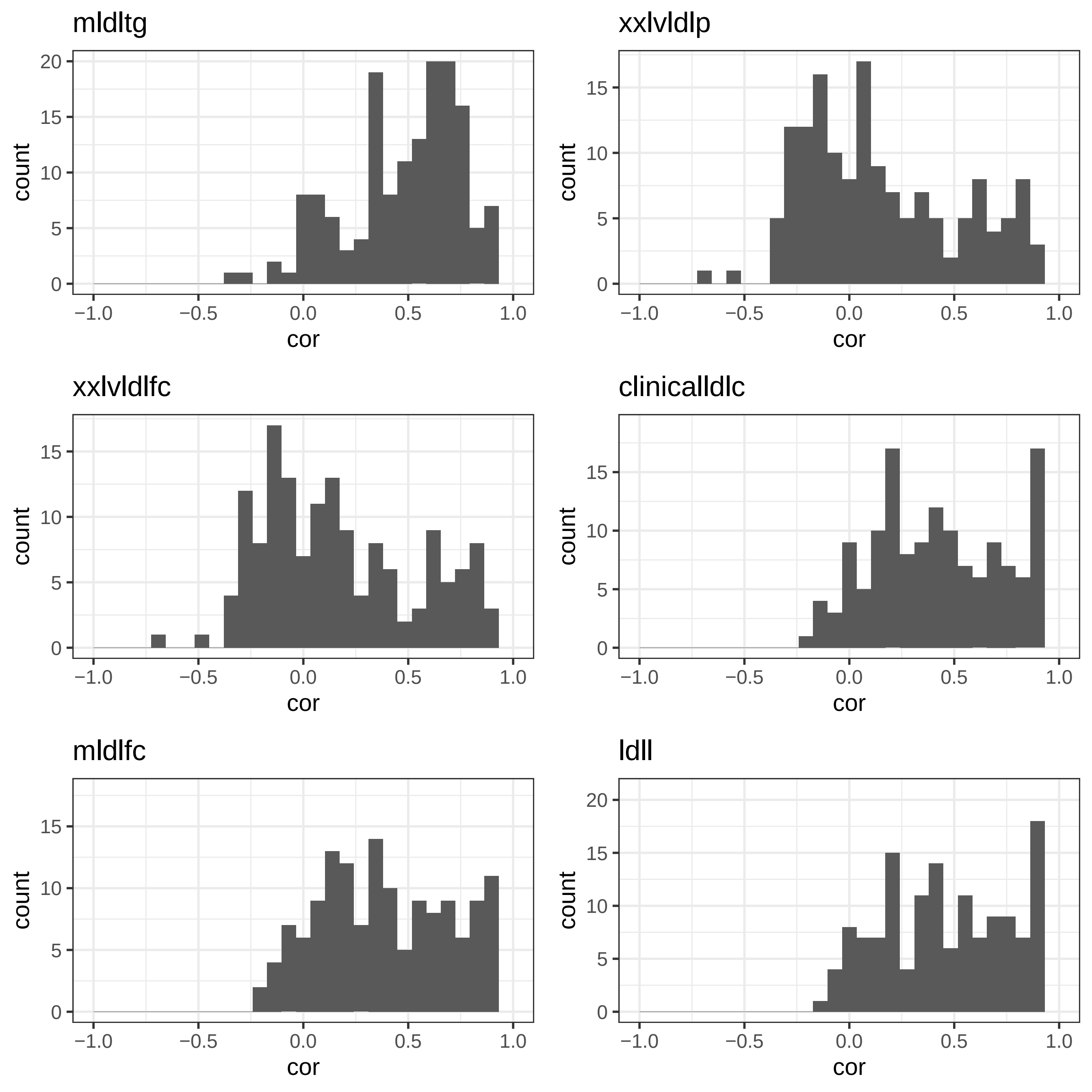
mldltg
xxlvldlp
xxlvldlfc
clinicalldlc
mldlfc
ldll
count
cor
0
5
10
15
20
−1.0
−0.5
0.0
0.5
1.0

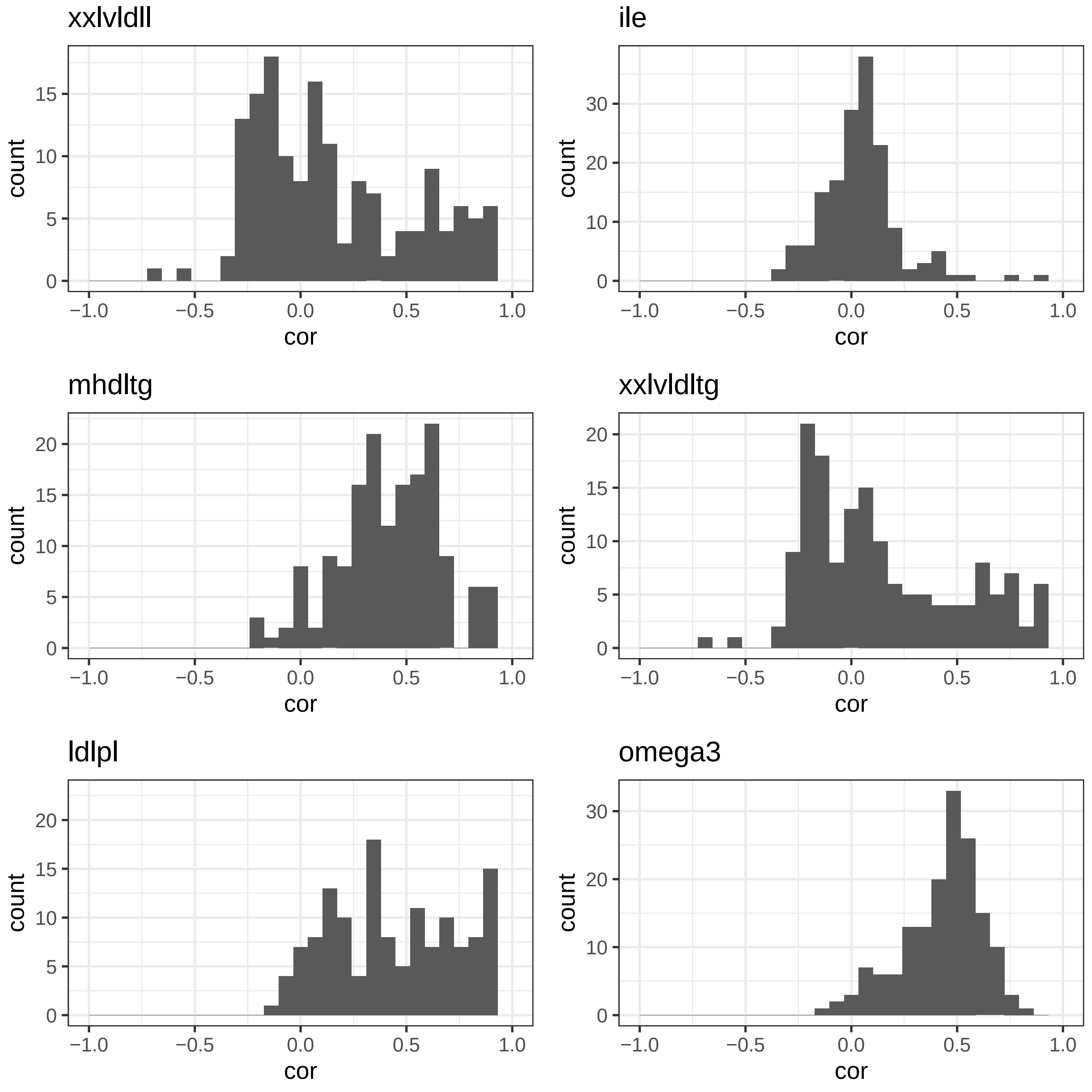
xxlvldll
count
0
5
10
15
−1.0
−0.5
0.0
0.5
1.0
cor
ile
count
0
10
20
30
−1.0
−0.5
0.0
0.5
1.0
cor
mhdltg
count
0
5
10
15
20
−1.0
−0.5
0.0
0.5
1.0
cor
xxlvldltg
count
0
5
10
15
20
−1.0
−0.5
0.0
0.5
1.0
cor
ldlpl
count
0
5
10
15
20
−1.0
−0.5
0.0
0.5
1.0
cor
omega3
count
0
10
20
30
−1.0
−0.5
0.0
0.5
1.0
cor

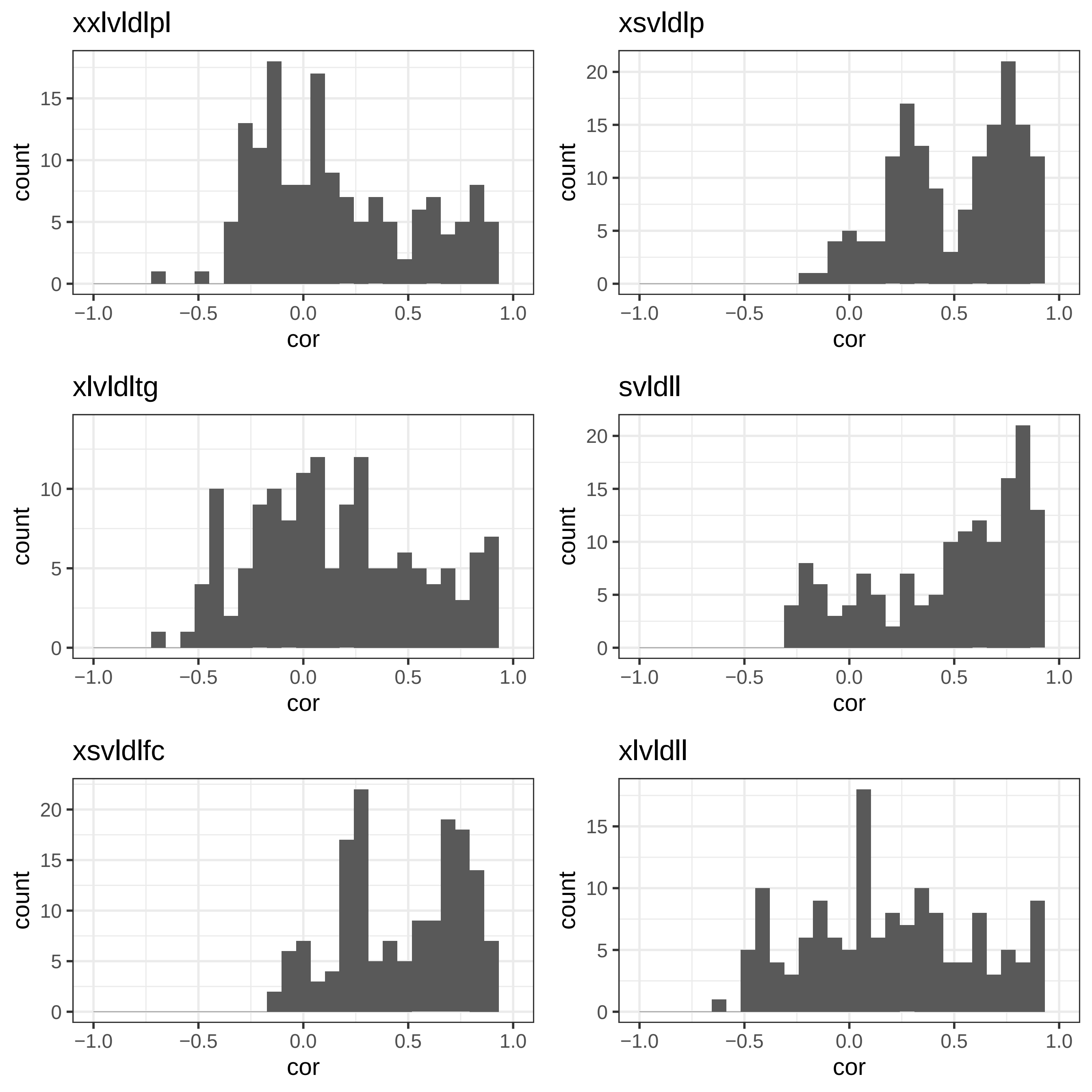
xxlvldlpl
xsvldlp
xlvldltg
svldll
xsvldlfc
xlvldll
count
cor
0
5
10
15
20
−1.0
−0.5
0.0
0.5
1.0

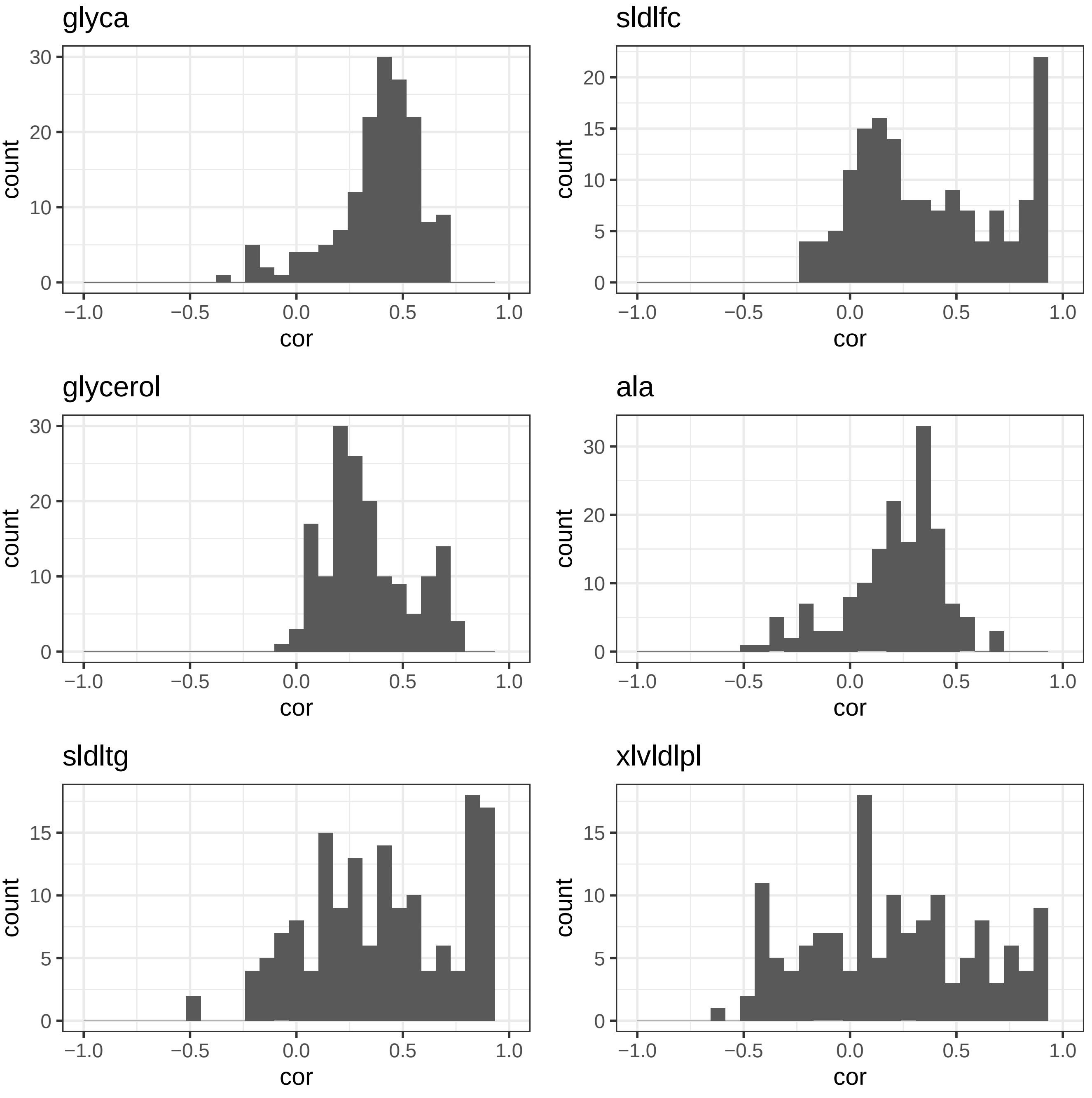
glyca
sldlfc
glycerol
ala
sldltg
xlvldlpl
count
cor
−1.0
−0.5
0.0
0.5
1.0

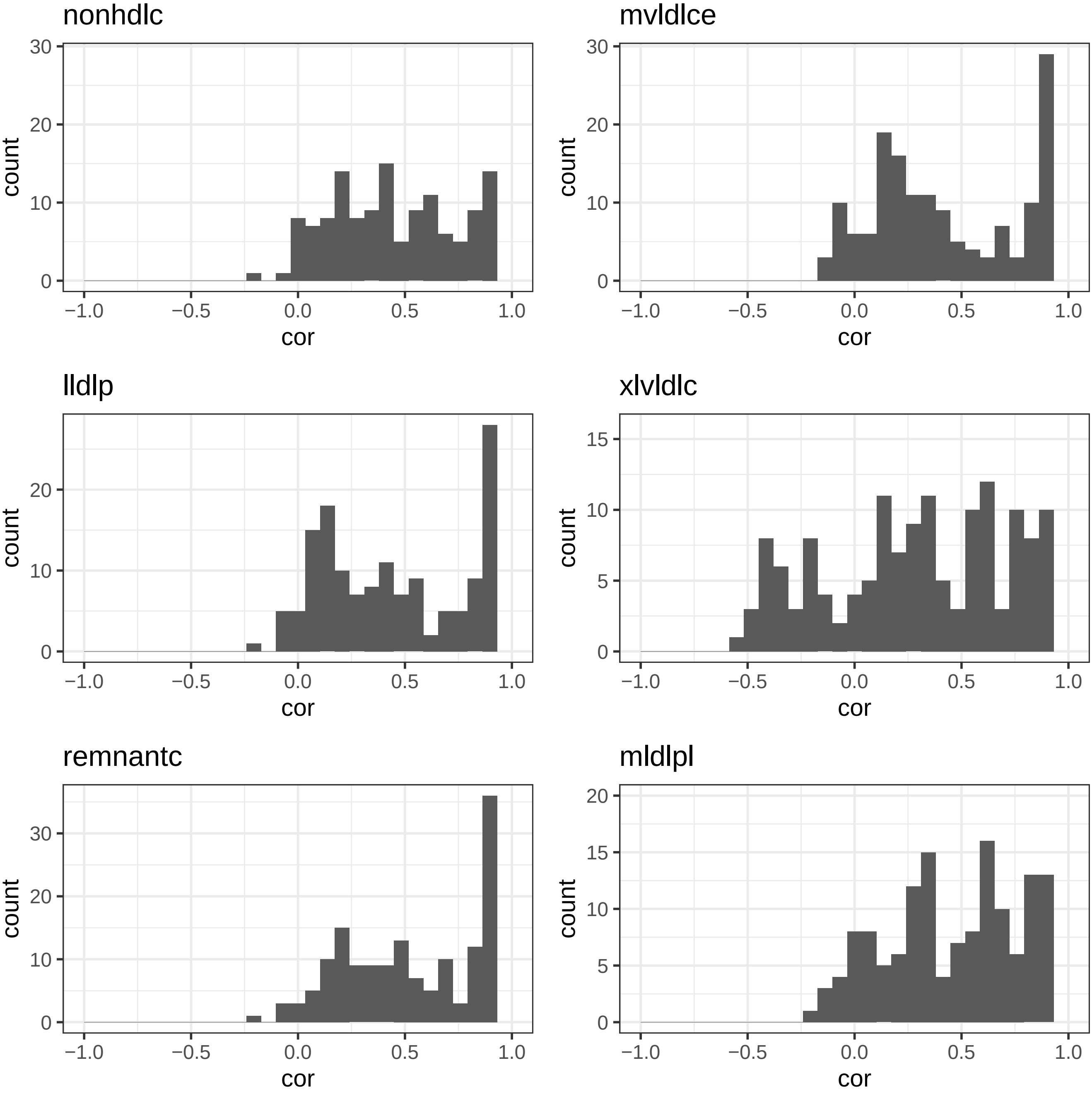
nonhdlc
mvldlce
lldlp
xlvldlc
remnantc
mldlpl
count
cor

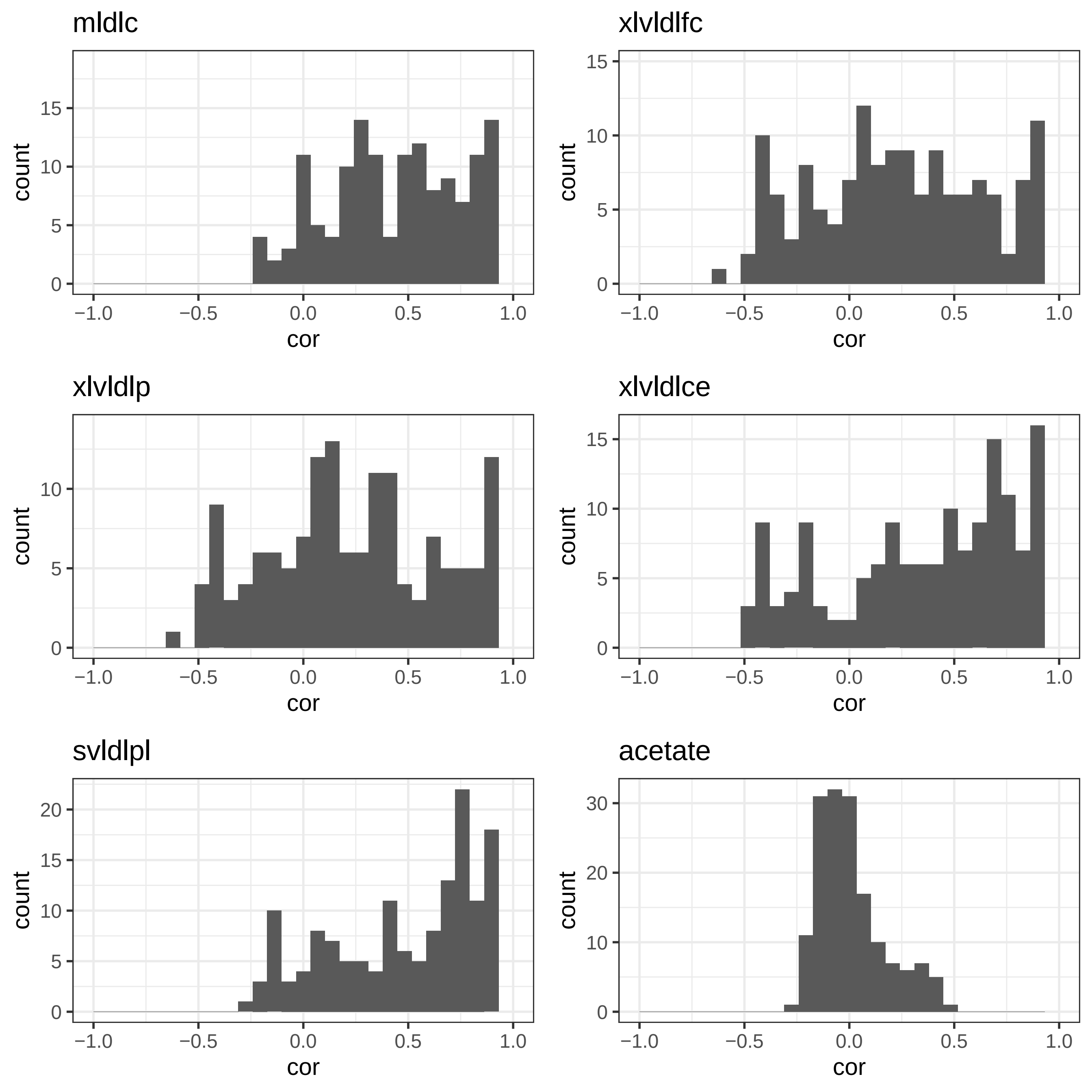

mldlc
xlvldlfc
xlvldlp
xlvldlce
svldlpl
acetate
count
cor

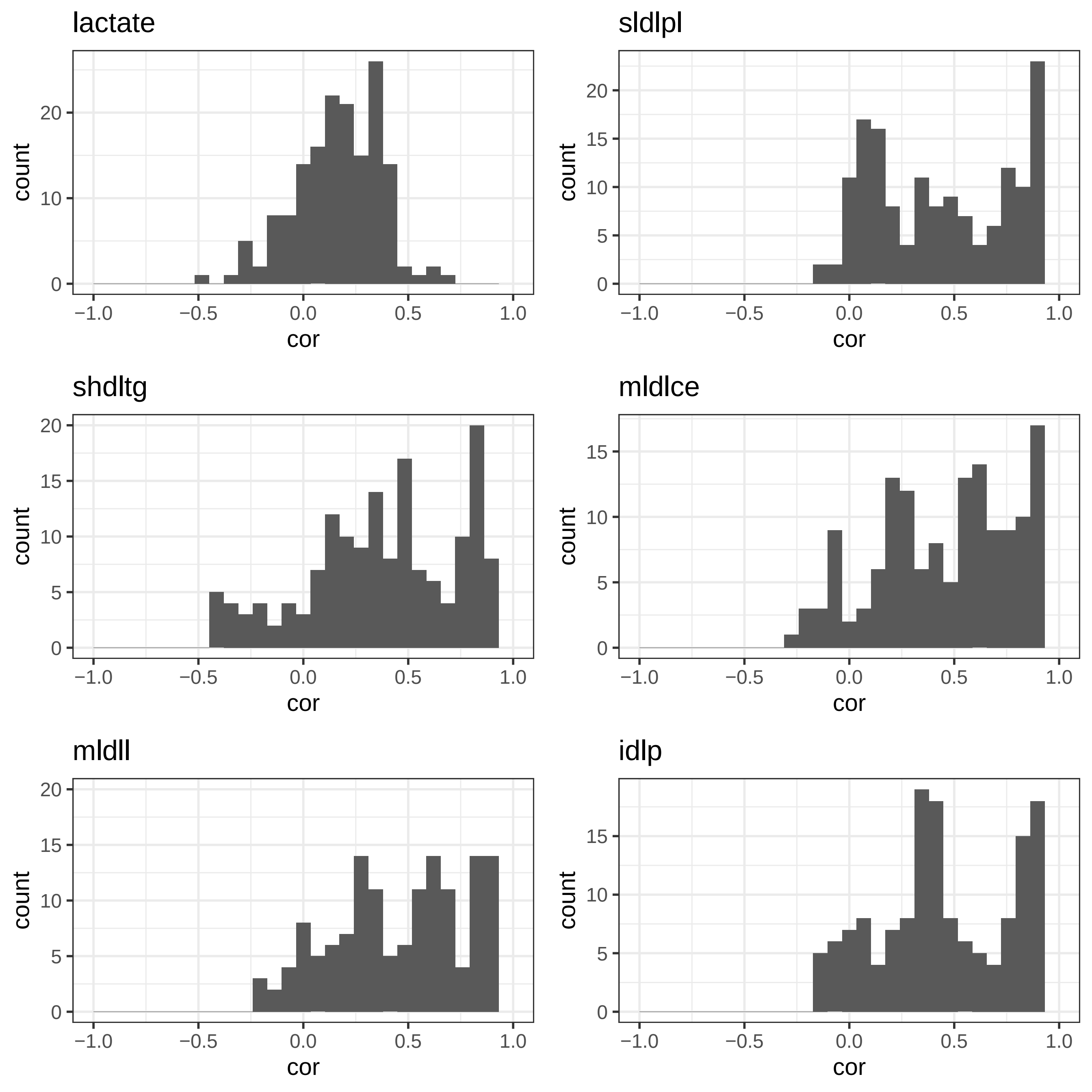

lactate
sldlpl
shdltg
mldlce
mldll
idlp
count
cor
−1.0
−0.5
0.0
0.5
1.0

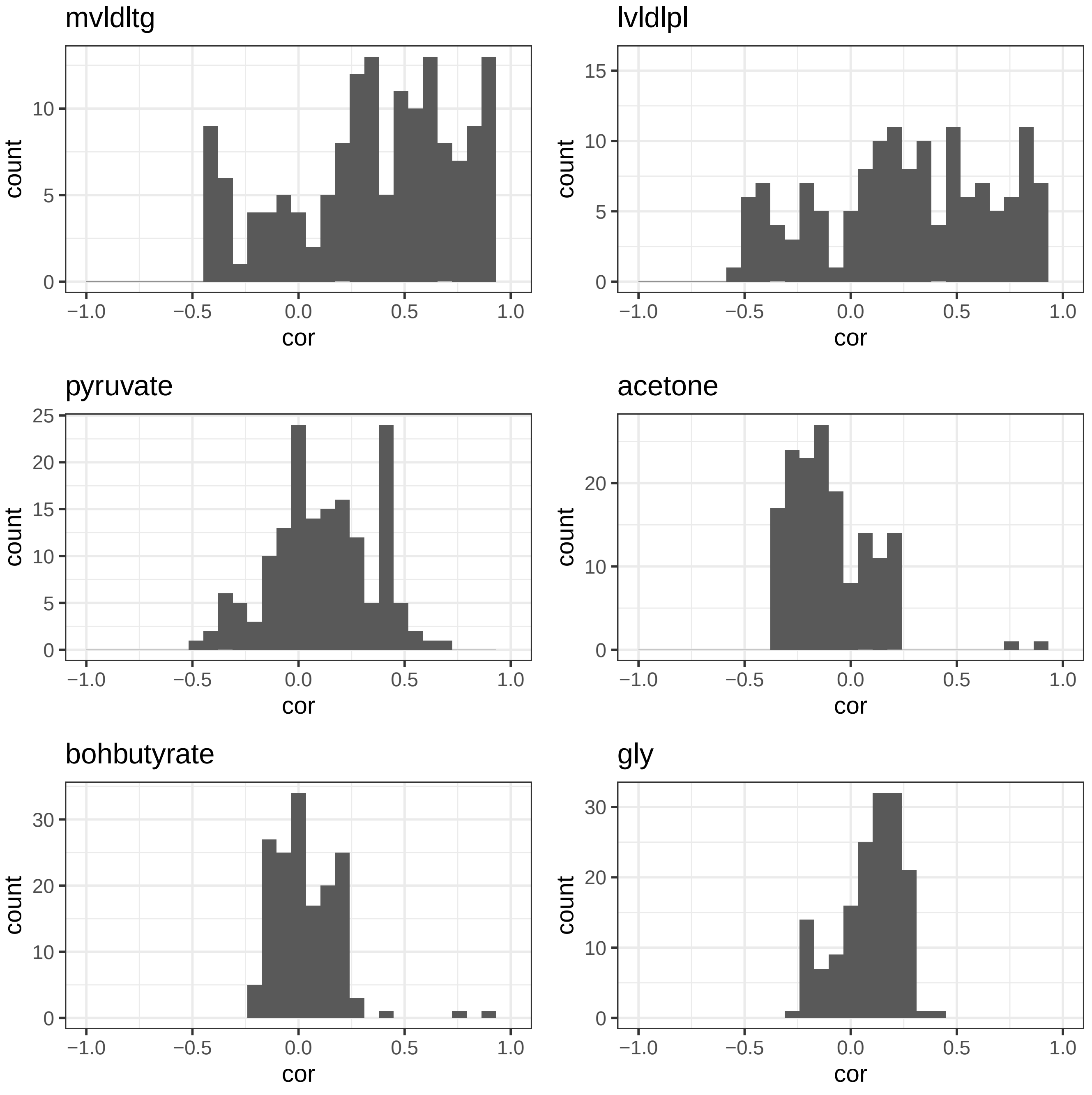
mvldltg
lvldlpl
pyruvate
acetone
bohbutyrate
gly
count
cor
−1.0
−0.5
0.0
0.5
1.0
0
5
10
15
20
25
30

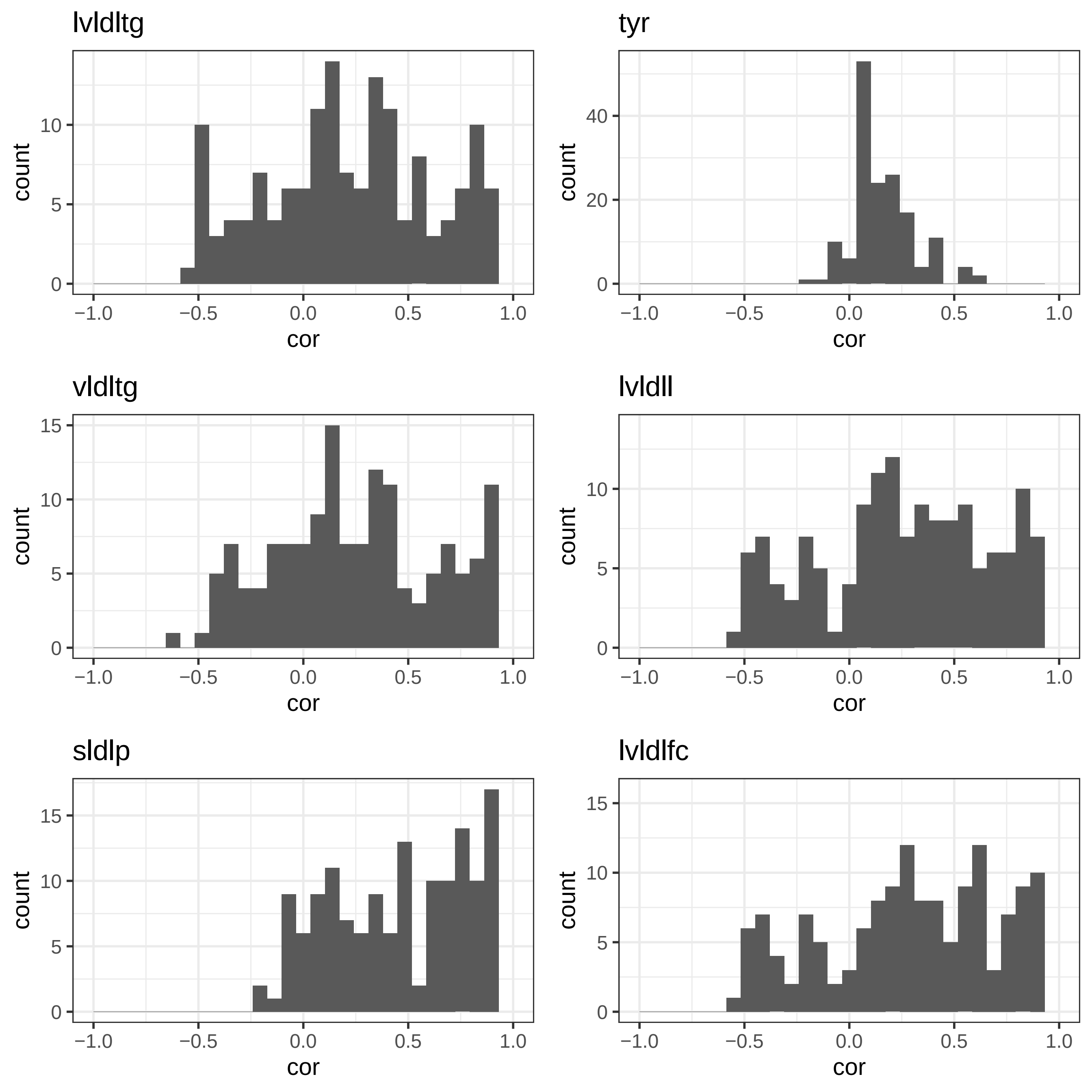

lvldltg
tyr
vldltg
lvldll
sldlp
lvldlfc
count
cor

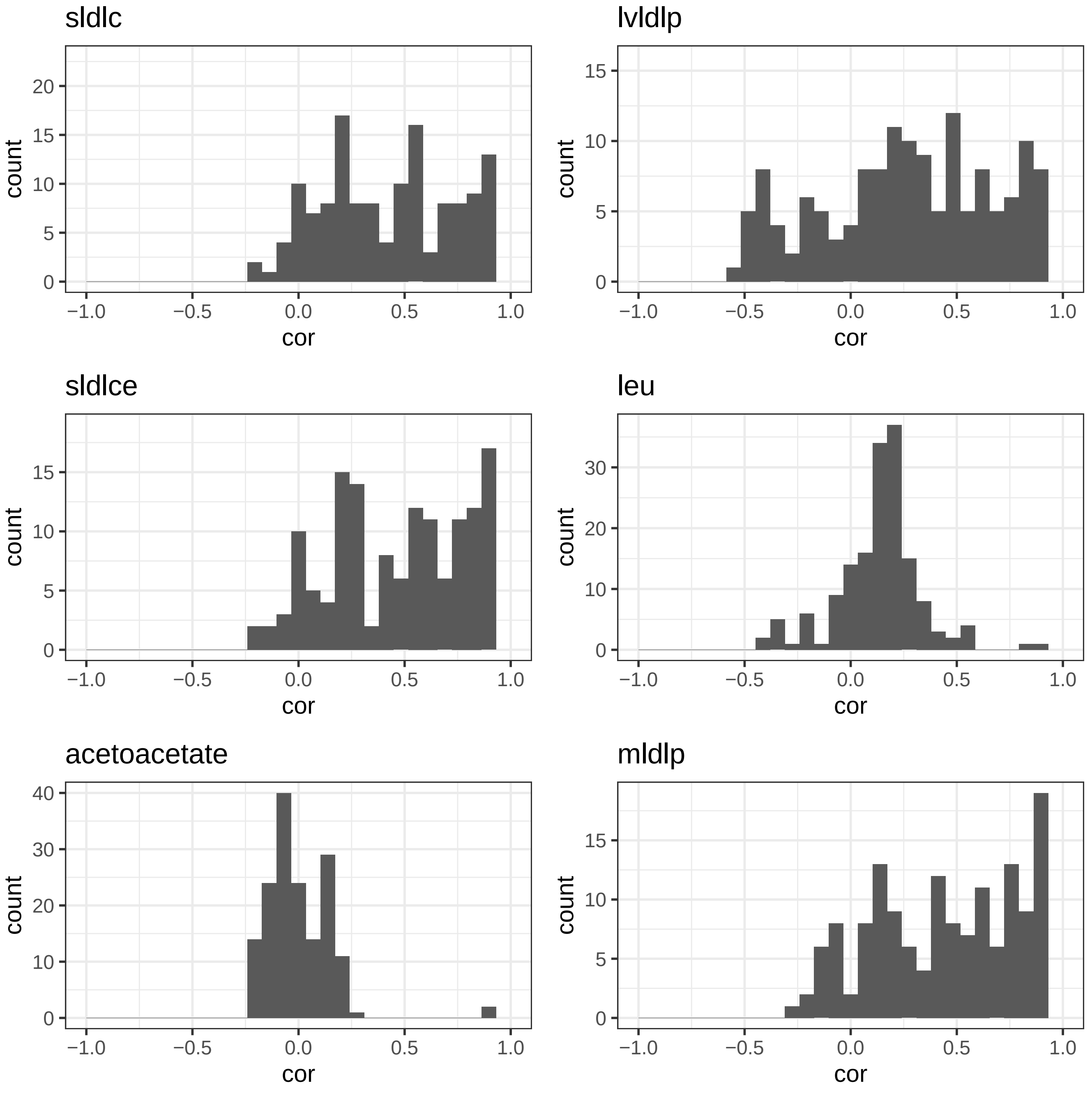
sldlc
lvldlp
sldlce
leu
acetoacetate
mldlp
count
cor
−1.0
−0.5
0.0
0.5
1.0
0
5
10
15
20
30
40

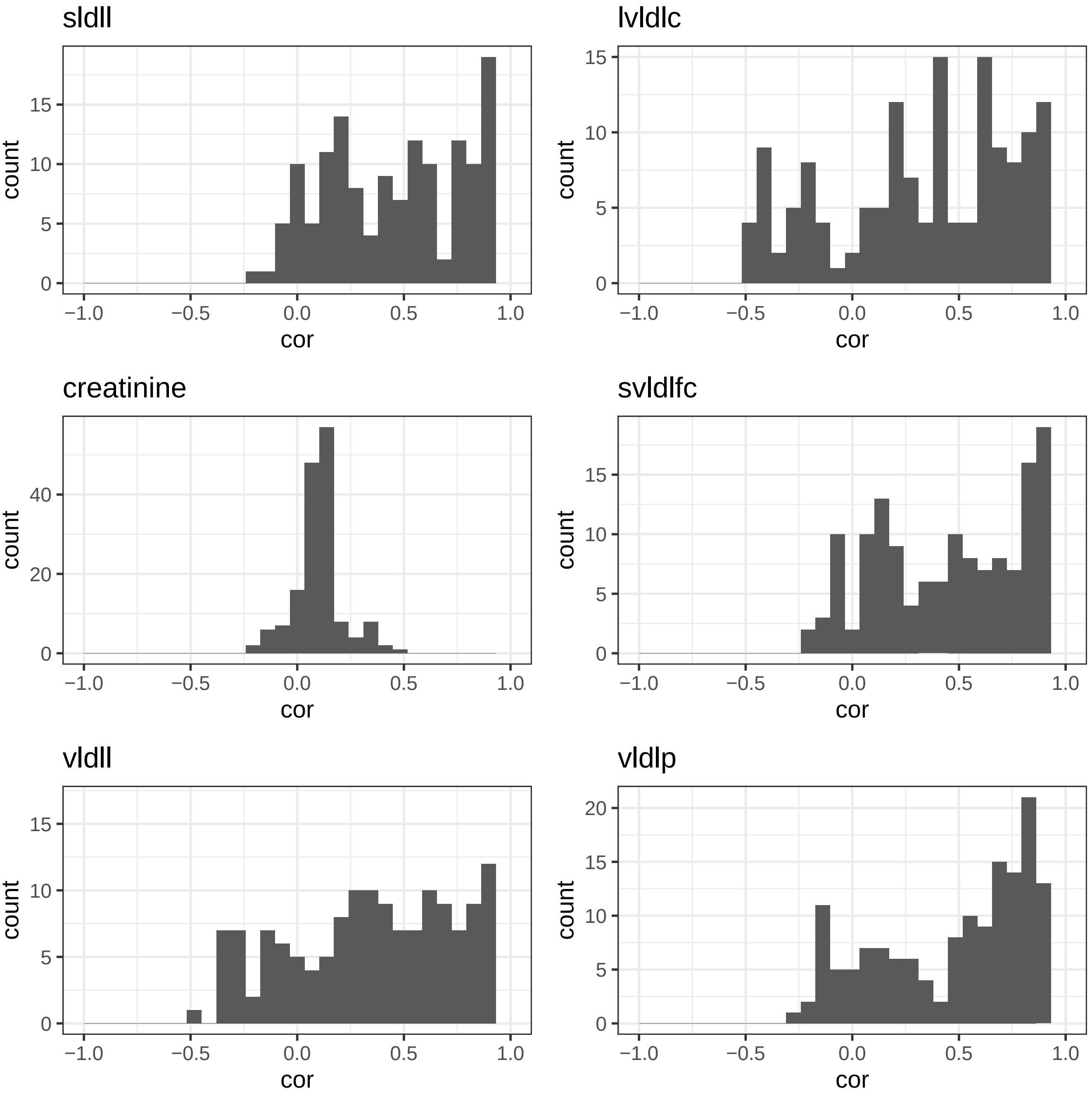
sldll
lvldlc
creatinine
svldlfc
vldll
vldlp
count
cor
-1.0
-0.5
0.0
0.5
1.0

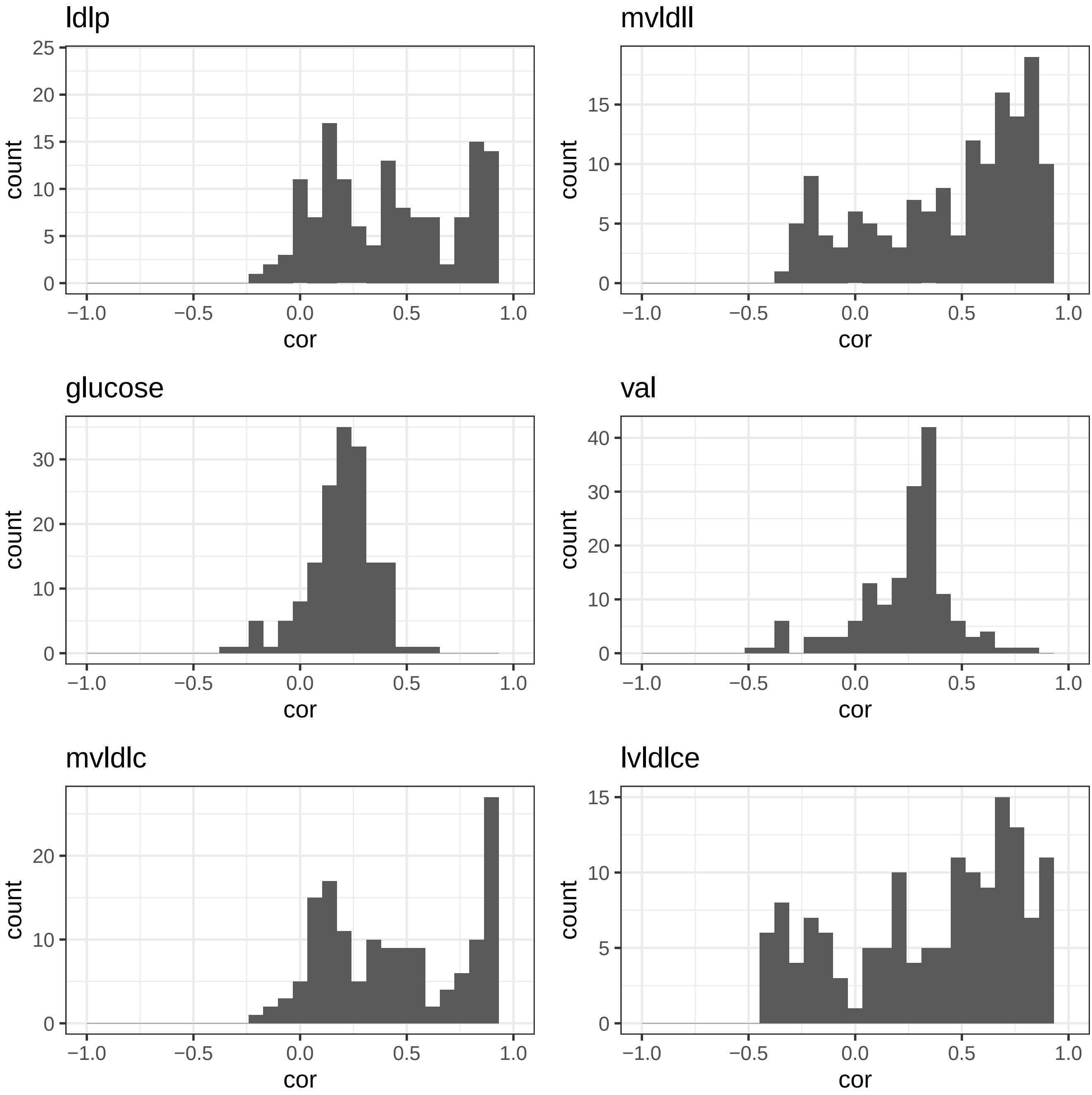
ldlp
mvldll
glucose
val
mvldlc
lvldlce
count
cor
−1.0
−0.5
0.0
0.5
1.0
0
5
10
15
20
25
30
40

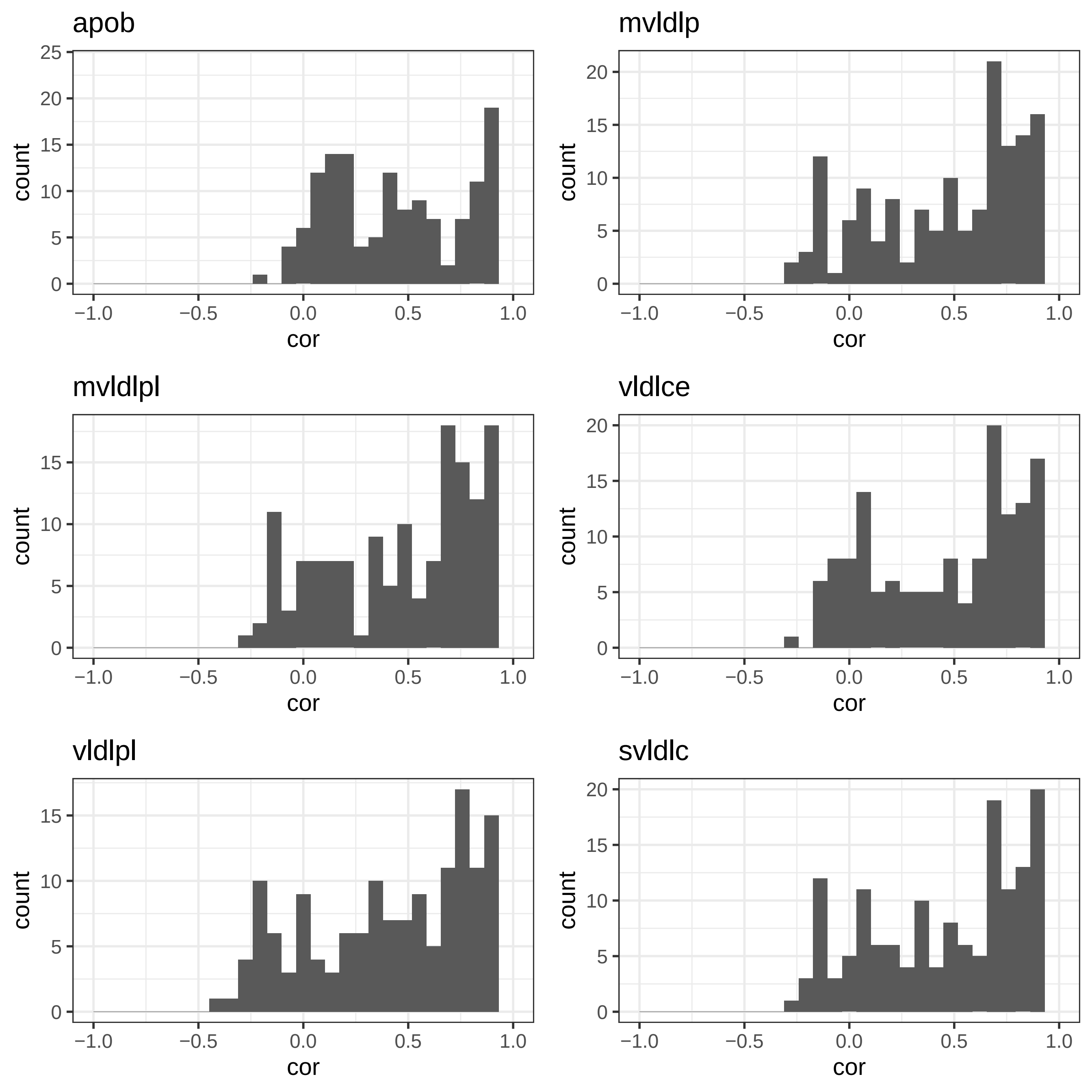
apob
mvldlp
mvldlpl
vldlce
vldlpl
svldlc
count
cor
−1.0
−0.5
0.0
0.5
1.0
0
5
10
15
20
25

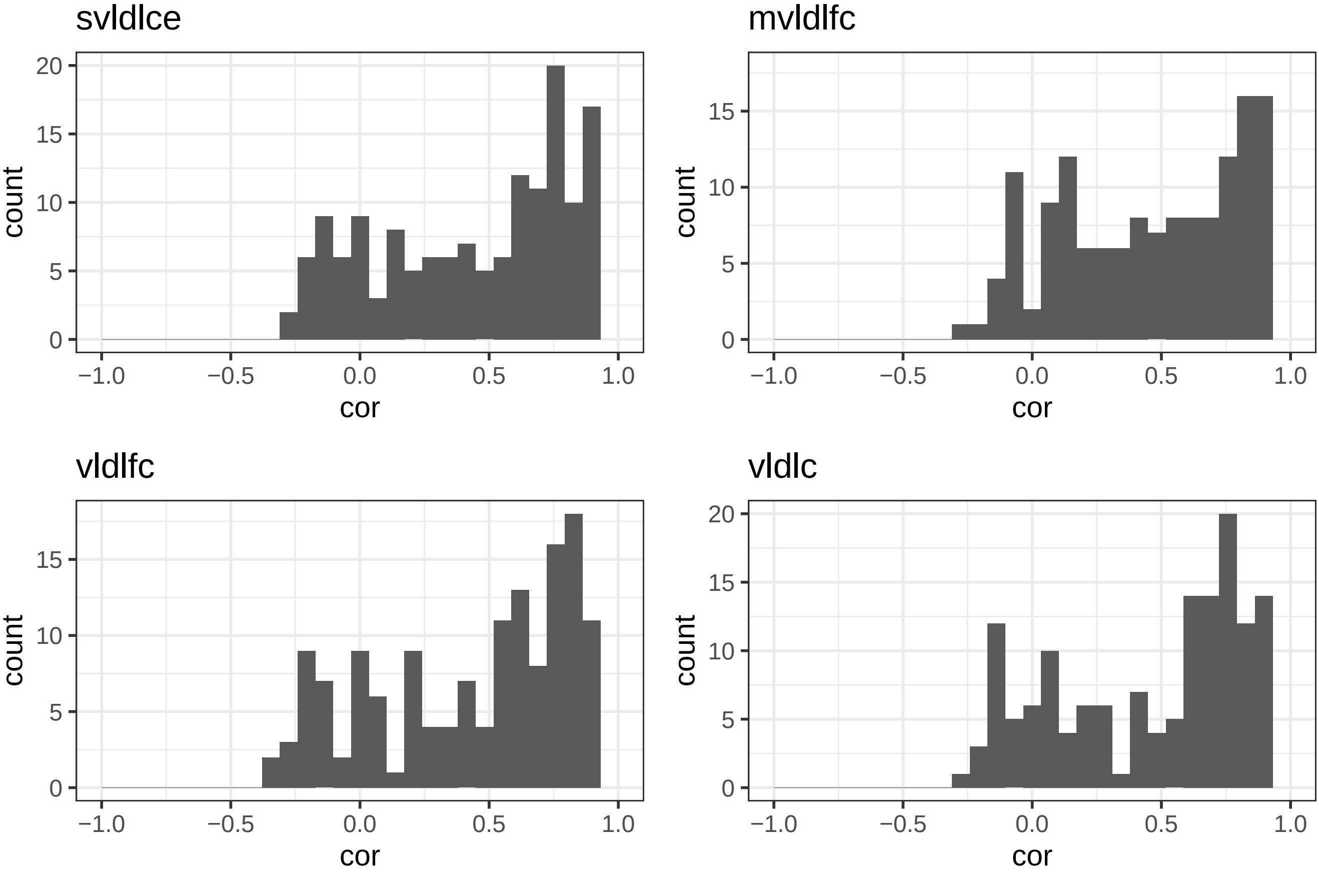
svldlce
mvldlfc
vldlfc
vldlc
count
cor
0
5
10
15
20
−1.0
−0.5
0.0
0.5
1.0
