## Supplementary figures and images for "Multivariate analysis of metabolomic data to identify biological pathways modified by a clinical intervention"

### Supplementary Data 5

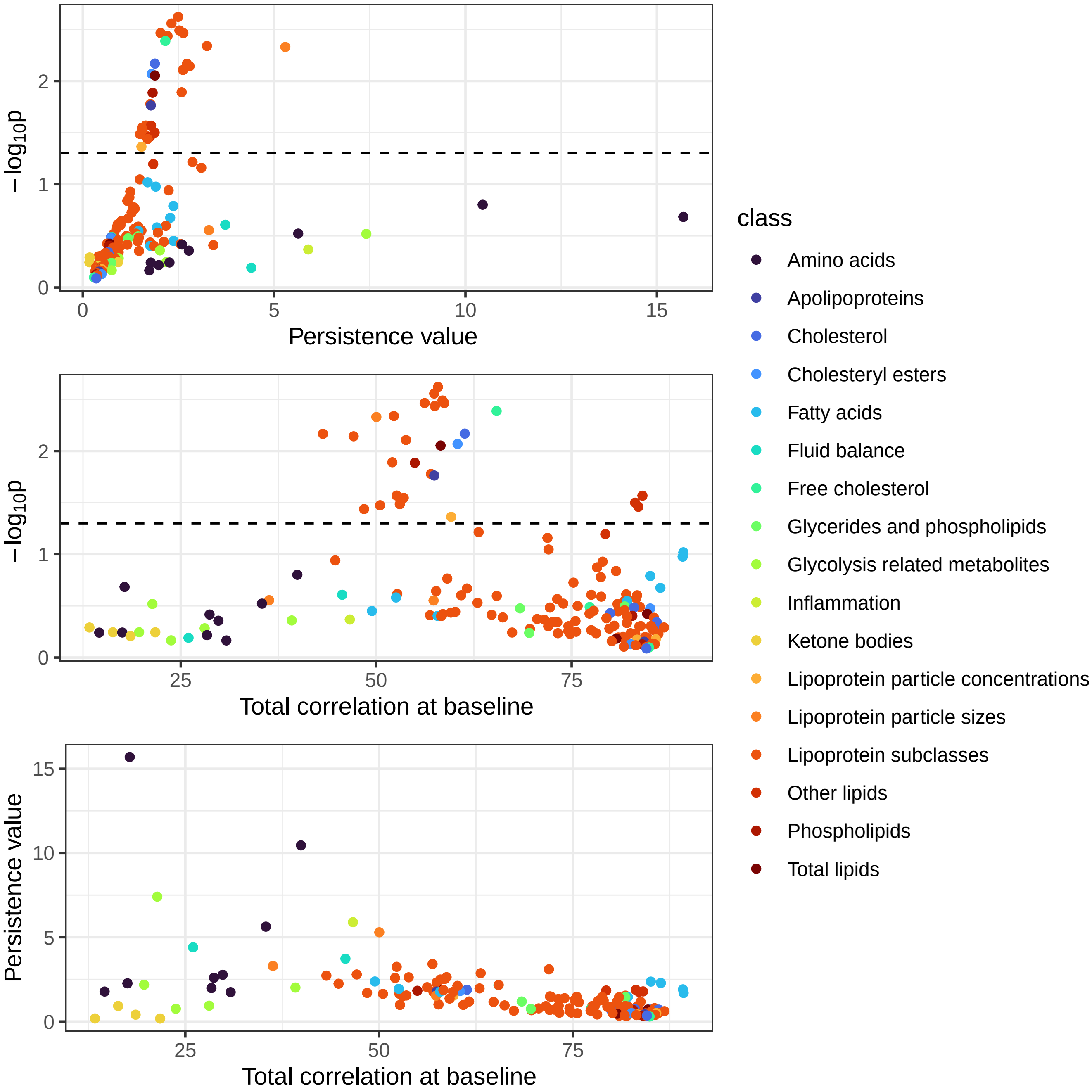
